# Prompting is all you need: LLMs for systematic review screening

**DOI:** 10.1101/2024.06.01.24308323

**Authors:** Christian Cao, Jason Sang, Rohit Arora, Robbie Kloosterman, Matt Cecere, Jaswanth Gorla, Richard Saleh, David Chen, Ian Drennan, Bijan Teja, Michael Fehlings, Paul Ronksley, Alexander A Leung, Dany E Weisz, Harriet Ware, Mairead Whelan, David B Emerson, Rahul Arora, Niklas Bobrovitz

## Abstract

Systematic reviews (SRs) are the highest standard of evidence, shaping clinical practice guidelines, policy decisions, and research priorities. However, their labor-intensive nature, including an initial rigorous article screen by at least two investigators, delays access to reliable information synthesis. Here, we demonstrate that large language models (LLMs) with intentional prompting can match human screening performance. We introduce Framework Chain-of-Thought, a novel prompting approach that directs LLMs to systematically reason against predefined frameworks. We evaluated our prompts across ten SRs covering four common types of SR questions (i.e., prevalence, intervention benefits, diagnostic test accuracy, prognosis), achieving a mean accuracy of 93.6% (range: 83.3-99.6%) and sensitivity of 97.5% (89.7-100%) in full-text screening. Compared to experienced reviewers (mean accuracy 92.4% [76.8-97.8%], mean sensitivity 75.1% [44.1-100%]), our full-text prompt demonstrated significantly higher sensitivity in four reviews (p<0.05), significantly higher accuracy in one review (p<0.05), and comparable accuracy in two of five reviews (p>0.05). While traditional human screening for an SR of 7000 articles required 530 hours and $10,000 USD, our approach completed screening in one day for $430 USD. Our results establish that LLMs can perform SR screening with performance matching human experts, setting the foundation for end-to-end automated SRs.

## Main

Systematic reviews (SR) are rigorous forms of knowledge synthesis that involve the gathering, critical appraisal, and analysis of evidence. Recognized as the gold standard in evidence-based practice, SRs bolster decision-making across various domains including medicine, business, and agriculture, among others.^1^ SRs are resource-intensive, typically requiring one year and upwards of $100,000 to complete.^2,3^ These costs stem from the comprehensive processes of conducting detailed searches, screening articles, extracting data, analyzing findings, and report writing.^2–4^ The screening phase is particularly demanding and typically involves two investigators working independently, and in duplicate, to identify articles that meet predefined eligibility criteria.^1,5^ Investigators begin with an initial sensitive title and abstract screen, followed by an accurate screen of article full-texts.^1,5^ Despite a growing catalog of tools and resources,^6^ SR automation is elusive as existing tools only supplement human workflows and lack the performance required for independent decision making.^7^

The rise of large language models (LLMs), such as GPT, with advanced generative capabilities creates new horizons for streamlining and automating SR processes.^8,9^ MedPrompt,^10^ a collection of prompting techniques to optimize GPT4 performance on medical benchmarks, has demonstrated that generalist foundation models can surpass traditional model fine-tuning methods simply through better prompting. However, evaluations of LLMs in the medical domain have faced issues such as irreproducibility due to the use of web browser applications with hidden prompts, unrecorded model versions and settings, and the transient nature of chatbot histories. Furthermore, basic zero-shot prompting–where LLMs are only given task instructions without examples–likely underestimates model capabilities. Consequently, past zero-shot assessments of LLMs for SR screening have demonstrated low sensitivity/recall, failing to capture relevant studies and compromising their use for automation.^11–14^

Here, we present a comprehensive evaluation of LLM performance in SR screening and prompting innovations that enhance screening efficacy. First, we address a critical gap in SR automation and create *BenchSR*, a robust set of 10 SR datasets covering diverse medical question types and domains. We then introduce two prompting innovations: ‘Framework Chain of Thought (CoT),’ a novel prompting approach to guide models towards systematic reasoning against predetermined frameworks, and ‘Instruction Structure Optimized (ISO)’ prompting, designed to address LLM context-loss. We propose *Abstract ScreenPrompt* and *ISO-ScreenPrompt* for abstract and full-text screening, and demonstrate that their performance can match human reviewers. We evaluate our strategies across seven generalist LLMs (GPT3.5, GPT4-0125-preview, GPT4-Turbo-0409, GPT4o-0513, Gemini Pro, Mixtral-8×22, Mistral-Large) and demonstrate that our findings are model agnostic. Our work highlights the importance of applying rigorous prompting techniques to fully leverage the capabilities of LLMs, and serves as a guide for future researchers interested in performing LLM evaluations for SR screening and beyond. Collectively, our datasets and findings illustrate the feasibility of LLM-automated SR screening and lay the groundwork for fully automating the SR workflow.

## Results

### Datasets and BenchSR

We curated *BenchSR*, a collection of 10 SR datasets comprising over 170,000 articles, spanning four of six Oxford Center for Evidence-Based Medicine (CEBM) question types,^15,16^ and nine different clinical domains^17^ (Table 1, Fig. 1). This compilation includes SR metadata (inclusion/exclusion criteria, study objectives), and the complete set of labeled articles (included, excluded) for each review.

**Figure 1:**
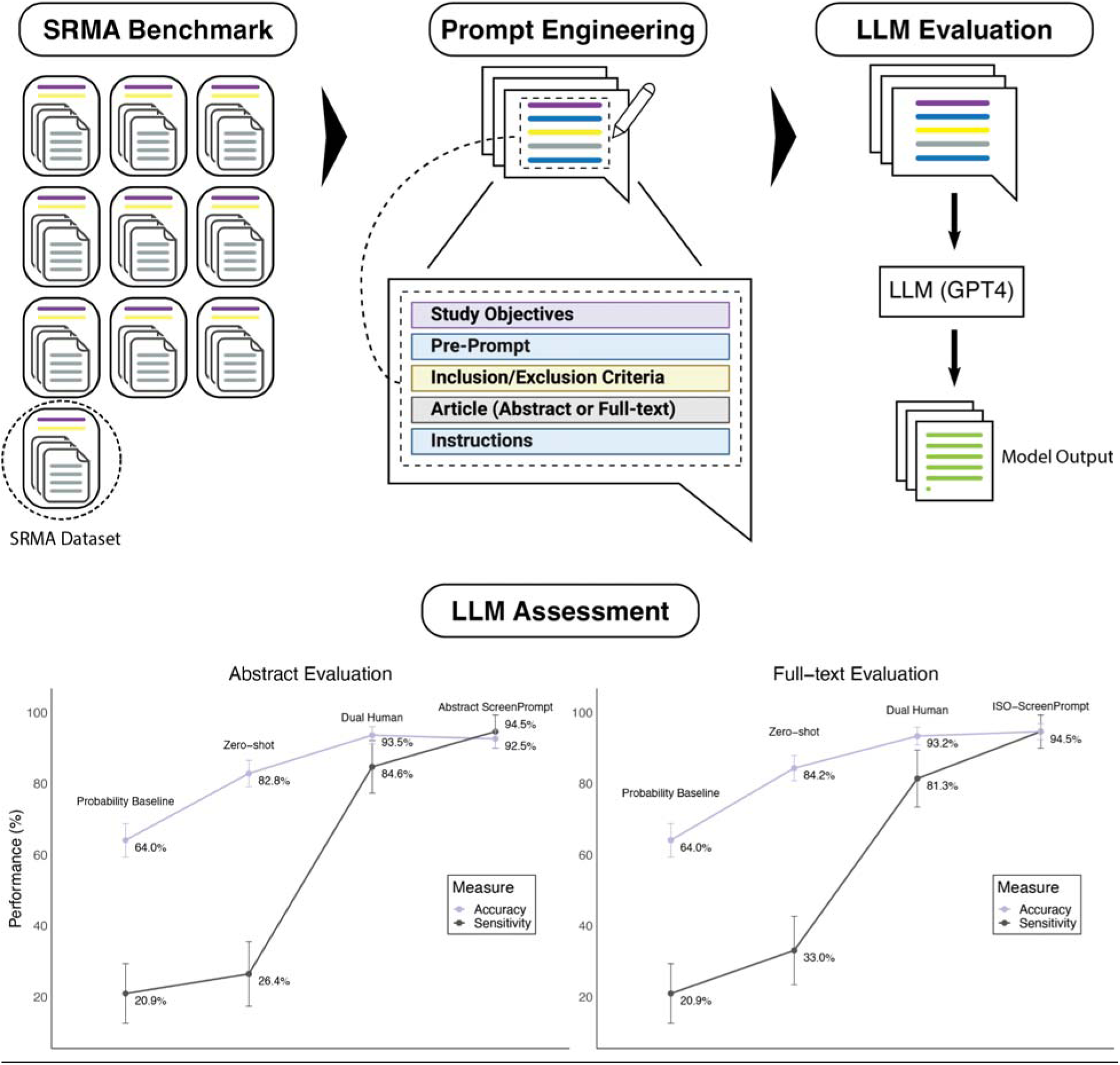
Infographic of study design. ScreenPrompt achieves SOTA performance for abstract and full-text SR screening. The LLM assessment component shows evaluation of the SeroTracker (ST) dataset (n=400) used in head-to-head human and LLM testing. Error bars represent 95% CIs for binomial proportions.

**Table 1:**
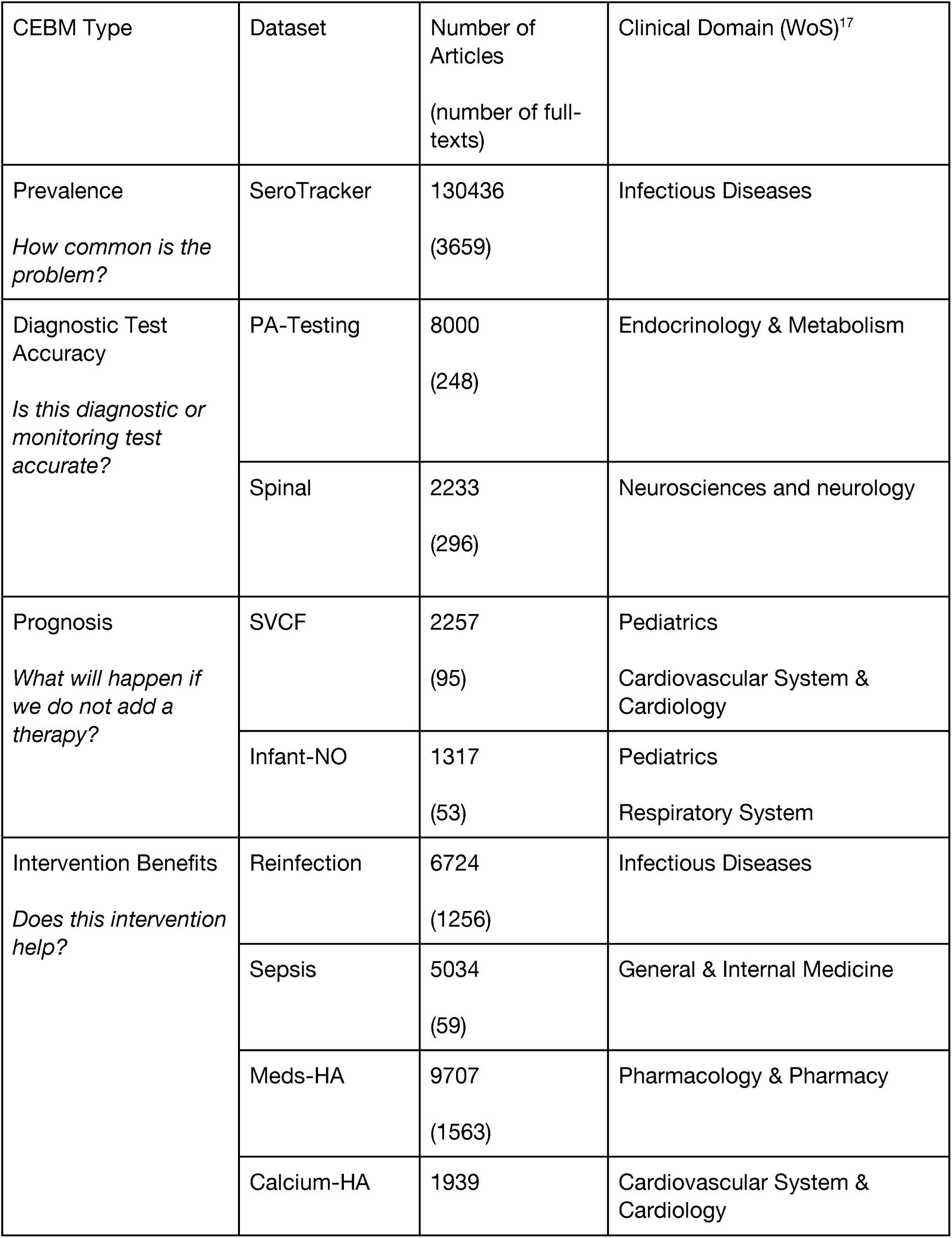
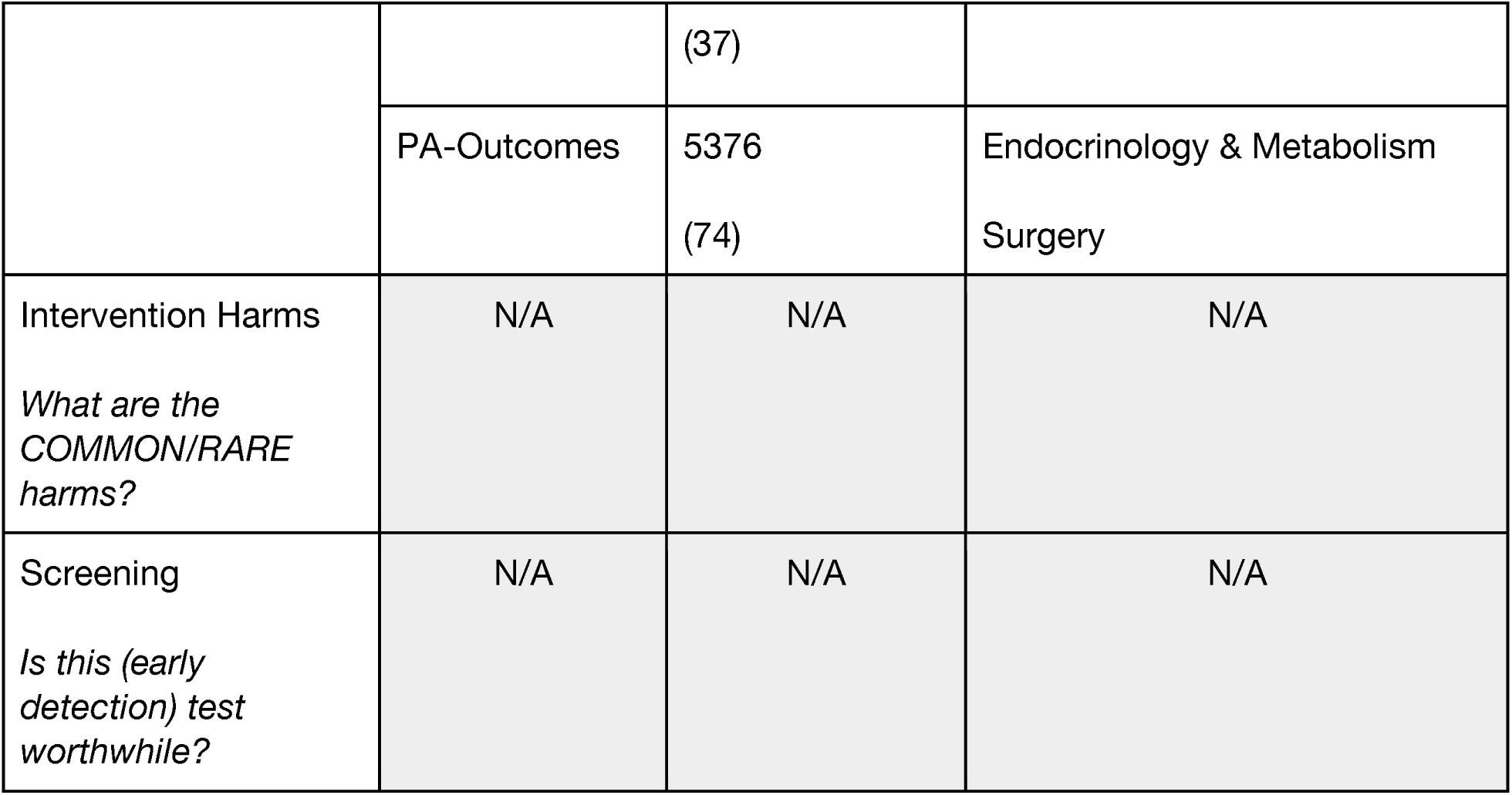
Descriptive Overview of *BenchSR* Datasets.

### Abstract Prompt Engineering

We evaluated the performance of three standard prompting methodologies on our training split dataset (SeroTracker [ST] train split; Methods, Testing Methodology) (Fig. 1, Fig. 2a, Table 2). Unless otherwise stated, we prompted the GPT4-0125-preview model with the ST train split. Initial tests with zero-shot prompting approaches (Methods Prompt Engineering, Table 2), adapted from Guo et al.,^11^ revealed suboptimal performance, with an accuracy of 65.0%, 30% sensitivity, and 100% specificity (Fig. 2b, Supplementary Table 1). These results were improved with random few-shot prompting (78% accuracy, 56% sensitivity, 100% specificity) (Fig. 2b, Supplementary Table 1). Rephrasing prompts to be more inclusive also improved sensitivity and performance (+3.3% accuracy, +6.5% sensitivity; Supplementary Table 2, Table 2), and we applied this consideration when prompting thereafter. Finally, Zero-shot CoT prompting^18^ was associated with notable improvements in accuracy (86.3% accuracy, 74.5% sensitivity, 98% specificity) (Fig. 2b, Supplementary Table 1), although model accuracy and sensitivity were relatively low for independent SR screening.

**Figure 2:**
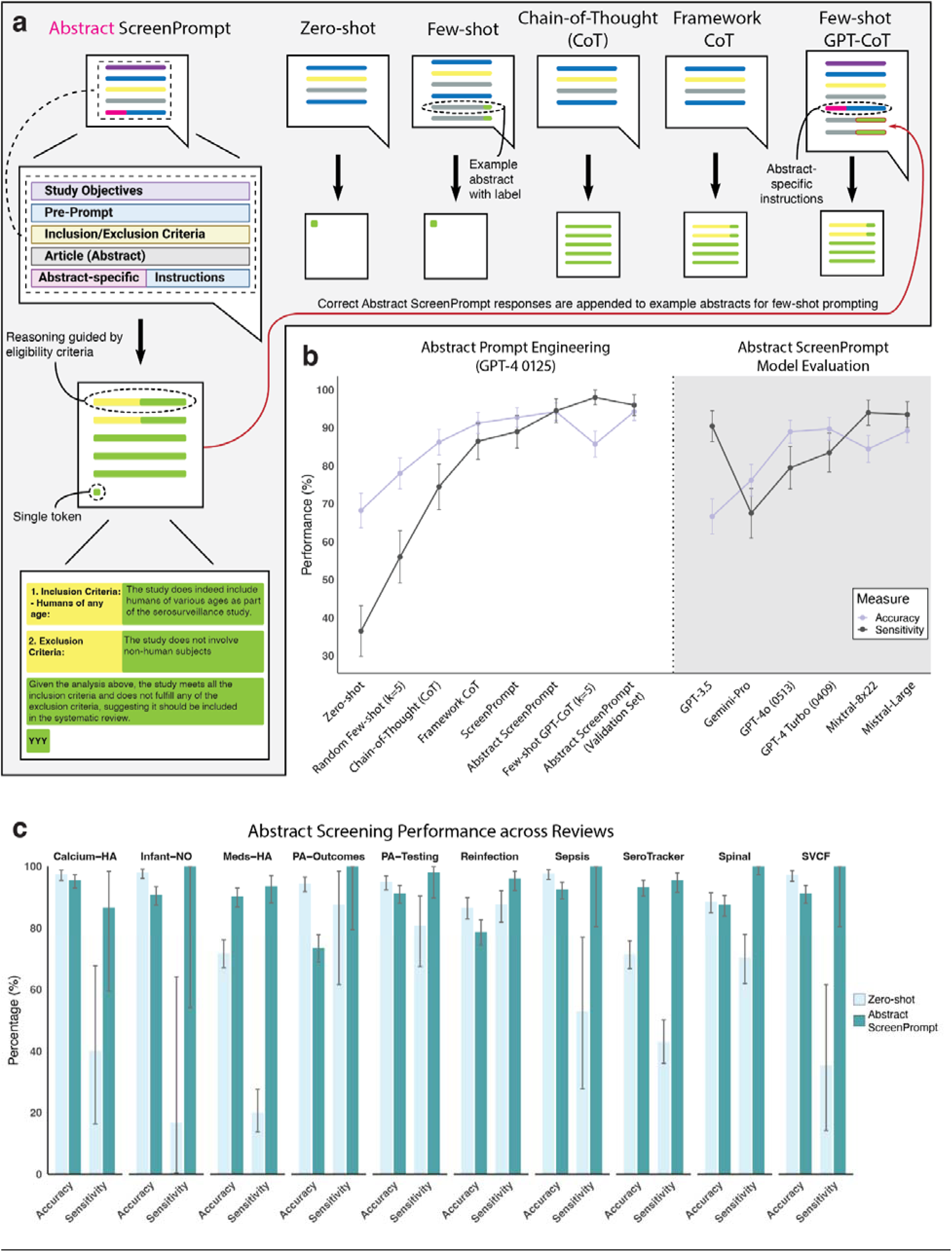
Abstract ScreenPrompt achieves SOTA performance for abstract screening, generalizing across studies. a) Diagram of different prompting strategies used for abstract screening, including Zero-shot, Few-shot, Chain-of-Thought (CoT), Framework CoT, and Few-shot GPT-CoT. The Abstract ScreenPrompt approach integrates study objectives, inclusion/exclusion criteria, and abstract-specific instructions to guide reasoning. b) Performance comparison of different abstract prompting methodologies on the SeroTracker training, showing accuracy and sensitivity. Abstract ScreenPrompt is evaluated within the ST training dataset (n=400) across GPT4-0125-preview, GPT-3-5, Gemini-Pro, GPT4-o-0513, GPT4-Turbo-0409, Mixtral-8×22, Mistral-Large. Abstract ScreenPrompt is also separately evaluated on the ST validation dataset (n=400). Error bars represent 95% CIs for binomial proportions. c) Barplot displaying generalizability of zero-shot Abstract ScreenPrompt sensitivity and accuracy across 10 different systematic review datasets from *BenchSR*. Error bars represent 95% CIs for exact proportions with the Clopper-Pearson method.

**Table 2:** Abstract and Full-text Screening Prompts.

| Prompting Technique | Prompt Structure. |
| --- | --- |
|  | Modules are enclosed in {} and were not inserted in the prompt.<sup>11</sup> |
| Zero-shot<br>*Adapted from Guo et al.<sup>11</sup> | {Pre-Prompt}<br>*You are a researcher rigorously screening titles and abstracts of scientific papers for inclusion or exclusion in a review paper. Use the criteria below to inform your decision. If any exclusion criteria are met or not all inclusion criteria are met, exclude the article. If all inclusion criteria are met, include the article.*<br>{Inclusion Criteria}<br>{Exclusion Criteria}<br>{Abstract in investigation}<br>{Instructions}<br>*Only type “YYY” for included articles or “XXX” for excluded articles to indicate your decision. Do not type anything else.*<br>{Model output} |
| Random few-shot<br>(k=10) | {Pre-Prompt}<br>*You are a researcher rigorously screening titles and abstracts of scientific papers for inclusion or exclusion in a review paper. Use the criteria below to inform your decision. If any exclusion criteria are met or not all inclusion criteria are met, exclude the article. If all inclusion criteria are met, include the article.*<br>**{Inclusion Criteria}**<br>**{Exclusion Criteria}**<br>{Example Include n= 5}<br>{Example Exclude n= 5}<br>{Abstract in investigation}<br>{Instructions}<br>*Only type “YYY” for included articles or “XXX” for excluded articles* |
|  | *to indicate your decision. Do not type anything else.*<br>{Model output} |
| Zero-shot CoT | {Pre-Prompt}<br>*You are a researcher rigorously screening titles and abstracts of scientific papers for inclusion or exclusion in a review paper. Use the criteria below to inform your decision. If any exclusion criteria are met or not all inclusion criteria are met, exclude the article. If all inclusion criteria are met, include the article.*<br>{Inclusion criteria}<br>{Exclusion Criteria}<br>{Abstract in investigation}<br>{Instructions}<br>***Let's think step by step for why an article should be included or excluded.*** We will conclude by outputting (on the very last line) 'XXX' if the paper warrants exclusion, or 'YYY' if inclusion is advised or uncertainty persists. We must output either 'XXX' or 'YYY'.<br>{Model output} |
| Zero-shot Framework CoT | {Pre-prompt}<br>*The following is an excerpt of two sets of criteria. A study is considered included if it meets all the inclusion criteria. If a study meets any of the exclusion criteria, it should be excluded. Here are the two sets of criteria:*<br>{Inclusion criteria}<br>{Exclusion Criteria}<br>{Abstract in investigation}<br>{Instructions}<br>*# Instructions*<br>***We now assess whether the paper should be included from*** |
|  | ***the systematic review by evaluating it against each and every predefined inclusion and exclusion criterion. First, we will reflect on how we will decide whether a paper should be included or excluded. Then, we will think step by step for each criteria, giving reasons for why they are met or not met.***<br>*We will conclude by outputting (on the very last line) 'XXX' if the paper warrants exclusion, or 'YYY' if inclusion is advised or uncertainty persists. We must output either 'XXX' or 'YYY'.*<br>{Model output} |
| ScreenPrompt | {Pre-prompt, including **objectives**}<br>***Our systematic review is governed by the following objectives: (i) describe the global prevalence of SARS-CoV-2 antibodies based on serosurveys; (ii) detect variations in seroprevalence arising from study design and geographic factors; (iii) identify populations at high risk for SARS-CoV-2 infection; and (iv) evaluate the extent to which surveillance based on detection of acute infection underestimates the spread of the pandemic.***<br>*The following is an excerpt of two sets of criteria. A study is considered included if it meets all the inclusion criteria. If a study meets any of the exclusion criteria, it should be excluded. Here are the two sets of criteria:*<br>{Inclusion criteria}<br>{Exclusion Criteria}<br>{Abstract in investigation}<br>{Instructions, including task considerations}<br>***# Instructions***<br>*We now assess whether the paper should be included from the systematic review by evaluating it against each and every predefined inclusion and exclusion criterion. First, we will reflect on how we will decide whether a paper should be included or excluded. Then, we will think step by step for each criteria, giving* |
|  | *reasons for why they are met or not met.*<br>*We will conclude by outputting (on the very last line) 'XXX' if the paper warrants exclusion, or 'YYY' if inclusion is advised or uncertainty persists. We must output either 'XXX' or 'YYY'.*<br>{Model output} |
| Abstract ScreenPrompt | {Pre-prompt, including objectives}<br>*Our systematic review is governed by the following objectives: (i) describe the global prevalence of SARS-CoV-2 antibodies based on serosurveys; (ii) detect variations in seroprevalence arising from study design and geographic factors; (iii) identify populations at high risk for SARS-CoV-2 infection; and (iv) evaluate the extent to which surveillance based on detection of acute infection underestimates the spread of the pandemic.*<br>*The following is an excerpt of two sets of criteria. A study is considered included if it meets all the inclusion criteria. If a study meets any of the exclusion criteria, it should be excluded. Here are the two sets of criteria:*<br>{Inclusion criteria}<br>{Exclusion Criteria}<br>{Abstract in investigation}<br>{Instructions, including abstract considerations}<br>*# Instructions*<br>*We now assess whether the paper should be included from the systematic review by evaluating it against each and every predefined inclusion and exclusion criterion. First, we will reflect on how we will decide whether a paper should be included or excluded. Then, we will think step by step for each criteria, giving reasons for why they are met or not met.*<br>***Studies that may not fully align with the primary focus of our inclusion criteria but provide data or insights potentially relevant to our review deserve thoughtful consideration. Given*** |
|  | ***the nature of abstracts as concise summaries of comprehensive research, some degree of interpretation is necessary.***<br>*Our aim should be to inclusively screen abstracts, ensuring broad coverage of pertinent studies while filtering out those that are clearly irrelevant. We will conclude by outputting (on the very last line) 'XXX' if the paper warrants exclusion, or 'YYY' if inclusion is advised or uncertainty persists. We must output either 'XXX' or 'YYY'.*<br>{Model output} |
| Few-shot GPT-CoT<br>Abstract ScreenPrompt<br>(k=10) | {Pre-prompt, including objectives}<br>*Our systematic review is governed by the following objectives: (i) describe the global prevalence of SARS-CoV-2 antibodies based on serosurveys; (ii) detect variations in seroprevalence arising from study design and geographic factors; (iii) identify populations at high risk for SARS-CoV-2 infection; and (iv) evaluate the extent to which surveillance based on detection of acute infection underestimates the spread of the pandemic.*<br>*The following is an excerpt of two sets of criteria. A study is considered included if it meets all the inclusion criteria. If a study meets any of the exclusion criteria, it should be excluded. Here are the two sets of criteria:*<br>{Inclusion criteria}<br>{Exclusion Criteria}<br>**{GPT-CoT Include n= 5}**<br>**{GPT-CoT Exclude n= 5}**<br>{Abstract in investigation}<br>{Instructions, including abstract considerations}<br>*# Instructions*<br>*We now assess whether the paper should be included from the* |
|  | *systematic review by evaluating it against each and every predefined inclusion and exclusion criterion. First, we will reflect on how we will decide whether a paper should be included or excluded. Then, we will think step by step for each criteria, giving reasons for why they are met or not met.*<br>*Studies that may not fully align with the primary focus of our inclusion criteria but provide data or insights potentially relevant to our review deserve thoughtful consideration. Given the nature of abstracts as concise summaries of comprehensive research, some degree of interpretation is necessary.*<br>*Our aim should be to inclusively screen abstracts, ensuring broad coverage of pertinent studies while filtering out those that are clearly irrelevant. We will conclude by outputting (on the very last line) 'XXX' if the paper warrants exclusion, or 'YYY' if inclusion is advised or uncertainty persists. We must output either 'XXX' or 'YYY'.*<br>{Model output} |
| Full-text |  |
| Numbered<br>ScreenPrompt: | {Pre-prompt, including objectives}<br>*Our systematic review is governed by the following objectives: (i) describe the global prevalence of SARS-CoV-2 antibodies based on serosurveys; (ii) detect variations in seroprevalence arising from study design and geographic factors; (iii) identify populations at high risk for SARS-CoV-2 infection; and (iv) evaluate the extent to which surveillance based on detection of acute infection underestimates the spread of the pandemic.*<br>*The following is an excerpt of two sets of criteria. A study is considered included if it meets all the inclusion criteria. If a study meets any of the exclusion criteria, it should be excluded. Here are the two sets of criteria:*<br>{**Numbered** Inclusion criteria}<br>**1. Humans of any age**<br>... |
|  | **11.** Reports the locations at which the study took places such that they could be categorized as neighbourhood, city, state/province/territory, or country<br>{**Numbered** Exclusion Criteria}<br>**1.** Non-human (e.g., in silico, animal, in vitro)<br>...<br>**10.** Does not report the location at which the study took place<br>{Abstract in investigation}<br>{Instructions}<br># Instructions<br>We now assess whether the paper should be included from the systematic review by evaluating it against each and every predefined inclusion and exclusion criterion. First, we will reflect on how we will decide whether a paper should be included or excluded. Then, we will think step by step for each criteria, giving reasons for why they are met or not met.<br>We will conclude by outputting (on the very last line) 'XXX' if the paper warrants exclusion, or 'YYY' if inclusion is advised or uncertainty persists. We must output either 'XXX' or 'YYY'.<br>{Model output} |
| ScreenPrompt (Init): | {Pre-prompt, including objectives}<br>Our systematic review is governed by the following objectives: (i) describe the global prevalence of SARS-CoV-2 antibodies based on serosurveys; (ii) detect variations in seroprevalence arising from study design and geographic factors; (iii) identify populations at high risk for SARS-CoV-2 infection; and (iv) evaluate the extent to which surveillance based on detection of acute infection underestimates the spread of the pandemic.<br>The following is an excerpt of two sets of criteria. A study is considered included if it meets all the inclusion criteria. If a study meets any of the exclusion criteria, it should be excluded. Here are the two sets of criteria: |
|  | {Inclusion criteria}<br>{Exclusion Criteria}<br>**{Instructions}**<br>*# Instructions*<br>*We now assess whether the paper should be included from the systematic review by evaluating it against each and every predefined inclusion and exclusion criterion. First, we will reflect on how we will decide whether a paper should be included or excluded. Then, we will think step by step for each criteria, giving reasons for why they are met or not met.*<br>*We will conclude by outputting (on the very last line) 'XXX' if the paper warrants exclusion, or 'YYY' if inclusion is advised or uncertainty persists. We must output either 'XXX' or 'YYY'.*<br>{Abstract in investigation}<br>{Model output} |
| ScreenPrompt (Fin): | {Abstract in investigation}<br>**{Pre-prompt, including objectives}**<br>*Our systematic review is governed by the following objectives: (i) describe the global prevalence of SARS-CoV-2 antibodies based on serosurveys; (ii) detect variations in seroprevalence arising from study design and geographic factors; (iii) identify populations at high risk for SARS-CoV-2 infection; and (iv) evaluate the extent to which surveillance based on detection of acute infection underestimates the spread of the pandemic.*<br>*The following is an excerpt of two sets of criteria. A study is considered included if it meets all the inclusion criteria. If a study meets any of the exclusion criteria, it should be excluded. Here are the two sets of criteria:*<br>**{Inclusion criteria}**<br>**{Exclusion Criteria}** |
|  | **{Instructions}**<br>*# Instructions*<br>*We now assess whether the paper should be included from the systematic review by evaluating it against each and every predefined inclusion and exclusion criterion. First, we will reflect on how we will decide whether a paper should be included or excluded. Then, we will think step by step for each criteria, giving reasons for why they are met or not met.*<br>*We will conclude by outputting (on the very last line) 'XXX' if the paper warrants exclusion, or 'YYY' if inclusion is advised or uncertainty persists. We must output either 'XXX' or 'YYY'.*<br>{Model output} |
| ScreenPrompt (Init + Fin-Instructions): | {Pre-prompt, including objectives}<br>*Our systematic review is governed by the following objectives: (i) describe the global prevalence of SARS-CoV-2 antibodies based on serosurveys; (ii) detect variations in seroprevalence arising from study design and geographic factors; (iii) identify populations at high risk for SARS-CoV-2 infection; and (iv) evaluate the extent to which surveillance based on detection of acute infection underestimates the spread of the pandemic.*<br>*The following is an excerpt of two sets of criteria. A study is considered included if it meets all the inclusion criteria. If a study meets any of the exclusion criteria, it should be excluded. Here are the two sets of criteria:*<br>{Inclusion criteria}<br>{Exclusion Criteria}<br>**{Instructions}**<br>*# Instructions*<br>*We now assess whether the paper should be included from the systematic review by evaluating it against each and every predefined inclusion and exclusion criterion. First, we will reflect on how we will decide whether a paper should be included or* |
|  | *excluded. Then, we will think step by step for each criteria, giving reasons for why they are met or not met.*<br>*We will conclude by outputting (on the very last line) 'XXX' if the paper warrants exclusion, or 'YYY' if inclusion is advised or uncertainty persists. We must output either 'XXX' or 'YYY'.*<br>{Abstract in investigation}<br>**{Instructions}**<br>*# Instructions*<br>*We now assess whether the paper should be included from the systematic review by evaluating it against each and every predefined inclusion and exclusion criterion. First, we will reflect on how we will decide whether a paper should be included or excluded. Then, we will think step by step for each criteria, giving reasons for why they are met or not met.*<br>*We will conclude by outputting (on the very last line) 'XXX' if the paper warrants exclusion, or 'YYY' if inclusion is advised or uncertainty persists. We must output either 'XXX' or 'YYY'.*<br>{Model output} |
| ScreenPrompt<br>(Init + Fin): | {Pre-prompt, including objectives}<br>*Our systematic review is governed by the following objectives: (i) describe the global prevalence of SARS-CoV-2 antibodies based on serosurveys; (ii) detect variations in seroprevalence arising from study design and geographic factors; (iii) identify populations at high risk for SARS-CoV-2 infection; and (iv) evaluate the extent to which surveillance based on detection of acute infection underestimates the spread of the pandemic.*<br>*The following is an excerpt of two sets of criteria. A study is considered included if it meets all the inclusion criteria. If a study meets any of the exclusion criteria, it should be excluded. Here are the two sets of criteria:*<br>{Inclusion criteria} |
|  | {Exclusion Criteria}<br>**{Instructions}**<br>*# Instructions*<br>*We now assess whether the paper should be included from the systematic review by evaluating it against each and every predefined inclusion and exclusion criterion. First, we will reflect on how we will decide whether a paper should be included or excluded. Then, we will think step by step for each criteria, giving reasons for why they are met or not met.*<br>*We will conclude by outputting (on the very last line) 'XXX' if the paper warrants exclusion, or 'YYY' if inclusion is advised or uncertainty persists. We must output either 'XXX' or 'YYY'.*<br>{Abstract in investigation}<br>**{Pre-prompt, including objectives}**<br>*Our systematic review is governed by the following objectives: (i) describe the global prevalence of SARS-CoV-2 antibodies based on serosurveys; (ii) detect variations in seroprevalence arising from study design and geographic factors; (iii) identify populations at high risk for SARS-CoV-2 infection; and (iv) evaluate the extent to which surveillance based on detection of acute infection underestimates the spread of the pandemic.*<br>*The following is an excerpt of two sets of criteria. A study is considered included if it meets all the inclusion criteria. If a study meets any of the exclusion criteria, it should be excluded. Here are the two sets of criteria:*<br>**{Inclusion criteria}**<br>**{Exclusion Criteria}**<br>**{Instructions}**<br>*# Instructions*<br>*We now assess whether the paper should be included from the systematic review by evaluating it against each and every* |
|  | *predefined inclusion and exclusion criterion. First, we will reflect on how we will decide whether a paper should be included or excluded. Then, we will think step by step for each criteria, giving reasons for why they are met or not met.*<br>*We will conclude by outputting (on the very last line) 'XXX' if the paper warrants exclusion, or 'YYY' if inclusion is advised or uncertainty persists. We must output either 'XXX' or 'YYY'.* |
| ISO-ScreenPrompt: | *{Pre-prompt, including objectives}*<br>*Our systematic review is governed by the following objectives: (i) describe the global prevalence of SARS-CoV-2 antibodies based on serosurveys; (ii) detect variations in seroprevalence arising from study design and geographic factors; (iii) identify populations at high risk for SARS-CoV-2 infection; and (iv) evaluate the extent to which surveillance based on detection of acute infection underestimates the spread of the pandemic.*<br>*The following is an excerpt of two sets of criteria. A study is considered included if it meets all the inclusion criteria. If a study meets any of the exclusion criteria, it should be excluded. Here are the two sets of criteria:*<br>*{**Numbered** Inclusion criteria}*<br>*{**Numbered** Exclusion Criteria}*<br>*{Instructions}*<br>*# Instructions*<br>*We now assess whether the paper should be included from the systematic review by evaluating it against each and every predefined inclusion and exclusion criterion. First, we will reflect on how we will decide whether a paper should be included or excluded. Then, we will think step by step for each criteria, giving reasons for why they are met or not met.*<br>*We will conclude by outputting (on the very last line) 'XXX' if the paper warrants exclusion, or 'YYY' if inclusion is advised or uncertainty persists. We must output either 'XXX' or 'YYY'.* |
|  | {Abstract in investigation}<br>**{Pre-prompt, including objectives}**<br>*Our systematic review is governed by the following objectives: (i) describe the global prevalence of SARS-CoV-2 antibodies based on serosurveys; (ii) detect variations in seroprevalence arising from study design and geographic factors; (iii) identify populations at high risk for SARS-CoV-2 infection; and (iv) evaluate the extent to which surveillance based on detection of acute infection underestimates the spread of the pandemic.*<br>*The following is an excerpt of two sets of criteria. A study is considered included if it meets all the inclusion criteria. If a study meets any of the exclusion criteria, it should be excluded. Here are the two sets of criteria:*<br>**{Numbered Inclusion criteria}**<br>**{Numbered Exclusion Criteria}**<br>**{Instructions}**<br>*# Instructions*<br>*We now assess whether the paper should be included from the systematic review by evaluating it against each and every predefined inclusion and exclusion criterion. First, we will reflect on how we will decide whether a paper should be included or excluded. Then, we will think step by step for each criteria, giving reasons for why they are met or not met.*<br>*We will conclude by outputting (on the very last line) 'XXX' if the paper warrants exclusion, or 'YYY' if inclusion is advised or uncertainty persists. We must output either 'XXX' or 'YYY'.*<br>{Model output} |

We analyzed zero-shot CoT outputs and identified inconsistencies in the generated reasoning structures for article inclusion (Supplementary Table 3). This insight inspired us to experiment with directing the LLM to articulate its reasoning according to the predefined inclusion and exclusion criteria. This novel zero-shot prompting approach was termed ‘Framework CoT’ and reported notable performance improvements (91.3% accuracy, 86.5% sensitivity, 96% specificity) (Fig. 2b, Supplementary Table 1, Methods Abstract Prompt Engineering).

Further analysis of incorrect zero-shot framework CoT model outputs revealed that the model generated incorrect inferences of review study objectives based on the inclusion and exclusion criteria (Supplementary Table 3). To counteract this, we inserted study objectives directly from the SR protocol or manuscript, and defined our prompting strategy as ‘ScreenPrompt’ (Table 2). ScreenPrompt was further refined by adding context about the limitations of abstract content and importance of article inclusion (‘+ abstract’, Supplementary Fig. 1a, Supplementary Table 4). This final abstract-specific prompting strategy was referred to as ‘Abstract ScreenPrompt’ (Table 2, Supplementary Fig. 1a, Supplementary Table 4).

We found that our Abstract ScreenPrompt prompting approach had the best performance on our ST training dataset (94.3% accuracy, 94.5% sensitivity, 94% specificity) (Fig. 2b, Supplementary Table 1). To check for risk of overfitting, we evaluated our Abstract ScreenPrompt approach on our ST validation dataset, and found comparable performance (94.3% accuracy, 96% sensitivity, 92.5% specificity) (Fig. 2b, Supplementary Table 1), confirming that our prompt engineering process did not overfit the training dataset.

While few-shot prompting (adding additional labeled examples) is traditionally believed to enhance LLM performance,^19^ our balanced few-shot GPT-CoT Abstract ScreenPrompt prompt was associated with decreased performance (85.8% accuracy, 98.0% sensitivity, 73.5% specificity). We hypothesized that we could modulate model specificity and sensitivity by adjusting the ratio of included and excluded few-shot examples (Supplementary Fig. 1b, Supplementary Table 5). Surprisingly, we found that our sensitivity analysis with varied ratios (9:1 inclusion- or exclusion-favored GPT-CoT Abstract ScreenPrompt few-shot examples) did not notably alter performance for inclusion-favored few-shot prompting (inclusion-favored: 85% accuracy, 98% sensitivity, 72% specificity), and was associated with a decrease in performance for exclusion-favored few-shot prompting (exclusion-favored: 83.8% accuracy, 98.5% sensitivity, 69% specificity).

### Abstract Screening Performance across LLMs

We conducted a comparative analysis across GPT4-0125-preview, GPT4-Turbo-0409, GPT4o-0513, GPT-3.5, Gemini Pro, Mixtral-8×22, and Mistral-Large LLM models to assess performance differences. We utilized the same ST training dataset and Abstract ScreenPrompt prompting strategy (Fig. 2b, Supplementary Table 6). Our cross-model evaluation found that the GPT4-0125-preview model was associated with the greatest performance (94.3% accuracy, 94.5% sensitivity, 94% specificity), but also had the highest cost ($12.54, 400 abstracts). In contrast, the GPT3.5 model was associated with the lowest overall performance, but had high sensitivity (66.9% accuracy, 90.5% sensitivity, 43% specificity). The Gemini Pro model had relatively low performance (77.5% accuracy, 67.5% sensitivity, 84.8% specificity), but was free to run. We found a moderate drop in accuracy and sensitivity with the newer GPT4-Turbo-0409 (89.8% accuracy, 83.5% sensitivity, 96% specificity) and GPT4o-0513 models (89.0% accuracy, 79.5% sensitivity, 98.5% specificity). To confirm that our prompting innovations were model agnostic, we compared Zero-Shot and Abstract ScreenPrompt performance in our lowest performing (GPT3.5) and next best performing (GPT4-Turbo-0409) models (Supplementary Table 7). We found that Abstract ScreenPrompt was associated with improvements in accuracy in both models, with the greatest sensitivity gains in GPT4-Turbo-0409 (+70% sensitivity).

### Generalizability of Abstract ScreenPrompt

We hypothesized that our Abstract ScreenPrompt could be readily adapted for abstract screening beyond the ST dataset. To test this, we applied our prompt to 10 distinct SR datasets within our *BenchSR* (Fig. 2c, Supplementary Table 8), using original study objectives and eligibility criteria for each dataset without modification. Our Abstract ScreenPrompt demonstrated high sensitivity across reviews (97.0% [range: 86.7-100.0%]), in contrast to zero-shot prompting (53.4% mean sensitivity [16.7-87.6%]). Both Abstract ScreenPrompt (88.4% [73.5-95.5%]) and zero-shot prompting (89.8% [71.5-98.0%]) produced comparable accuracy.

Our previous evaluations were performed with a single sampled generation. However, due to the stochasticity of LLM generations, the model may produce varying reasoning traces for a given prompt. Therefore, we applied 11-vote self-consistency (SC)^20^ to Abstract ScreenPrompt on the ST test dataset, and the Reinfection dataset, which had the lowest specificity (Supplementary Fig. 1c, Supplementary Table 9). We found that Abstract ScreenPrompt-SC was associated with an overall gain in performance in both datasets across all metrics (ST: +2.8% accuracy, +2% sensitivity, +3.5% specificity; Reinfection: +1.9% accuracy, +1.7% sensitivity, +1.8% specificity). Additionally, we could modulate sensitivity and specificity by setting different self-consistency thresholds (number of votes needed) for article inclusion/exclusion (Supplementary Fig. 1d, Methods Prompt Engineering). Collectively, these findings suggest that our Abstract ScreenPrompt prompting strategy is readily generalizable and can perform well across different SR contexts.

### Full-text Prompt Engineering

Building on the performance of Abstract ScreenPrompt, we evaluated the capabilities of the LLMs for full-text article screening (Fig. 3a, Supplementary Table 10-11, Table 2). Unless otherwise stated, we prompted the GPT4-0125-preview model with the full-text ST training dataset. Our original Abstract ScreenPrompt approach, tailored for inclusion, demonstrated high sensitivity (84.2% accuracy, 99% sensitivity, 69.2% specificity) (Fig. 3a). However, we sought to prioritize accuracy in downstream optimization steps.

**Figure 3:**
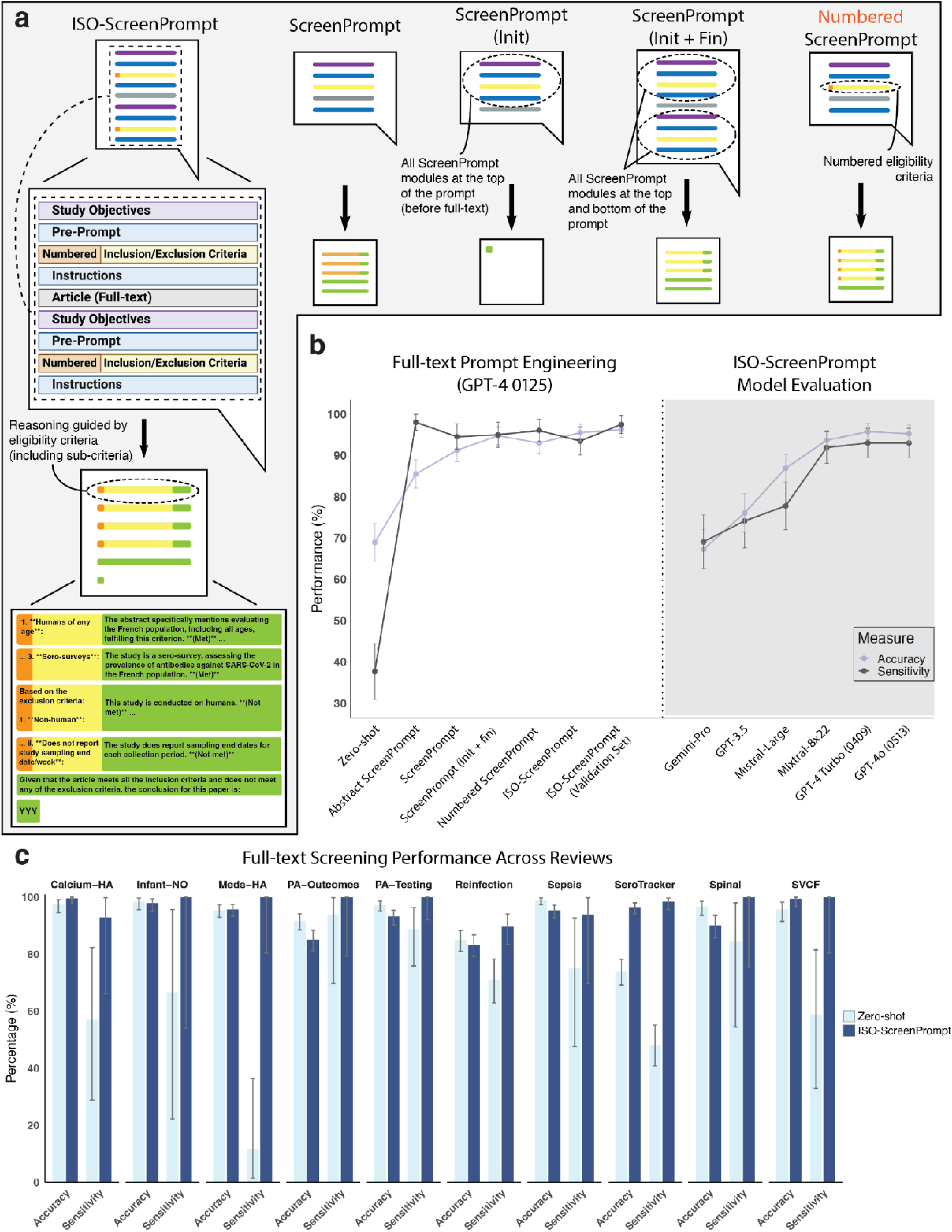
ISO-ScreenPrompt achieves SOTA performance for full-text screening, generalizing across studies. a) Diagram illustrating the various prompting strategies used for full-text screening, including Zero-shot, Few-shot, Chain-of-Thought (CoT), Framework CoT, and Few-shot GPT-CoT. The ISO-ScreenPrompt approach incorporates repeated prompt modules (i.e., objectives, pre-prompt, eligibility criteria, instructions) and numbering to enhance performance. b) Performance comparison of different full-text prompting methodologies on the SeroTracker (ST) training dataset (n=400), showing accuracy and sensitivity. ISO-ScreenPrompt is evaluated across multiple models with the ST training dataset: GPT4-0125-preview, GPT-3.5, Gemini-Pro, GPT4o-0513, GPT4-Turbo-0409, Mixtral-8×22, and Mistral-Large. ISO-ScreenPrompt is also separately evaluated on the ST validation dataset (n=400). Error bars represent 95% CIs for binomial proportions. c) Barplot displaying the generalizability of the zero-shot ISO-ScreenPrompt sensitivity and accuracy across 10 different systematic review datasets from *BenchSR*. Error bars represent 95% CIs for exact proportions with the Clopper-Pearson method.

While abstracts are concise summaries of study findings, full-text articles can span thousands of words. Therefore we hypothesized that adjustments to our prompt structure—without making semantic changes to prompt content—could aid full-text screening. This hypothesis was based on the ‘lost-in-the-middle’ phenomenon,^21,22^ which refers to the variable information retrieval rates of LLMs within lengthy texts. Our experiments positioning the pre-prompt, inclusion criteria, and instruction prompt modules before the full-text article was associated with modest performance gains (Init: 92.5% accuracy, 94.5% sensitivity, 90.5% specificity), while placing them after the full-text articles reduced performance (Fin: 82.2% accuracy, 98% sensitivity, 66.3% specificity) (Supplementary Fig. 2a, Supplementary Table 11). With ‘init’ prompting, the model occasionally provided a single token decision with no additional reasoning (Supplementary Table 12). Appending additional instructions at the end of the prompt helped mitigate this inconsistency (‘Init + Fin-Instructions’, Supplementary Table 12). Further improvements were observed with our Framework CoT (init + fin) prompt, where we appended the complete set of prompt modules before and after the full-text article (94.8% accuracy, 95% sensitivity, and 94.5% specificity) (Fig. 3a, Supplementary Fig. 2a, Supplementary Table 11). Applying the same prompting strategy to abstract screening did not notably improve results, (Supplementary Fig. 2b, Supplementary Table 13) possibly because abstracts are too short to manifest the lost-in-the-middle phenomenon.

When analyzing the model outputs, we also found that LLM responses occasionally lacked sufficiently granular reasoning against each inclusion and exclusion criterion (Supplementary Table 12). We refined our Framework CoT prompt to elicit detailed CoT reasoning for each individual sub-criterion by numbering them, while preserving the original criteria content. We term this revised prompt as ‘Numbered Framework CoT’ and similarly found improved performance (95.2% accuracy, 98% sensitivity, 92.4% specificity) (Fig. 3b, Supplementary Table 10).

Merging ‘Numbered Framework CoT’ with our optimal prompt structure (init + fin) resulted in a prompting strategy dubbed ‘*Instruction-Structure-Optimized (ISO) ScreenPrompt*’. ISO-ScreenPrompt exhibited the best overall performance on our training dataset (95.5% accuracy, 94% sensitivity, 98% specificity) (Fig. 3b, Supplementary Table 10). Evaluation on the separate ST validation dataset confirmed the robustness of our approach, showing comparable performance (96.3% accuracy, 97.5% sensitivity, 95% specificity) and confirmed that we did not overfit our training dataset.

### Full-text Screening Performance across LLMs

Consistent with our abstract findings, our comparative analysis between LLM models revealed that Gemini Pro and GPT3.5 models had poor performance across all metrics (Supplementary Table 14). Interestingly, Mixtral-8×22 (93.7% accuracy, 91.9% sensitivity, 95.4% specificity) outperformed Mistral-Large (86.9% accuracy, 77.8% sensitivity, 96% specificity). GPT4-0125-preview, GPT4-Turbo-0409, and GPT4o-0513 models all had similar performance (95.3-95.8% accuracy, 93.0-93.5% sensitivity, 97.5-98.5% specificity). We then compared Zero-Shot and ISO-ScreenPrompt performance in our lowest performing (GPT3.5) and best performing model (GPT4-Turbo-0409) to assess model agnosticism (Supplementary Table 16). We found that ISO-ScreenPrompt was associated with improvements in accuracy across all models, with the greatest sensitivity gains in GPT4-Turbo-0409 (+79.5% sensitivity).

### Generalizability of ISO-ScreenPrompt

We assessed the generalizability of our ISO-ScreenPrompt approach for full-text screening across datasets from *BenchSR* (Fig. 3c, Supplementary Table 16). We found that ISO-ScreenPrompt demonstrated high sensitivity across reviews, with 97.5% (89.7-100.0%) mean sensitivity, in contrast to zero-shot prompting (65.6% [11.8-93.8 %]). ISO-ScreenPrompt (93.6% [83.2-99.6%]) and zero-shot prompting (93.1% [73.9-99.0%]) demonstrated comparable accuracy, but ISO-ScreenPrompt had slightly higher accuracy with lower variance. When compared to Abstract ScreenPrompt, ISO-ScreenPrompt had a modest improvement in accuracy while maintaining sensitivity.

We applied self-consistency (SC) to ISO-ScreenPrompt on the ST test dataset (Methods, Testing Methodology) and the Reinfection dataset (Supplementary Fig. 2c, Supplementary Table 17). We found that ISO-ScreenPrompt-SC was associated with an overall gain in performance in both datasets across all metrics, except sensitivity in ST (ST: +1% accuracy, -1% sensitivity, +3% specificity; Reinfection: +4.3% accuracy, +2.8% sensitivity, +5.1% specificity). We could also modulate sensitivity and specificity by setting different self-consistency thresholds for article inclusion/exclusion (Supplementary Fig. 2d). Collectively, these findings suggest that our ISO-ScreenPrompt prompting strategy is generalizable and can perform well across different SR contexts.

### Real World Implementation Assessment and Benefit

Dual human screening is considered the gold-standard approach for SR screening workflows.^23,24^ In this process, at least two reviewers independently perform title and abstract screening, resolving any discrepancies through consensus or decision by a third reviewer (Fig. 4a). Relevant abstracts are then moved to a full-text screen, and the process is repeated to culminate a ‘full dual screen’ that identifies the final set of articles for inclusion (Fig. 4a).

**Figure 4:**
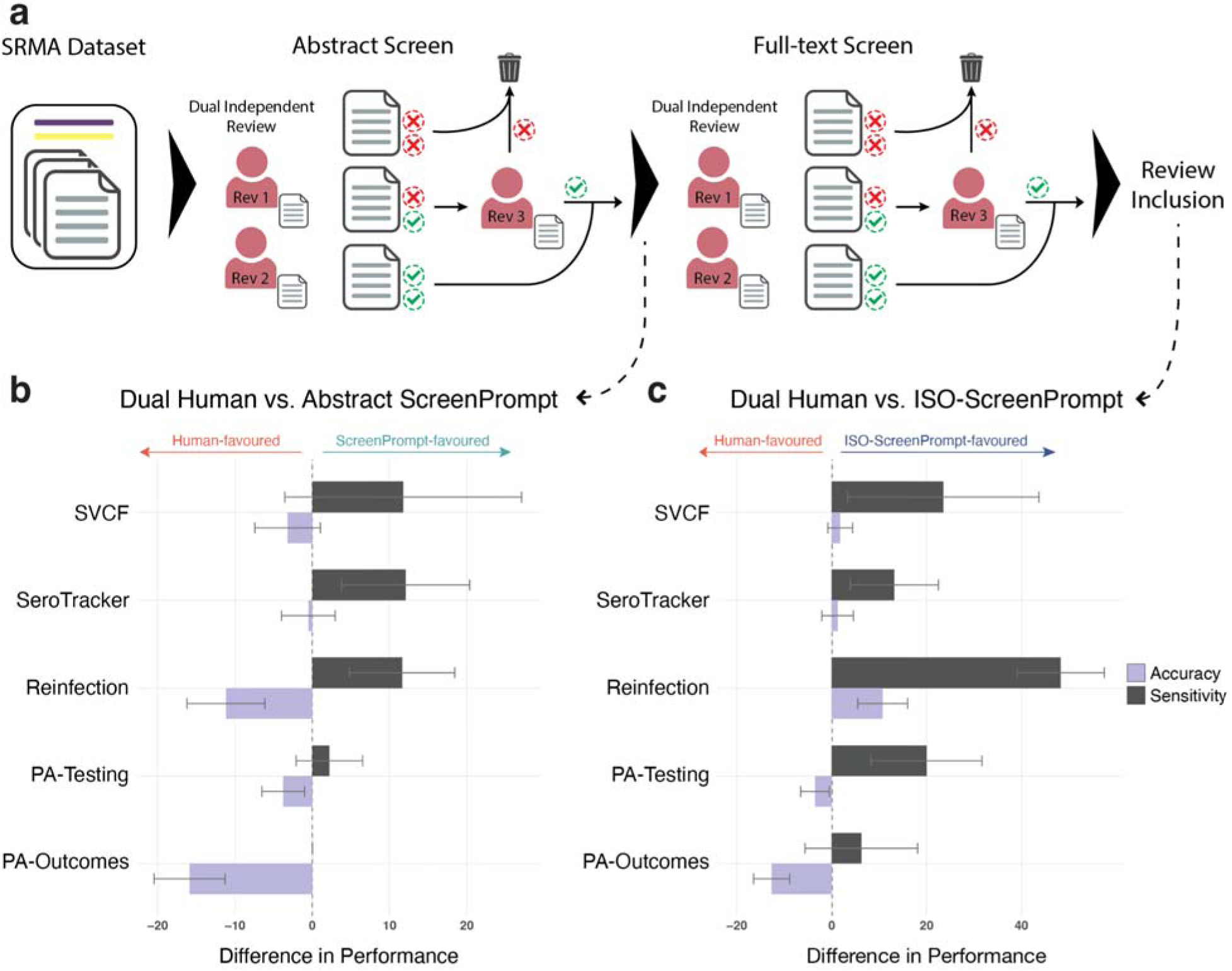
ScreenPrompt and ISO-ScreenPrompt perform comparably, or better than dual human review a) Overview of the dual human review process for systematic reviews, illustrating dual independent review stages for both abstract and full-text screening, leading to final review inclusion. Conflicts between reviewers are resolved by a third reviewer. b) Difference in performance between Dual Human Review and Abstract ScreenPrompt across five systematic reviews (SVCF n=167, SeroTracker n=400, Reinfection n=400, PA-Testing n=400, PA-Outcomes n=400). The barplot shows differences in accuracy and sensitivity, with human-favored differences as negative (left) and ScreenPrompt-favored differences as positive (right). Error bars represent 95% CIs for the difference in binomial proportions. c) Difference in performance between Dual Human Review and ISO-ScreenPrompt across five systematic reviews (SVCF, SeroTracker, Reinfection, PA-Testing, PA-Outcomes). The barplot shows differences in accuracy and sensitivity, with human-favored differences as negative (left) and ISO-ScreenPrompt-favored differences as positive (right). Error bars represent 95% CIs for the difference in binomial proportions.

To evaluate the real-world applicability of our approach, we performed head-to-head comparisons between our Abstract ScreenPrompt and ISO-ScreenPrompt with the traditional gold-standard dual human screening workflow (Fig. 4a). We selected reviews using stratified probability sampling for each Oxford CEBM question type, with two spots for intervention benefit due to their higher prevalence in the dataset. The article inclusion/exclusion labels set by the original study authors were used as the gold-standard.

We recruited a team of four researchers with previous SR experience and performed a ‘calibration’ exercise to evaluate their baseline screening performance, finding acceptable performance (Supplementary Table 18, Methods Head-to-Head Comparisons). Reviewers then screened the randomly selected reviews according to the standard dual screening workflow. We found that human reviewers performed well with dual abstract screening (mean accuracy 94.6% [89.5-97.8%]), mean sensitivity 90.9% [84.1-100%]), but performance dropped with full dual screening (mean accuracy 92.4% [76.8-97.8%], mean sensitivity 75.1% [44.1-100%]) (Supplementary Table 19-20).

Head-to-head comparisons between Abstract ScreenPrompt and dual abstract screening revealed comparable sensitivity across three reviews (difference in binomial proportions, two-sided p>0.05). However, human reviewers exhibited significantly higher accuracy in three reviews (p<0.05) (Fig. 4b, Supplementary Fig. 3a, Supplementary Table 19), while Abstract ScreenPrompt demonstrated higher sensitivity than humans in two reviews (p<0.05). When compared to single human-reviewer abstract screening, Abstract ScreenPrompt exhibited significantly higher sensitivity across three reviews (p<0.05), and comparable accuracy across four reviews (p>0.05) (Supplementary Fig. 3b-c). Further, head-to-head comparisons between ISO-ScreenPrompt and full dual screening revealed that ISO-ScreenPrompt had significantly higher sensitivity to humans across four reviews (p<0.05), significantly higher accuracy in one review (p<0.05), and comparable accuracy across two reviews (p>0.05) (Fig. 4c, Supplementary Fig. 3d, Supplementary Table 20).

We derived cost- and time-estimates for our LLM and human screening workflows (Supplementary Table 21-22; Methods, Cost Analysis). Traditional dual screening costs ranged from $1,385.67 to $67,872.00 USD (2,257-130,436 articles), while ISO-ScreenPrompt costs ranged from $196.43 to $12,661.30 USD at a compensation rate of $20 USD per hour. For Abstract ScreenPrompt, costs ranged from $55.25 to $4,122.49 USD, compared to $376.17 to $21,739.33 USD for single human-reviewer abstract screening (Supplementary Table 22; Methods Cost Analysis). Our LLM approach was also substantially faster. Where dual screening took approximately 69.3 to 3393.6 hours, our ISO-ScreenPrompt approach completed screening in 0.9 to 52.2 hours (Supplementary Table 22). Similarly, single human-reviewer abstract screening was estimated to take 18.8 to 1087.0 hours, whereas Abstract ScreenPrompt completed screening in 16 minutes to 15.2 hours (Supplementary Table 22). Moreover, implementing the new OpenAI batch API further reduced costs by 50% for both prompting methods (ISO-ScreenPrompt: $98.22 to $6,330.65 USD; Abstract ScreenPrompt: $27.63 to $2061.25 USD), and reduced the maximum screening time to under 24 hours.

## Discussion

Systematic review workflows are encumbered by resource- and time-intensive screening processes. Despite efforts to automate SR screening, existing tools demonstrate inadequate performance and are unable to independently screen abstracts and full-texts.^7,25,26^ Here, we leverage the capabilities of text-based LLMs, such as GPT4, for SR abstract and full-text screening. Our experiments spotlight the importance of strategic prompting, demonstrating substantial performance improvements over basic zero-shot prompting approaches, and serve as a valuable resource for researchers interested in performing LLM evaluations. In this context, we introduce Framework CoT and ISO-prompting as effective and generalizable strategies for enhancing SR screening. Our findings reveal that our Abstract ScreenPrompt and ISO-ScreenPrompt can achieve performance levels matching human reviewers, and in some cases can even surpass humans. Finally, we introduce *BenchSR*, a curated collection of SR datasets that may be used to evaluate the effectiveness of screening automation tools and support advancements in SR automation.

Previous studies evaluating LLM performance for abstract screening have primarily utilized zero-shot prompting approaches with unsatisfactory results. Guo et al.^11^ reported 59.3-100% sensitivity with GPT4 models, and Gargari et al.^13^ reported 38-69% sensitivity with GPT3.5 models. These observations demonstrate that zero-shot prompting, akin to assessing a race car’s performance without shifting gears, likely underestimates the true capabilities of LLM models for downstream tasks. Zero-shot prompting has been frequently used for other tasks such as medical question answering,^27^ medical code mapping,^28^ and disease risk stratification.^29^ Furthermore, these evaluations are often accompanied by other problematic practices, such as web browser-based evaluations (i.e., ChatGPT).^27,30–32^ Unknown system prompts, variable GPT versions, and the transient nature of chatbot context in these settings severely limit research reproducibility. Future studies should adopt more sophisticated prompting techniques, such as reasoning elicitation techniques (i.e., CoT), few-shot, and task-specific prompting. Additionally, researchers should use API calls to support reliable and reproducible research. Our analysis across various model versions demonstrated that model updates do not always improve performance, and can occasionally lead to performance drops. This variability further highlights the importance of transparent model and parameter reporting for LLM evaluations.

Our work offers several novel prompt engineering insights. CoT reasoning instructs LLMs to break down questions into intermediate reasoning steps before generating answers. However, the unstructured nature of freeform rationales can result in errors.^33^ For instance, we observed that some CoT responses did not elicit reasoning against exclusion criteria. In response, we introduced ‘Framework CoT,’ a novel prompting approach that leverages predefined criteria or ‘frameworks’ for LLMs to reason against. This facilitates a structured analysis that mimics human cognitive processes, inducing the model to systematically consider each criterion before making a final decision.

Furthermore, adjusting the proportion of GPT-CoT few-shot label distributions (i.e., balanced, exclusion-favored, inclusion-favored) did not influence model accuracy. We hypothesize that the role of labeled examples in few-shot prompting is complex and likely dependent on the task, model, and prompt design.^34,35^ The benefits of in-context learning may be attributed to the format of few-shot responses, which guide the model to better structure its output response.^34^ Few-shot GPT-Cot also resulted in worse performance than zero-shot methods. We speculate that the decreased accuracy was due to semantic contamination,^36^ where the model misinterprets the example reasoning as directly relevant to the task. Consequently, we observed heightened sensitivity at the cost of reliable reasoning, and highlight the nuanced challenge in applying few-shot prompting.

Next, we address the ‘lost-in-the-middle’ phenomenon,^21,22,37^ where as context length increases, much like stretching dough, the LLMs’ ability to adhere to instructions wanes as ‘gaps’ or ‘holes’ begin to appear within the context. This limitation of autoregressive LLMs is particularly evident in lengthy documents such as full-text articles. Our ScreenPrompt (init) prompt occasionally produced single-token outputs, possibly because the instructions to ‘think step-by-step’ were lost in context. In response, we developed *ISO-ScreenPrompt*, and repeated our prompt modules before and after the full-text content to capitalize on the LLMs’ context retrieval strength at these points.^22^ Additionally, we numerically labeled each sub-criterion to leverage the proficiency of LLMs with structured input formats.^38^ This adjustment facilitated reasoning against each specific sub-criterion, rather than broader meta-criteria. Finally, we demonstrate that self-consistency can introduce additional performance improvements at the cost of additional generations. We find these simple methods are effective strategies for bolstering LLM performance with long-context documents and anticipate that our prompting techniques can extend to other text classification domains with structured decision frameworks, such as identifying patients eligible for clinical trials^39^ and medical code mapping using electronic health records.^28^

Our research highlights that our *Abstract ScreenPrompt* and *ISO* -*ScreenPrompt* prompts can excel in abstract and full-text screening, surpassing previous tools such as Abstrackr, Rayyan, and RobotAnalyst.^25,26^ Both prompts achieve high sensitivity, with a mean value of 97.0% for Abstract ScreenPrompt and 97.5% for ISO-ScreenPrompt, across ten different reviews. Our prompts also maintain robust accuracy, with 88.4% mean accuracy for Abstract ScreenPrompt and 93.6% mean accuracy for ISO-ScreenPrompt. Furthermore, we did not modify or attempt to optimize the eligibility criteria or study objectives from each review, highlighting that our approach can be readily implemented. Our findings ultimately underscore the efficacy of prompt design in real-world applications and sets a new standard for SR screening automation efforts, which have historically demonstrated unsatisfactory performance.^25,26^

We address a critical gap within the realm of SR screening automation by providing one of the most comprehensive reproducibility evaluations for gold-standard dual human screening. Previous evaluations have been limited to title/abstract screening phases across three or fewer SRs.^23^ In contrast, our study performed full dual screen evaluations across five reviews covering four common Oxford CEBM question types. Our reviewers were calibrated for screening proficiency, and revealed that dual human screening can be error prone, with many articles erroneously excluded at the full-text stage.

Crucially, our ISO-ScreenPrompt approach is the first to demonstrate that SR automation efforts can match, and in some instances, outperform gold-standard human dual screening. ISO-ScreenPrompt had higher sensitivity than dual screening in all reviews, with significantly higher sensitivity in four of five reviews. Furthermore, it maintained comparable or greater accuracy in three reviews. Barring full-text accessibility issues, this superior sensitivity and comparable accuracy could revolutionize the SR process by potentially eliminating the need for an initial human screening phase of abstracts. Instead, reviewers could initiate data extraction on the subset of articles deemed included by ISO-ScreenPrompt, and iteratively remove ‘false-positive’ articles.

Abstract ScreenPrompt, which displayed superior sensitivity relative to dual abstract screening, offers another viable alternative for immediate implementation. While it may not replace dual abstract screening workflows, it achieved comparable accuracy to single human reviewers in four reviews and exceeded human sensitivity in all five reviews, with significantly higher sensitivity in three reviews. These findings suggest that Abstract ScreenPrompt could reliably serve as a single human reviewer vote at the abstract stage without compromising quality. Finally, both ISO-ScreenPrompt and Abstract ScreenPrompt are associated with substantial cost- and time-savings. Where traditional SR screening can take weeks to months, our approach completed screening in minutes to hours, freeing reviewers for deeper scientific evaluations and accelerating synthesis of study conclusions.

Finally, our work addresses a critical gap in the realm of SR screening by introducing *BenchSR*, a growing collection of 10 SRs that span four types of medical questions across nine different clinical domains. Existing benchmarks for SR screening such as CLEF,^40^ Seed Collection,^41^ and Cohen^42^ have important limitations. The CLEF and Seed Collection benchmark lack critical study metadata, such as detailed inclusion and exclusion criteria; and feature incomplete datasets with only article identifiers, necessitating additional efforts to gather article abstracts and/or full-texts. Furthermore, the Cohen benchmark is no longer publicly accessible.

*BenchSR* provides (i) a comprehensive set of all included and excluded abstracts (with article identifiers) found during the initial search, (ii) essential study metadata not typically reported (detailed eligibility criteria), sourced directly from the original study authors, and (iii) a non-online hosting solution to mitigate concerns about LLM pre-training data contamination. Study metadata is currently limited to eligibility criteria and study objectives, but will be expanded significantly in future work (i.e., data extraction templates). Future research should focus on enhancing transparency and completeness in reporting SRs, including detailed study metadata and data extraction templates, to better support and refine automation efforts. We invite other researchers to contribute to our growing *BenchSR* benchmark.

Our study has several limitations. Although we apply our analysis across a wide range of SRs, the generalizability of our findings to other clinical questions, non-English SRs and other review methodologies (e.g. scoping reviews) requires further study. However, our prompt templates would likely perform well in reviews with PICO-structured inclusion/exclusion criteria. Furthermore, our work may extend to non-English SRs as LLM models continually improve language accessibility. Our full-text analysis was limited by the availability of freely accessible texts in the PubMed Central (PMC) database. While we made efforts to enhance our dataset by manually scraping ‘included’ full-texts for select reviews, we did not manually gather all ‘excluded’ texts due to accessibility and resource constraints. Our LLM screening was also solely conducted on text content. Including figures and tables could potentially enhance performance and accuracy and warrants future investigation. Moreover, our analysis explored Gemini Pro, GPT3.5, and GPT4-0125-preview, GPT4-Turbo-0409, and GPT4o-0513, but not the Claude 3 LLMs due to regional availability issues. Due to GPT4 context length limitations (128k tokens), we only tested few-shot prompting on abstracts. Furthermore, while we surveyed a wide range of prompting techniques, we did not explore every possible method. The influence of subtle changes (i.e., word choice) may further optimize models. Our focus on developing ready-to-use and generally applicable prompts meant that we did not modify original SR inclusion and exclusion criteria. However, we suspect that additional optimizations could improve performance.

In conclusion, our study underscores the importance of deliberate prompting to achieve human-level performance for SR screening. The promising results from ISO-ScreenPrompt and Abstract ScreenPrompt marks a major advancement in the automation of SR processes. We encourage further research into the capabilities of LLMs for other SR tasks, such as data extraction and meta-analysis. Fully-automated SRs will revolutionize evidence-based practice and offer indispensable value to medicine and beyond.

## Methods

### Datasets and data acquisition

We acquired abstract screening data from 10 distinct SRs spanning eight unique clinical domains through purposeful convenience sampling based on Oxford CEBM SR question types. To compile these datasets, we engaged with systematic review investigators at the University of Calgary and the University of Toronto. We extracted study information concerning review objectives from the published manuscript or PROSPERO protocol, and contacted study authors for internal reviewer inclusion/exclusion criteria.

In brief, ‘SeroTracker’ (ST) was a living SR of prevalence, exploring observational cohort studies that reported single-estimate prevalence in the context of COVID-19.^43^ ‘Reinfection’ was an SR of intervention benefits that assessed the comparative effectiveness of vaccination and past COVID-19 infection relative to past COVID-19 infection alone in observational studies reporting associations.^44^ ‘PA-Outcomes’ was an SR of intervention benefits comparing clinical outcomes between surgery and medication treatments in patients with primary aldosteronism.^45^ ‘PA-Testing’ was an SR of diagnostic test accuracy evaluating guideline-recommended confirmatory tests (i.e., saline infusion test, salt loading test, fludrocortisone suppression test, and captopril challenge test) relative to a reference standard.^46^ ‘Sepsis’ was an SR of intervention benefits that assessed the comparative effectiveness and safety of fludrocortisone plus hydrocortisone, hydrocortisone alone and placebo/usual care in adults with septic shock.^47^ ‘Spinal’ was an SR of diagnostic test accuracy that evaluated the efficacy of intraoperative neurophysiological monitoring among patients undergoing spine surgery for any indication.^48^ ‘Calcium-HA’ was an SR of intervention benefits that compared the outcomes of routine calcium administration to no calcium administration for cardiac arrest in adults or children.^49^ ‘Infant-NO’ was an SR of prognosis that assessed whether an immediate response to inhaled nitric oxide therapy was associated with reduced mortality in preterm infants with hypoxemic respiratory failure and pulmonary hypertension.^50^ ‘Meds-HA’ was an SR of SRs, covering intervention benefits, that identified medications that affected hospital admissions.^51^ ‘SVCF’ was an SR of prognosis that evaluated the association of low SVC flow, diagnosed in the first 48 hours after birth echocardiography, with neurological morbidity and mortality, among very preterm neonates.^52^

Included and excluded abstracts were downloaded from Covidence (Veritas Health Innovation, Melbourne, Australia), a systematic review screening software. Abstracts were stored in csv files. We downloaded all ‘Included’ (included articles at full-text screening), ‘Excluded’ (excluded articles at full-text screening), and ‘Irrelevant’ (excluded articles at abstract screening) articles. Excluded and irrelevant articles were collated to form our excluded article dataset. To obtain full-text articles, we utilized the PMC ID Converter API to convert abstract DOIs into Pubmed IDs (PMIDs), followed by the BioC API for PMC to obtain XML full-text files from each abstract PMID. Full-texts with over 150,000 characters were excluded (i.e., abstract conference proceedings). Human authors (CC, JS, DC, RA, NB) manually scraped all ‘included’ full-text articles not captured by the BioC API for SRs selected in our head-to-head human-screening performance evaluation in plain text files.

The datasets described here including labeled abstract sets, and all associated metadata, are available at (link made available upon publication). Full-text articles are not provided due to copyright concerns. We invite other researchers to contribute to our growing *BenchSR*.

### Abstract Prompt Approaches

During the evaluation of GPT4 abstract screening performance, we applied eight distinct prompting methodologies, discussed in more detail below. A brief illustration and the structure for each prompt is shown in Fig. 2a and Table 2.

#### Zero-shot prompting

Models are prompted with only instructions for completing the task at hand, including the required context directly related to the task, i.e. abstract text to analyze. No additional context or examples are provided. For our zero-shot prompts, we adapted a prompt from Guo et al.^11^ that evaluated GPT4 and GPT3.5 SR screening performance. We similarly specified that the LLM returns only a single token output with its final decision.

#### Random few-shot prompting

Models are prompted with the instruction-task at hand, with the addition of (k=n) labeled examples relevant to the task at hand. For our purposes, the examples are randomly selected and accurately labeled as included (‘YYY’) or excluded (‘XXX’). We set k=10 (5 included, 5 excluded), in agreement with MedPrompt and general prompting guidelines.^10^

#### Zero-shot Chain-of-Thought (CoT)

Models are prompted with the instruction-task at hand, along with additional natural language statements, such as “*Let’s think step by step* ” to encourage the model to generate intermediate reasoning steps before generating a final answer.^18^

#### Zero-shot Framework CoT

We devised a new prompting approach, termed ‘Framework CoT,’ wherein we deliberately prompt the model to reason against each criterion. Similar to zero-shot prompting, no additional context or examples are provided.

#### Zero-shot ScreenPrompt

We included additional well-defined study objectives (adapted from published manuscripts) to our Framework CoT prompt to better orient our prompt to screening tasks.

#### Zero-shot Abstract ScreenPrompt

We further incorporated additional context that acknowledged the inherent content limitations of abstracts and goals of inclusivity to our ScreenPrompt prompt to better orient our prompt to the task of abstract screening.

#### Few-shot GPT-CoT Abstract ScreenPrompt

We prompted our model with *Abstract ScreenPrompt*, but also included additional examples that contained *Abstract ScreenPrompt* GPT-generated reasoning. We discarded answers that did not match the ground truth label to uphold ‘correctness’ in example reasoning. This approach was adapted from MedPrompt, which has suggested that GPT-generated CoT reasoning can outperform human experts, as well as automate the CoT example process. We set k=10 (5 included, 5 excluded), in agreement with MedPrompt and general prompting guidelines.^10^

#### Self-consistency

To address variability in model outputs due to stochasticity, we conducted repeat evaluations using different seed parameters for the same prompt. The final answer is the decision to ‘include’ or ‘exclude’ according to majority vote. We set the number of self-ensembles to 11, in accordance with literature recommendations.^20^ For our AUROC analysis, we adjusted the self-consistency threshold, which dictates the number of votes required for an article’s inclusion or exclusion, within a range from 0 to 12 votes (0 always include; 12 always exclude). This method enables us to fine-tune sensitivity and specificity by leveraging the consensus of multiple model generations.

### Full-text Prompt Approaches

During the evaluation of GPT4 full-text screening performance, we applied six distinct prompting methodologies, discussed in more detail below. A brief illustration and the structure for each prompt is shown in Fig. 3a, and Table 2. Our standard ‘prompt structure’ used for abstract screening consisted of: {Pre-Prompt}, {Inclusion Criteria}, {Exclusion Criteria}, {Article}, {Instructions}. Prompt elements refer to all modules except {Article}

#### ScreenPrompt (Init)

We modify our ‘prompt structure’ by appending all of our prompt elements ({Objectives}, {Inclusion criteria}, {Exclusion criteria}, {Instructions}) to the start of the prompt, before the {Article} text content.

#### ScreenPrompt (Fin)

We modify our ‘prompt structure’ by appending all of our prompt elements to the end of the prompt, after the {Article} text content.

#### *ScreenPrompt* (Init + Fin-Instructions)

We modify our ‘prompt structure’ by appending all of our prompt elements to the start of the prompt, before the {Article} text content. We additionally append {Instructions} after the {Article} text content.

#### *ScreenPrompt* (Init + Fin)

We modify our ‘prompt structure’ by appending all of our prompt elements to the start of the prompt (before the {Article} text content) and end of the prompt (after the {Article} text content).

#### Numbered ScreenPrompt

We preserve the semantic content of our prompt and add number symbols for each individual inclusion and exclusion sub-criterion. We removed meta-criteria headings (i.e., population, intervention, etc.) where applicable.

#### ISO-ScreenPrompt

We apply a combination of our Numbered ScreenPrompt where each individual inclusion/exclusion sub-criterion is numbered, and our (Init + Fin) prompt structure, where all prompt elements are appended to the start and end of the prompt.

### Prompt Testing Methodology

To avoid concerns of prompting ‘overfitting’ during training and testing our prompt engineering process, we apply principles of sound testing methodology for machine learning studies and randomly sampled train/validation/test splits for our downstream analysis. Our iterative prompt optimization process was only performed on train splits, and we validated the performance of our optimized prompting strategy on validation splits. Test splits were held out from prompt engineering consideration until the final testing phase to assess real world, or ‘eyes-off’ performance.

To determine the minimum sample size for abstract evaluation, we used the Cochran’s sample size formula.^53^ We set our desired confidence level to 95% (p=0.05), Margin of Error (MoE) to 5%, and model performance to 50% (assuming 50% chance of inclusion/exclusion labels), resulting in a minimum sample size of 385 abstracts.

Due to its substantial dataset, we utilized data from the SeroTracker SR (n=130k excluded abstracts, n=3000 included abstracts) to derive balanced sets of included and excluded abstracts for our train, validation, and test splits (Train: n=200 included, n=200 excluded; Validation: n=200 included, n=200 excluded; Test: n=200 included, n=200 excluded;). Furthermore, we randomly sampled sets of included and excluded abstracts for our GPT-CoT prompting to prevent cross contamination (GPT-CoT: n=100 included, n=100 excluded).

Following our prompt-optimization procedures, we also tested the performance of our prompting strategy on the ST Test split, and 9 other SR abstract datasets to model its real-world performance and generalizability across different SR domains. As these datasets were held-out from any prompt engineering steps, we included all ‘included’ articles, and randomly sampled ‘excluded’ articles to reach a test sample size of 400 articles.

We replicated the same procedures for full-text evaluations. In brief, we randomly sampled the SeroTracker SR (n=5137 excluded, n=1797 included PMC-scraped full-texts) to derive balanced training and validation datasets (Train: n=200 included, n=200 excluded; Validation: n=200 included, n=200 excluded). Furthermore, we randomly sampled sets of included and excluded abstracts for our GPT-CoT prompting to prevent cross contamination (GPT-CoT: n=100 included, n=100 excluded). The test dataset was composed of all remaining articles (Test: n=1297 included, n=4637 excluded), but we opted to randomly sample a smaller subset (n=200 included, n=200 excluded) in the interest of cost and rate limits.

Following our prompt-optimization procedures, we tested the performance of our *ISO-ScreenPrompt* prompting strategy for full-text screening on the ST Test split, and 9 other SR datasets. For each SR evaluation, we set our sample size to n=400, and incorporated all included articles that were available from the PMC web scrape. For instances where we were unable to obtain a sample size of 400 (i.e., spinal), we evaluated all available ‘included’ and ‘excluded articles. As previously mentioned, human researchers performed additional manual scraping of all ‘included’ articles for SRs chosen in our head-to-head evaluation of human screening performance.

### Head-to-head comparisons with human screening

We assembled a panel of four researchers with past SR experience (1 BSc, 3 MSc) to perform end-to-end SR screening while adhering to canonical dual reviewer screening protocols.^1^ The reviewers blinded from our study objectives (comparing GPT performance with human reviewers), and were only provided with the list of references for screening, internal inclusion/exclusion criteria, and SR objectives (Supplementary Note 1). Researchers were instructed to perform standard SR screening, with a sensitive abstract screen followed by an accurate full-text screen. Screening for each review was performed independently by two reviewers in duplicate. Any conflicts during screening were resolved by a 3rd independent reviewer. We analyzed the performance of single human-reviewer abstract screening, dual human abstract screening (based on independent votes from two reviewers and resolved conflicts at the abstract stage), and complete dual screening (based on independent full-text votes and resolved conflicts at the full-text stage). For comparisons of single human-reviewer abstract screening with Abstract ScreenPrompt, we randomly selected the performance of one reviewer from the two reviewers for each review.

We calibrated our reviewers according to screening proficiency by having prospective reviewers first screen a ‘calibration set’ of abstracts. This set was sourced from a prior study by the SeroTracker group,^54^ which assessed the performance of dual human reviewer workflows. Notably, the SeroTracker researchers were experienced SR screeners, having contributed to the SeroTracker living SR for over a year, and represent a high-performing baseline for screening accuracy. The results from this prior study, and calibration set were used in head-to-head comparison analysis for the SeroTracker (ST) dataset. Reviewers were recruited for subsequent screening tasks if their accuracy was within 5% of the performance benchmarks set by the SeroTracker group. The performance metrics of our reviewers were highly similar to those by the SeroTracker group and are detailed in Supplementary Table 18.

For a representative comparison of GPT4-0125-preview and human screening performance, we used stratified random probability sampling across the four Oxford CEBM review questions, selecting one SR for each type. Our sample included various datasets: the SeroTracker dataset for reviews of prevalence (adapted from Perlman-Arrow et al.^54^), the Reinfection dataset for reviews of intervention benefits, the PA-Testing dataset for reviews of diagnostic test accuracy, and SVCF dataset for reviews of prognosis. Due to the high volume of SR studies focusing on intervention benefits, we also included the PA-Outcomes dataset in our sample through additional random sampling.

### Time and Cost Analysis

To derive cost estimates for traditional human dual screening for each review, we began by determining the total number of abstracts and full-texts screened at each stage. For abstracts, we used the total number of articles for a given review. For full-texts, we counted the total number of ‘Included’ and ‘Excluded’ articles from Covidence. Based on previous studies, the time required to screen a single abstract ranges from 20-461 seconds,^5,54,55^ and 4.3-20 minutes for a single full-text article.^55,56^ We aligned with Perlman-Arrow et al.^54^ due to our use of the same ST dataset and set the screening time at 30 seconds per abstract and 10 minutes per full-text.

We calculated time estimates for human reviewers by multiplying the total number of screened abstracts by 30 seconds and the total number of screened full-texts by 10 minutes. The resulting value was then doubled to derive the cost of dual human screening (in duplicate). For single human abstract screening, we only took the total number of abstracts. We assumed a compensation rate of $20 USD per hour for human reviewers to calculate costs. It is important to note that our cost estimates do not consider the additional costs of conflict screening (additional abstracts and full-texts screened due to conflicts between reviewer decisions), and likely represents a conservative estimate of human screening costs.

For our ISO-ScreenPrompt and Abstract ScreenPrompt approach using the GPT4-0125-preview, we calculated the exact cost for each full-text and abstract run used in our head-to-head comparison analysis ($10 per million input tokens, $30 per million output tokens, OpenAI Pricing). We then derived the cost per article by dividing the total cost of our runs by the number of articles in each run. We multiplied this cost per article by the total number of articles for each review to estimate the cost of our ISO-ScreenPrompt and Abstract ScreenPrompt approach for each review. To derive estimates for our two approaches following implementation of the OpenAI Batch API, we divided the estimated cost by two (Batch API offers a 50% discount). We derived time estimates by obtaining the time per article (ISO-ScreenPrompt: 1.44s/full-text; Abstract ScreenPrompt: 0.42s/abstract), and multiplied this estimate against the total number of articles for each review.

### LLM API and LLM Evaluations

We used GPT3.5-Turbo-0125 (GPT3.5), GPT4-0125-preview, GPT4-Turbo-0409, GPT4o-0513, Gemini Pro, open Mixtral-8×22-0424, and Mistral-Large-0224 models and compared their performance for abstract and full-text screening with our optimized ‘Abstract ScreenPrompt’ and ‘ISO-ScreenPrompt prompts’, respectively. For all models, we set the maximum tokens to 2048, and used the advised default settings. GPT model settings were set to temperature=1, top_p=1, frequency_penalty=0, presence_penalty=0. The Gemini pro model settings were set to temperature=1, top_p=1. The Mixtral-8×22-0424, and Mistral-Large-0224 model settings were set to temperature=0.7, top_p=1. We note that for Gemini Pro, the default temperature was changed to temperature=0.9 as of May 2024 (after our testing). We set the seed for all of our LLM models at 0. For our self-consistency analysis (n=11), we set different seed parameters from 0 to (n-1) for each repeat evaluation.

To evaluate the model responses, we required an output containing an ‘evaluation token’: either “XXX” (exclude) or “YYY” (include). When responses contained both “XXX” and “YYY”, we checked the last 500 characters of the output and used the final instance of the token.

We conducted our initial runs synchronously, sending requests directly to the API and waiting for responses in the same network call. This approach allowed us to easily retry individual calls to a model’s API. For each abstract, if a response was interrupted by a technical error (rate limit, disconnect, timeout) or was missing an ‘evaluation token’, we retried the instance up to three times. If all three attempts failed, we deemed the request as ‘unparseable’ and disqualified the request from the total count, occasionally reducing the set of 400 articles. During the writing of this paper, OpenAI released the Batch API, which significantly reduced costs and improved run times. Shifting to this infrastructure meant we could no longer efficiently retry individual abstracts, but the new system greatly reduced technical errors and rarely resulted in missing “evaluation tokens”.

### Data analysis

We assessed the performance of our prompts by analyzing accuracy, sensitivity, and specificity metrics, and calculated binomial 95% confidence intervals (CIs) for sensitivity and accuracy. “Unparseable requests” were discarded and the metrics computed only for those responses that were parseable. To compare the performance of our Abstract ScreenPrompt and ISO-ScreenPrompt with zero-shot methods across all ten reviews, we calculated 95% CIs using the Clopper-Pearson method,^57^ employing the binom package in R. This approach was necessary due to the small sample size of included articles in some reviews (n<10). We determined statistical significance and 95% CIs with a two-tailed test computing the difference between two independent binomial proportions in our comparative analysis of human versus LLM screening performance.

## DATA AVAILABILITY

Researchers can access our data via the following github repository (link made available upon publication).

## CODE AVAILABILITY

All code used for experiments in this study can be found in a github repository (link made available upon publication).

## Additional Information

## Acknowledgements

We would like to thank Dr. Michelle Baczynski for her support in providing access to the Infant-NO dataset. We would like to thank Dr. Mohammed Ali Alvi for his support in providing access to the Spinal dataset.

## Author Information

These authors contributed equally: C.C., J.S. These authors jointly supervised the work: R.K.A., N.B.

### Author contribution statements

C.C., J.S., N.B., R.K.A contributed to the conception and design of the work. C.C., J.S., R.A., R.K., M.C., J.G., R.S., D.C., I.D., B.T., M.F., P.R., A.A.L., D.E.W., H.W., M.W., N.B., contributed to the data acquisition and curation. C.C., J.S., contributed to original investigation and formal analysis. C.C., J.S., R.A., D.E.W., N.B., R.K.A., interpreted results and provided feedback on the study. C.C., J.S., R.A., D.E.W., contributed to validation of the data. C.C., J.S., H.W., M.W., N.B., R.K.A., contributed to project supervision. C.C., J.S., R.A., N.B., prepared the original draft of the manuscript with input form all co-authors. All authors were responsible for review and editing of the manuscript. All authors debated, discussed, edited, and approved the final version of the manuscript.

## Supplementary Information

**Supplementary Figure 1:**
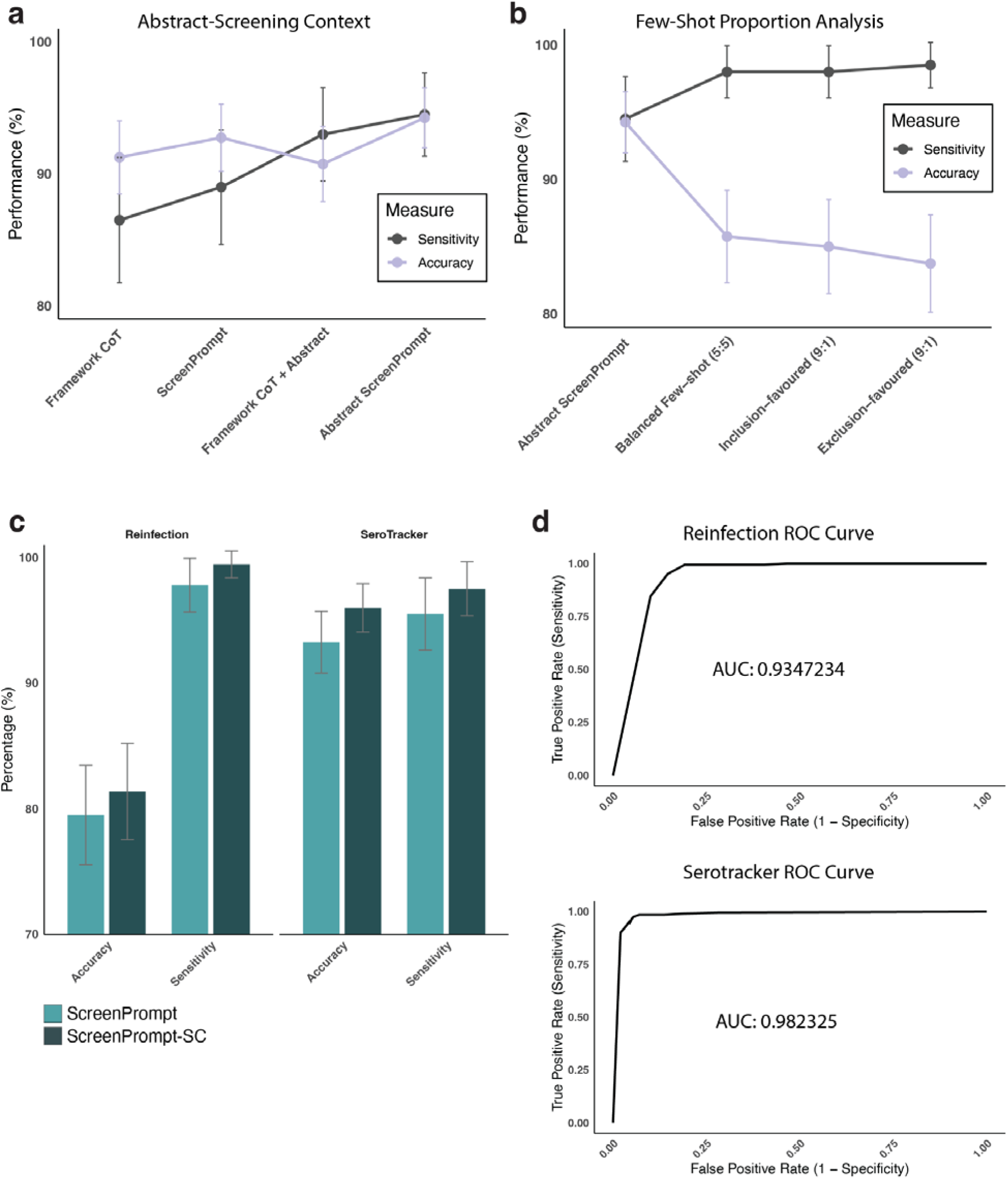
Abstract screening prompt optimization. a) Performance comparison of different abstract prompting methodologies relating to the addition of study objectives and abstract-specific considerations on the SeroTracker (ST) training dataset, (n=400) showing accuracy and sensitivity. Error bars represent 95% CIs for binomial proportions. b) Performance comparisons of different few-shot prompting methodologies on the ST training dataset, showing accuracy and sensitivity. Prompts compare differing proportions of inclusion-labeled and exclusion-labeled few-shot examples. Error bars represent 95% CIs for binomial proportions. c) Barplot displaying the performance of Abstract ScreenPrompt sensitivity and accuracy with and without self-consistency (SC) in SeroTracker and Reinfection test datasets (n=400). Error bars represent 95% CIs for binomial proportions. d) Receiver operating characteristics (ROC) curves generated from differing self-consistency thresholds (number of votes needed, 0-12) for article inclusion in SeroTracker and Reinfection test datasets.

**Supplementary Figure 2:**
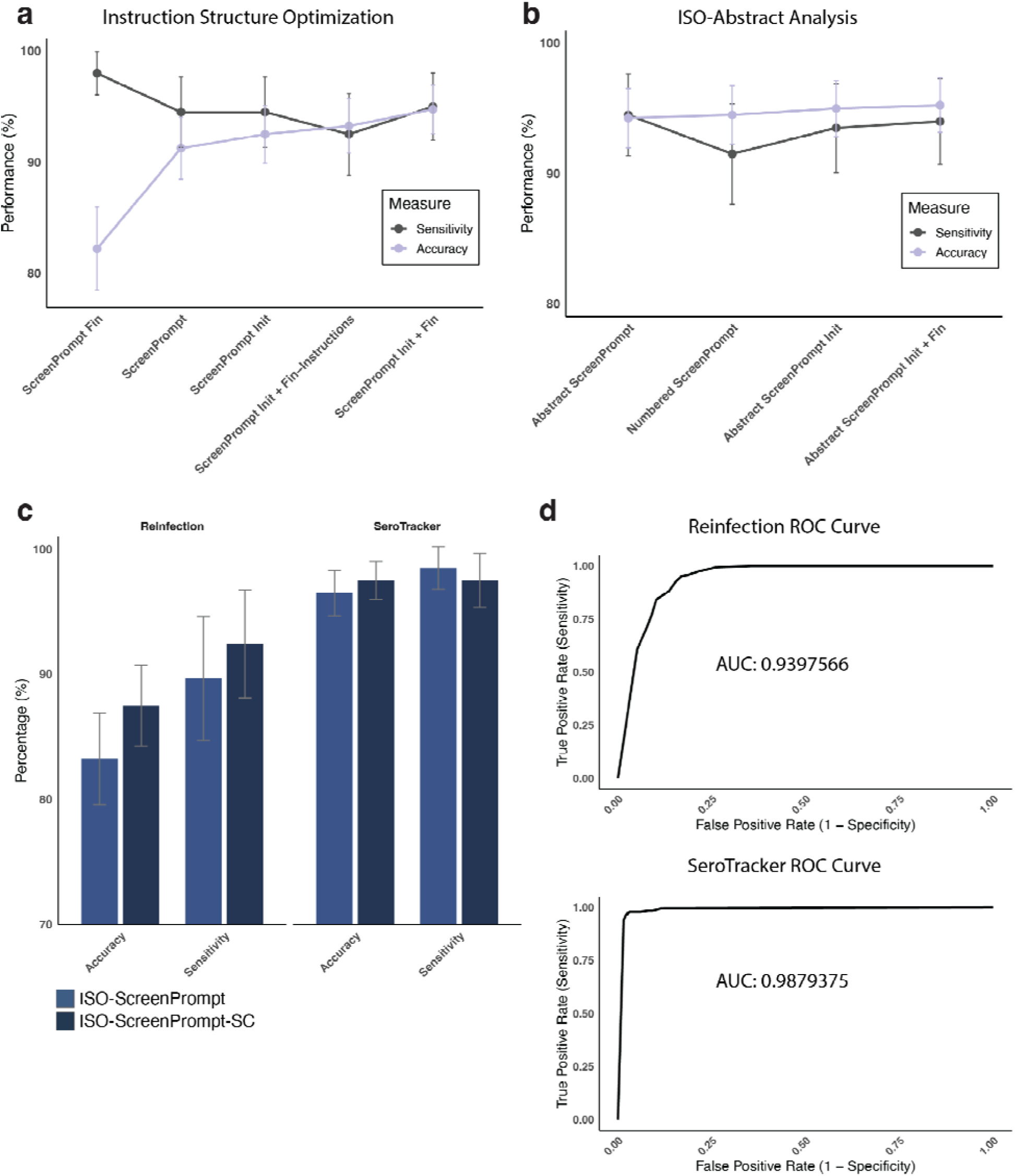
Full-text screening prompt optimization. a) Performance comparison of different full-text prompting methodologies with modifications in prompt structure on the ST training dataset (n=400), showing accuracy and sensitivity. Error bars represent 95% CIs for binomial proportions. b) Performance comparison of abstract screening performance with modifications in prompt structure on the ST training dataset, showing accuracy and sensitivity. Error bars represent 95% CIs for binomial proportions. c) Barplot displaying the performance of ISO-ScreenPrompt sensitivity and accuracy with and without self-consistency (SC) in SeroTracker and Reinfection test datasets (n=400). Error bars represent 95% CIs for binomial proportions. d) Receiver operating characteristics (ROC) curves generated from differing self-consistency thresholds (number of votes needed, 0-12) for article inclusion in SeroTracker and Reinfection test datasets.

**Supplementary Figure 3:**
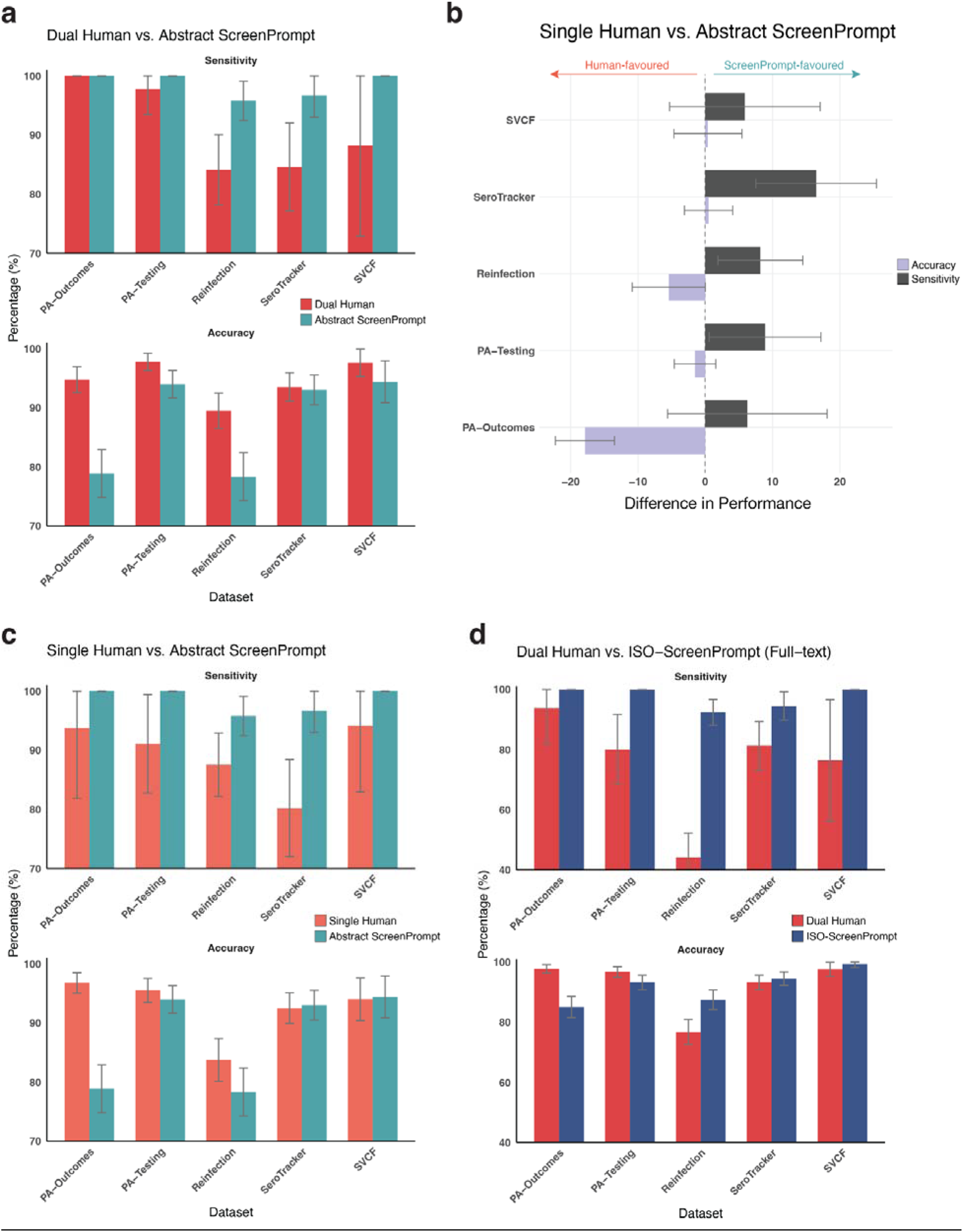
ScreenPrompt and ISO-ScreenPrompt vs. human screening. a) Barplot displaying the performance of Abstract ScreenPrompt and dual human abstract screening sensitivity and accuracy. Error bars represent 95% CIs for binomial proportions. b) Difference in performance between single human-reviewer and Abstract ScreenPrompt across five systematic reviews (SVCF n=167, SeroTracker n=400, Reinfection n=400, PA-Testing n=400, PA-Outcomes n=400). The barplot shows differences in accuracy and sensitivity, with human-favored differences as negative (left) and ScreenPrompt-favored differences as positive (right). Error bars represent 95% CIs for the difference in binomial proportions. c) Barplot displaying the performance of Abstract ScreenPrompt and single-human reviewer screening sensitivity and accuracy. Error bars represent 95% CIs for binomial proportions. d) Barplot displaying the performance of ISO-ScreenPrompt and full dual human screening sensitivity and accuracy. Error bars represent 95% CIs for binomial proportions.

**Supplementary Table 1.** Abstract prompt engineering performance.

| Dataset | Prompting Strategy | Accuracy | Sensitivity | Specificity |
| --- | --- | --- | --- | --- |
| Training<br>(n=200 include,<br>n=200 exclude) | Zero-shot - | 65.0%<br>260/400 | 30.0%<br>60/200 | **100.0%**<br>**200/200** |
|  | Random Few-shot<br>Balanced:<br>5 include<br>5 exclude | 78.0%<br>312/400 | 56.0%<br>112/200 | 100.0%<br>200/200 |
|  | Zero-shot Chain of Thought (CoT) | 86.3%<br>345/400 | 74.5%<br>149/200 | 98.0%<br>196/200 |
|  | Zero-shot framework CoT | 91.3%<br>365/400 | 86.5%<br>173/200 | 96.0%<br>192/200 |
|  | ScreenPrompt | 92.8%<br>371/400 | 89.0%<br>178/200 | 96.5%<br>193/200 |
|  | Abstract ScreenPrompt | **94.3%**<br>**377/400** | 94.5%<br>189/200 | 94.0%<br>188/200 |
|  | Few-shot GPT-CoT<br>ScreenPrompt<br>Balanced:<br>5 include<br>5 exclude | 85.8%<br>343/400 | **98.0%**<br>**196/200** | 73.5%<br>147/200 |
| Validation<br>(n=200 include,<br>n=200 exclude) | Abstract ScreenPrompt | 94.3%<br>377/400 | 96.0%<br>192/200 | 92.5%<br>185/200 |

**Supplementary Table 2:** Inclusive Language Prompting.

| Dataset | Zero-shot prompt | Accuracy | Sensitivity | Specificity |
| --- | --- | --- | --- | --- |
| Training<br>(n=200 include,<br>n=200 exclude) | ...<br>Only type “YYY” for included articles or “XXX” for excluded articles to indicate your decision. Do not type anything else. | 65.0%<br>260/400 | 30.0%<br>60/200 | **100.0%**<br>**200/200** |
|  | ...<br>We will conclude by outputting (on the very last line) 'XXX' if the paper warrants exclusion, or 'YYY' if inclusion is advised or uncertainty persists. We must output either 'XXX' or 'YYY'. | 68.3%<br>273/400 | 36.5%<br>73/200 | 100.0%<br>200/200 |

**Supplementary Table 3:** Exemplar Wrong Outputs during Abstract Prompt Engineering.

| Prompt | Example LLM Output | Author Feedback |
| --- | --- | --- |
| Zero-shot CoT: | {Model output}<br>**\*\*Population\*\*:**<br>1. The study involves humans of any age. [Meets criteria]<br>2. The subjects include those with suspected disease without requiring them to have been deliberately sampled for being antigen positive. [Meets criteria]<br>**\*\*Study Design\*\*:**<br>1. This is a cross-sectional serologic survey. It fits the inclusion criteria for study design as it is one of the specified designs interested in measuring seroprevalence. [Meets criteria]<br>2. Serology measurements were presumably taken at a single time point as common in cross-sectional designs. [Meets criteria]<br>**\*\*Special Design\*\*:**<br>1. This paper is not a systematic review or meta-analysis but rather a primary seroprevalence study. However, non-systematic reviews/meta-analyses are not listed as exempt from consideration; the inclusion criteria express a specific interest in systematic reviews and meta-analyses without excluding primary studies. [Meets criteria]<br>**\*\*Sampling\*\*:**<br>1. The method of sampling is not entirely detailed in the abstract, but given that it's a cross-sectional study among a defined number of participants (888), it suggests a deliberate approach to sampling, which is compliant with the inclusive approach to any sampling method. [Meets criteria]<br>**\*\*Types of Evidence\*\*:**<br>1. Assuming this is a published academic article (given its detailed results and conclusions, along with the referencing of copyright by Wiley & Sons Ltd.), it meets the criteria for the types of evidence being sought. [Meets criteria]<br>**\*\*Outcome Measures\*\*:**<br>1. The study reports a seroprevalence estimate, with 30% of the study volunteers having detected neutralizing antibodies. [Meets criteria]<br>2. The number of participants enrolled in the study is clearly stated as 888. [Meets criteria]<br>3. While the study sampling end date/week is not explicitly mentioned, this detail might be present in the full text; however, for abstract screening, its absence is noted but its potential availability in the full report could still mean the study might meet the criteria upon full review. [Unclear]<br>4. The study clearly reports the location as being in Egypt, fitting the requirement to categorize the location of the study. [Meets criteria]<br>Given that all but one of the clear inclusion criteria are met with only the study sampling end date/week not explicitly stated in the abstract, there's a stronger leaning towards inclusion, with the assumption that further details necessary to fully satisfy all criteria might be found in the full text.<br>**\*\*Conclusion: YYY\*\*** | Output did not assess/evaluate exclusion criteria<br>→ Led to Framework CoT |
| Zero-Shot Framework CoT | Let's evaluate the abstract provided against the inclusion and exclusion criteria systematically.<br>### Inclusion Criteria Evaluation: | Erroneously inferred study objectives |
|  | 1. **Population:** The study includes humans of any age, which meets the criterion. It doesn't specify excluding only those with suspected or confirmed COVID-19, suggesting broader inclusion, thus meeting this requirement.<br>2. **Study design:** The study design focuses on the qualitative assay of immunoglobulins G and M in diagnosed patients, aiming to assess transmissibility and the possibility of reinfection. This can be interpreted as part of a broader sero-survey effort, but it appears more focused on specific diagnostic outcomes rather than prevalence estimation. It lacks a clear mention of being a cross-sectional, repeated cross-sectional, or cohort study as defined by our criteria.<br>3. **Special design:** The study does not present itself as a systematic review or meta-analysis of seroprevalence studies.<br>4. **Sampling:** The method of sampling is not explicitly described, but the study included a total of 553 patients, indicating a form of sampling was done.<br>5. **Types of evidence:** Presumably, this is published or unpublished academic literature, considering it has authorship and an abstract, thus meeting this criterion.<br>6. **Outcome measures:** The abstract reports on IgG and IgM levels in patients, implying an assessment of seroprevalence to some extent. However, it does not clearly state it reports a seroprevalence estimate in terms of the proportion of the population with detectable antibodies. The number of participants is reported, but it lacks clear information on study sampling end date/week or detailed location categorization.<br>**Exclusion Criteria Evaluation:**<br>1. **Population:** This study is conducted in humans, thus not meeting any of the population-based exclusion criteria directly.<br>2. **Study design:** The study could arguably fall under the exclusion due to its focus on evaluating immunological responses rather than explicitly being a cross-sectional or cohort design aimed at seroprevalence estimation.<br>3. **Types of evidence:** There's no mention of using excluded multimedia sources or unassociated dashboards.<br>4. **Outcome measures:** The abstract mainly discusses the detection of IgG and IgM antibodies but does not only report on antigen prevalence. However, it does not fully comply with reporting requirements around the study sampling end date/week or location detail.<br>**Conclusion:**<br>**<u>Given the specific focus on evaluating immunological responses in patients rather than estimating the seroprevalence across a population,</u>** and the absence of clear compliance with several crucial inclusion criteria (study design specifics, complete outcome measure reporting), the study appears to lean more towards exclusion based on its design and reported outcomes.<br>However, considering the nature of abstract reviews, the decision leans on available information which might not fully capture the study's relevance to the systematic review aims. Given the potential relevance of immunological response data to the broader topic of seroprevalence and immunity estimation, there is a fine line of interpretation.<br>Considering the primary focus on diagnostic outcomes as opposed to population-level antibody prevalence estimation and the critical unfulfilled aspects of both inclusion and exclusion criteria, the conclusion leans towards:<br>XXX | → Led to ++ Objectives |

**Supplementary Table 4:**
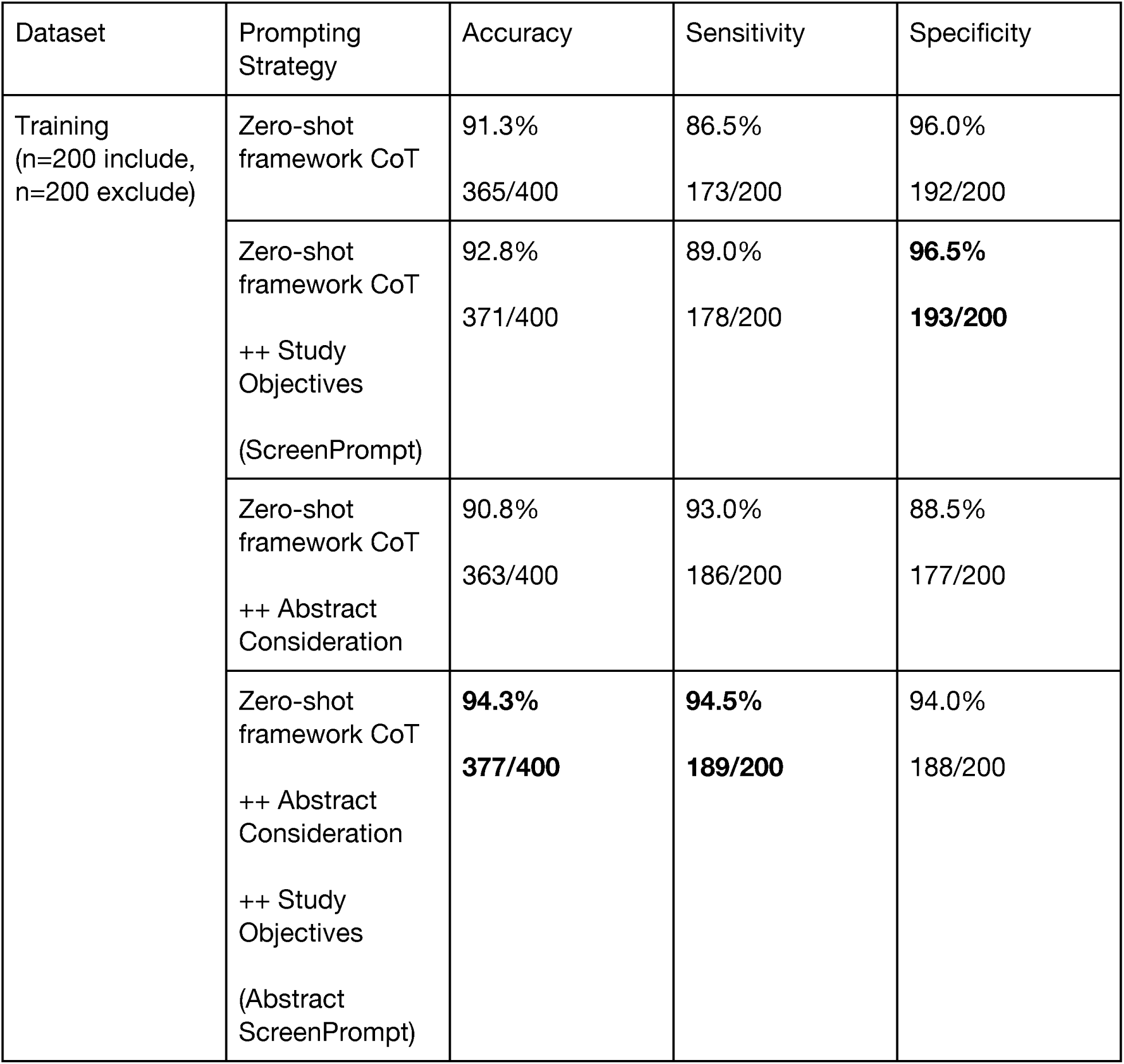
Abstract ScreenPrompt Ablation Prompting.

**Supplementary Table 5:** Few-shot GPT-CoT Label Proportion Analysis.

| Dataset | Prompting Strategy | Accuracy | Sensitivity | Specificity |
| --- | --- | --- | --- | --- |
| Training<br>(n=200 include,<br>n=200 exclude) | Few-shot GPT-CoT<br>ScreenPrompt<br>Balanced:<br>5 include<br>5 exclude | **85.8%**<br>**343/400** | 98.0%<br>196/200 | **73.5%**<br>**147/200** |
|  | Few-shot GPT-CoT<br>ScreenPrompt<br>Inclusion-favored:<br>9 include<br>1 exclude | 85.0%<br>340/400 | 98.0%<br>196/200 | 72.0%<br>144/200 |
|  | Few-shot GPT-CoT<br>ScreenPrompt<br>Exclusion-favored:<br>1 include<br>9 exclude | 83.8%<br>335/400 | **98.5%**<br>**197/200** | 69.0%<br>138/200 |

**Supplementary Table 6:** Comparative analysis of Abstract ScreenPrompt across LLM models.

| Dataset | Prompting Strategy | Accuracy | Sensitivity | Specificity | Cost (CAD) |
| --- | --- | --- | --- | --- | --- |
| ST Training<br>n=200 include<br>n=200 exclude | GPT4-0125-preview | **94.3%** | **94.5%** | 94.0% | \$12.54 |
|  | Abstract ScreenPrompt | **377/400** | **189/200** | 188/200 |  |
| | GPT4-Turbo-0409 | 89.8% | 83.5% | 96.0% | \$12.16 |
|  | Abstract ScreenPrompt | 359/400 | 167/200 | 192/200 |  |
| | GPT3.5 | 66.7% | 90.5% | 43.0% | \$0.37 |
|  | Abstract ScreenPrompt | 266/399 | 180/199 | 86/200 |  |
|  | Gemini Pro | 76.2% | 67.5% | 84.8% | **Free** |
|  | Abstract ScreenPrompt | 301/395 | 133/197 | 168/198 |  |
| | Mistral-Large | 89.3% | 93.5% | 85.0% | \$2.65 |
|  | Abstract ScreenPrompt | 357/400 | 187/200 | 170/200 |  |
| | Mixtral-8x22 | 84.5% | 94.0% | 74.9% | \$4.15 |
|  | Abstract ScreenPrompt | 337/399 | 188/200 | 149/199 |  |
| | GPT4o-0513 | 89.0% | 79.5% | **98.5%** | \$7.37 |
|  | Abstract ScreenPrompt | 356/400 | 159/200 | **197/200** |  |

**Supplementary Table 7.** Zero-shot vs. Abstract ScreenPrompt across LLM Models.

| Dataset | Model | Prompt | Accuracy | Sensitivity | Specificity |
| --- | --- | --- | --- | --- | --- |
| Training<br>n=200 include<br>n=200 exclude | GPT4-0125-preview | Abstract<br>ScreenPrompt | **94.3%**<br>**377/400** | **94.5%**<br>**189/200** | 94.0%<br>188/200 |
|  |  | Zero-shot | 65.0%<br>260/400 | 30.0%<br>60/200 | **100.0%**<br>**200/200** |
|  | GPT4-Turbo-0409 | Abstract<br>ScreenPrompt | **89.8%**<br>**359/400** | **83.5%**<br>**167/200** | 96.0%<br>192/200 |
|  |  | Zero-shot | 56.8%<br>227/400 | 13.5%<br>27/200 | **100.0%**<br>**200/200** |
|  | GPT3.5 | Abstract<br>ScreenPrompt | **66.7%**<br>**266/399** | 90.5%<br>180/199 | **43.0%**<br>**86/200** |
|  |  | Zero-shot | 64.5%<br>258/400 | **97.0%**<br>**193/200** | 32.5%<br>65/200 |

**Supplementary Table 8.** Generalizability of Abstract ScreenPrompt across SRs.

| Dataset | # of Abstracts<br>n = total<br>(include/<br>exclude) | Prompt | Accuracy | Sensitivity | Specificity |
| --- | --- | --- | --- | --- | --- |
| SeroTracker<br>(Test Dataset) | 400<br>(200/200) | Zero-shot | 71.5%<br>286/400 | 43.0%<br>86/200 | 100.0%<br>200/200 |
|  |  | Abstract<br>ScreenPrompt | 93.3%<br>373/400 | 95.5%<br>191/200 | 91.0%<br>182/200 |
| Reinfection | 400<br>(181/219) | Zero-shot | 86.9%<br>345/397 | 87.6%<br>156/178 | 86.3%<br>189/219 |
|  |  | Abstract<br>ScreenPrompt | 79.5%<br>318/400 | 97.8%<br>177/181 | 64.4%<br>141/219 |
| PA-Testing | 400<br>(52/348) | Zero-shot | 95.0%<br>380/400 | 80.8%<br>42/52 | 97.1%<br>338/348 |
|  |  | Abstract<br>ScreenPrompt | 91.3%<br>365/400 | 98.1%<br>51/52 | 90.2%<br>314/348 |
| PA-Outcomes | 400<br>(16/384) | Zero-shot | 94.5%<br>378/400 | 87.5%<br>14/16 | 94.8%<br>364/384 |
|  |  | Abstract<br>ScreenPrompt | 73.5%<br>294/400 | 100.0%<br>16/16 | 72.4%<br>278/384 |
| Meds-HA | 400<br>(140/260) | Zero-shot | 71.8%<br>287/400 | 20.0%<br>28/140 | 99.6%<br>259/260 |
|  |  | Abstract<br>ScreenPrompt | 90.3%<br>361/400 | 93.6%<br>131/140 | 88.5%<br>230/260 |
| Sepsis | 400<br>(17/383) | Zero-shot | 97.8%<br>391/400 | 52.9%<br>9/17 | 99.7%<br>382/383 |
|  |  | Abstract<br>ScreenPrompt | 92.5%<br>370/400 | 100.0%<br>17/17 | 92.2%<br>353/383 |
| Spinal | 400<br>(135/265) | Zero-shot | 88.5%<br>354/400 | 70.4%<br>95/135 | 97.7%<br>259/265 |
|  |  | Abstract<br>ScreenPrompt | 87.5%<br>350/400 | 100.0%<br>135/135 | 81.1%<br>215/265 |
| Infant-NO | 400<br>(6/394) | Zero-shot | 98.0%<br>392/400 | 16.7%<br>1/6 | 99.2%<br>391/394 |
|  |  | Abstract<br>ScreenPrompt | 90.8%<br>363/400 | 100.0%<br>6/6 | 90.6%<br>357/394 |
| Calcium-HA | 400<br>(15/385) | Zero-shot | 97.5%<br>390/400 | 40.0%<br>6/15 | 99.7%<br>384/385 |
|  |  | Abstract<br>ScreenPrompt | 95.5%<br>382/400 | 86.7%<br>13/15 | 95.8%<br>369/385 |
| SVCF | 400<br>(17/383) | Zero-shot | 97.3%<br>389/400 | 35.3%<br>6/17 | 100.0%<br>383/383 |
|  |  | Abstract<br>ScreenPrompt | 91.3%<br>365/400 | 100.0%<br>17/17 | 90.9%<br>348/383 |

**Supplementary Table 9.** Abstract ScreenPrompt Self-Consistency Analysis.

| Dataset | Method | Accuracy | Sensitivity | Specificity | Cost (USD) |
| --- | --- | --- | --- | --- | --- |
| SeroTracker | Abstract ScreenPrompt | 93.3%<br>373/400 | 95.5%<br>191/200 | 91.0%<br>182/200 | **\$12.56** |
| | Abstract ScreenPrompt + Self-Consistency | **95.9%**<br>**383/399** | **97.5%**<br>**195/200** | **94.5%**<br>**188/199** | \$88.02<br>Batch API |
| Reinfection | Abstract ScreenPrompt | 79.5%<br>318/400 | 97.8%<br>177/181 | 64.4%<br>141/219 | **\$14.77** |
| | Abstract ScreenPrompt + Self-Consistency | **81.4%**<br>**323/397** | **99.4%**<br>**180/181** | **66.2%**<br>**143/216** | \$89.18<br>Batch API |

**Supplementary Table 10.** Full-text prompt engineering performance.

| Dataset | Prompting Strategy | Accuracy | Sensitivity | Specificity |
| --- | --- | --- | --- | --- |
| Training<br>(n=200 include,<br>n=200 exclude) | Zero-shot | 68.9%<br>275/399 | 37.7 %<br>75/199 | 100.0%<br>200/200 |
|  | Abstract<br>ScreenPrompt | 85.5%<br>342/400 | **98.0%**<br>**196/200** | 73.0%<br>146/200 |
|  | ScreenPrompt | 91.3%<br>365/400 | 94.5%<br>189/200 | 88.0%<br>176/200 |
|  | ScreenPrompt<br>Init + Fin | 94.8%<br>379/400 | 95.0%<br>190/200 | 94.5%<br>189/200 |
|  | Numbered<br>ScreenPrompt | 95.3%<br>381/400 | 96.0%<br>192/200 | 94.5%<br>189/200 |
|  | **ISO-ScreenPrompt** | **95.5%**<br>**382/400** | 93.5%<br>187/200 | **97.5%**<br>**195/200** |
| Validation<br>(n=200 include,<br>n=200 exclude) | ISO-ScreenPrompt | 96.3%<br>385/400 | 97.5%<br>195/200 | 95%<br>190/200 |

**Supplementary Table 11.** : Full-text Prompt Structure Testing.

| Dataset | Prompting Strategy<br>(Structure) | Accuracy | Sensitivity | Specificity |
| --- | --- | --- | --- | --- |
| Training<br>(n=200 include,<br>n=200 exclude) | ScreenPrompt | 91.3%<br>365/400 | 94.5%<br>189/200 | 88.0%<br>176/200 |
|  | ScreenPrompt<br>Init | 92.5%<br>370/400 | 94.5%<br>189/200 | 90.5%<br>181/200 |
|  | ScreenPrompt<br>Fin | 82.2%<br>328/399 | 98.0%<br>196/200 | 66.3%<br>132/199 |
|  | ScreenPrompt<br>Init + Fin-Instructions | 93.3%<br>373/400 | 92.5%<br>185/200 | 94.0%<br>188/200 |
|  | ScreenPrompt<br>Init + Fin | 94.8%<br>379/400 | 95.0%<br>190/200 | 94.5%<br>189/200 |

**Supplementary Table 12.** : Exemplar Wrong Outputs during Full-text Prompt Engineering.

| Prompt | Example LLM Output | Author feedback |
| --- | --- | --- |
| ScreenPrompt<br>Init | XXX | Single token output<br>→ Led to experiments testing adding instructions and whole prompts at the bottom of the prompt |
| ScreenPrompt<br>Init + Fin-Instructions | ### Decision Process for Inclusion or Exclusion<br>To assess the paper's suitability for inclusion in the systematic review, we must methodically examine it against each criterion listed under the inclusion and exclusion categories. This approach ensures that the decision is objective, based on predefined standards and takes the study's characteristics into account. Let's delve into each criterion, evaluating how the paper stands against them.<br>### Inclusion Criteria<br>1. **Population:** The study focuses on humans of any age across districts in India, which meets the first inclusion criterion.<br>2. **Study design:** The paper appears to employ a cross-sectional analysis within a defined population over a certain period, aiming to correlate BMI with the severity and spatial variation of COVID-19 in India. This aligns with the inclusion criterion related to study designs accepted.<br>3. **Special design:** While not a systematic review or meta-analysis itself, the paper's systematic approach to evidence synthesis can indirectly contribute to understanding seroprevalence studies' landscape.<br>4. **Sampling:** The paper does not detail its sampling method distinctly, but its analysis based on district-level data suggests an organized systematic approach, likely consistent with acceptable sampling methods for inclusion.<br>5. **Types of evidence:** The manuscript is an academic literature piece. Even though it does not mention its publication status, given the context, it's assumed to meet the inclusion criteria related to evidence types.<br>6. **Outcome measures:** <u>The paper reports on several outcome measures, including confirmed cases and severity indicators of COVID-19, which indirectly inform on the pandemic's spread, though not seroprevalence estimates directly.</u><br>### Exclusion Criteria<br>1. **Population:** The study solely includes human subjects, thus not meeting any exclusion criteria related | Inclusion/exclusion criteria evaluated by meta-criteria headings, reducing important granularity.<br>For example, seroprevalence was not reported, but the LLM inferred that outcome measures were correctly reported, thus deeming the criteria (**incorrectly**) satisfied<br>→ Led us to enhance our instruction format and model outputs with 'Numbered framework CoT' |
|  | to the population.<br>2. **\*\*Study design:\*\*** The paper embodies a cross-sectional study design, thus not falling under any excluded designs like case reports or evaluations of serological tests.<br>3. **\*\*Types of evidence:\*\*** The manuscript does not rely on multimedia, dashboards unrelated to a defined serology study, or unattributable slide deck presentations, thereby not meeting the exclusion criteria regarding evidence types.<br>4. **\*\*Outcome measures:\*\*** Although the paper does not directly report seroprevalence estimates, its focus on COVID-19's intensity and spatial variation based on BMI associations indirectly contributes to understanding the pandemic's spread. Hence, it doesn't directly fulfill any of the specific outcome measure exclusion criteria.<br>### Conclusion<br>Considering the analysis above, the paper does not fit neatly into the exclusion criteria and aligns with several critical inclusion criteria, notably around population, study design, and types of evidence. Despite its focus not being directly on seroprevalence estimates, its insight into the pandemic's spread through indirect measures like BMI's impact offers valuable, albeit tangential, contributions to seroprevalence discussions. The decision, thus, veers towards inclusion based on the information provided and the indirect relevance of its findings to understanding aspects of the pandemic's spread, which is of interest in the broader context of seroprevalence studies.<br>YYY |  |
| Numbered ScreenPrompt | ### Decision-Making Process<br>To decide whether to include or exclude the abstract, we'll evaluate it against the inclusion and exclusion criteria systematically.<br>### Inclusion Criteria Assessment<br>1. **\*\*Humans of any age.\*\*** [Met]<br>- The study discusses the impacts of COVID-19 policies on the Swedish population, implying human subjects of unspecified ages.<br>2. **\*\*Including COVID-19 antigen positive persons and those with suspected disease if not deliberately sampled.\*\*** [Not Met]<br>- The abstract focuses on policy outcomes and pandemic metrics rather than sampling individuals based on antigen positivity or suspected disease.<br>3. **\*\*Sero-surveys.\*\*** [Not Met]<br>- This study does not mention sero-surveys or the collection and testing of serum to estimate SARS-CoV-2 antibody prevalence.<br>4. **\*\*Study Designs.\*\*** [Not Met]<br>- The study follows a longitudinal observational design, evaluating pandemic variables over time but does not fit | Model now correctly evaluates each individual inclusion/exclusion sub-criterion |
|  | the specified designs focused on serology measurements.<br>5. **\*\*Include systematic reviews and meta-analysis of seroprevalence studies.\*\*** [Not Met]<br>- The study does not review or meta-analyze seroprevalence studies; it examines policy outcomes and pandemic data.<br>6. **\*\*Any sampling method.\*\*** [Not Met]<br>- Since it doesn't involve direct sero-surveys, it doesn't specify a sampling method for such purposes.<br>7. **\*\*Literature type.\*\*** [Met]<br>- The abstract seems part of academic literature, which is eligible.<br>8. **\*\*Reports a seroprevalence estimate.\*\*** [Not Met]<br>- No seroprevalence or proportion of the population with antibodies is reported or estimated.<br>9. **\*\*Reports the number of participants enrolled (denominator).\*\*** [Not Met]<br>- The study does not involve enrolling participants in the traditional sense nor reports such a denominator.<br>10. **\*\*Reports study sampling end date/week.\*\*** [Met]<br>- Provides detailed timelines regarding the pandemic data collection period.<br>11. **\*\*Reports the locations at which the study takes place.\*\*** [Met]<br>- The study clearly takes place in Sweden, satisfying the geographical reporting requirement.<br>### Exclusion Criteria Assessment<br>1. **\*\*Non-human studies.\*\*** [Not Met]<br>- The study involves human data.<br>2. **\*\*Study only included individuals with confirmed COVID-19 cases.\*\*** [Not Applicable]<br>- Does not exclusively focus on individuals with confirmed or suspected COVID-19 cases in a clinical assessment sense but evaluates general pandemic outcomes.<br>3. **\*\*Study only included vaccinated individuals.\*\*** [Not Applicable]<br>- Vaccination status is not the focus.<br>4. **\*\*Study designs other than specified.\*\*** [Met]<br>- The study does not adhere to the specified cross-sectional, cohort, or serology measurement designs.<br>5. **\*\*Multimedia sources or unverifiable slides.\*\*** [Not Met]<br>- The abstract is presented as part of academic literature.<br>6. **\*\*Dashboards not associated with a defined serology study.\*\*** [Not Applicable]<br>- The study is not about a dashboard per se but uses data from national health registries.<br>7. **\*\*Only reports incidence or prevalence of antigen.\*\*** [Not Met]<br>- The study reports on broader pandemic variables without focusing solely on antigen incidence or prevalence.<br>8. **\*\*Does not report sampling end date/week.\*\*** [Not |  |
|  | Met]<br>- It reports the period of data collection.<br>9. **Does not report the number of participants.** [Met]<br>- As it is not a sero-survey-based study, it does not report on specific participant numbers in that context.<br>10. **Does not report the location.** [Not Met]<br>- Location is reported.<br>### Conclusion<br>The paper is predominantly focused on evaluating the effects of policy measures and their outcomes in terms of general pandemic variables over time in Sweden. It lacks a direct focus on sero-surveys, seroprevalence estimates regarding SARS-CoV-2 antibodies, and does not fit the required study design for seroprevalence studies as dictated by the inclusion criteria. Therefore, the paper does not meet the inclusion criteria essential for the systematic review focused on SARS-CoV-2 antibodies prevalence, variations, and underestimation of pandemic spread through acute infection surveillance.<br>Consequently, based on the systematic evaluation: XXX |  |

**Supplementary Table 13.** : ISO-Prompting in Abstract Screening.

| Dataset | Prompting Strategy | Accuracy | Sensitivity | Specificity |
| --- | --- | --- | --- | --- |
| Training<br>(n=200 include,<br>n=200 exclude) | Abstract ScreenPrompt<br>(Regular structure) | 94.3%<br>377/400 | 94.5%<br>189/200 | 94.0%<br>188/200 |
|  | Numbered Abstract ScreenPrompt | 94.5%<br>378/400 | 91.5%<br>183/200 | 97.5%<br>195/200 |
|  | Abstract ScreenPrompt<br>Init | 95.0%<br>380/400 | 93.5%<br>187/200 | 96.5%<br>193/200 |
|  | Abstract ScreenPrompt<br>Init + Fin | 95.3%<br>381/400 | 94.0%<br>188/200 | 96.5%<br>193/200 |

**Supplementary Table 14.**
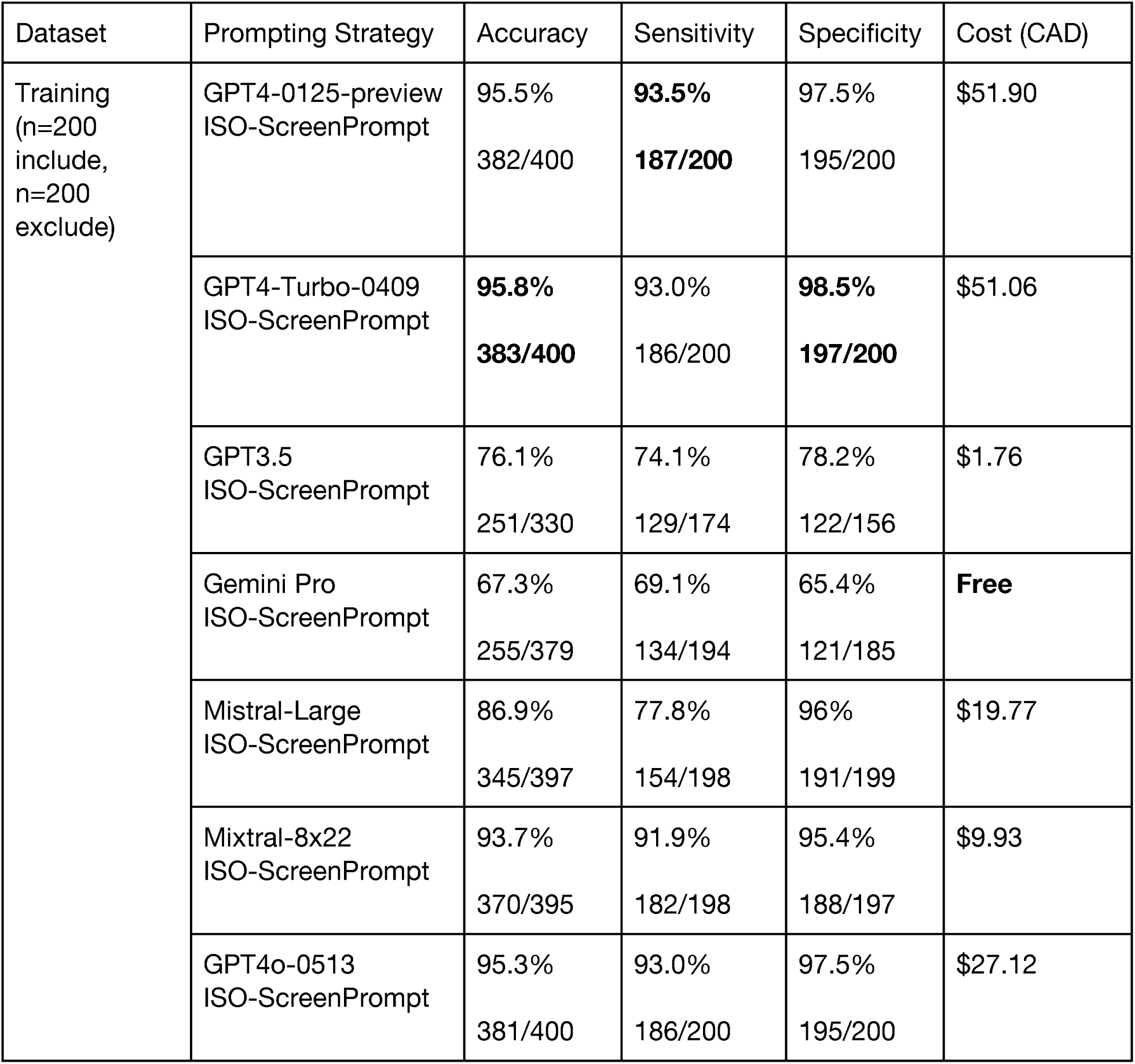
Comparative analysis of ISO-ScreenPrompt across LLM models.

| Dataset | Prompting Strategy | Accuracy | Sensitivity | Specificity | Cost (CAD) |
| --- | --- | --- | --- | --- | --- |
| Training<br>(n=200<br>include,<br>n=200<br>exclude) | GPT4-0125-preview<br>ISO-ScreenPrompt | 95.5%<br>382/400 | **93.5%**<br>**187/200** | 97.5%<br>195/200 | \$51.90 |
| | GPT4-Turbo-0409<br>ISO-ScreenPrompt | **95.8%**<br>**383/400** | 93.0%<br>186/200 | **98.5%**<br>**197/200** | \$51.06 |
| | GPT3.5<br>ISO-ScreenPrompt | 76.1%<br>251/330 | 74.1%<br>129/174 | 78.2%<br>122/156 | \$1.76 |
|  | Gemini Pro<br>ISO-ScreenPrompt | 67.3%<br>255/379 | 69.1%<br>134/194 | 65.4%<br>121/185 | **Free** |
| | Mistral-Large<br>ISO-ScreenPrompt | 86.9%<br>345/397 | 77.8%<br>154/198 | 96%<br>191/199 | \$19.77 |
| | Mixtral-8x22<br>ISO-ScreenPrompt | 93.7%<br>370/395 | 91.9%<br>182/198 | 95.4%<br>188/197 | \$9.93 |
| | GPT4o-0513<br>ISO-ScreenPrompt | 95.3%<br>381/400 | 93.0%<br>186/200 | 97.5%<br>195/200 | \$27.12 |

**Supplementary Table 15.** Zero-shot vs. ISO-ScreenPrompt across LLM Models.

| Dataset | Model | Prompt | Accuracy | Sensitivity | Specificity |
| --- | --- | --- | --- | --- | --- |
| Training<br>(n=200<br>include,<br>n=200<br>exclude) | GPT4-0125-preview | **ISO-ScreenPrompt** | **95.5%**<br>**382/400** | **93.5%**<br>**187/200** | 97.5%<br>195/200 |
|  |  | Zero-shot | 68.9%<br>275/399 | 37.7%<br>75/199 | **100.0%**<br>**200/200** |
|  | GPT4-Turbo-0409 | **ISO-ScreenPrompt** | **95.8%**<br>**383/400** | **93.0%**<br>**186/200** | 98.5%<br>197/200 |
|  |  | Zero-shot | 56.8%<br>227/400 | 13.5%<br>27/200 | **100.0%**<br>**200/200** |
|  | GPT3.5 | **ISO-ScreenPrompt** | 76.1%<br>251/330 | 74.1%<br>129/174 | **78.2%**<br>**122/156** |
|  |  | Zero-shot | 72.1%<br>253/351 | **92.8%**<br>**168/181** | 50.0%<br>85/170 |

**Supplementary Table 16.** Generalizability of ISO-ScreenPrompt across SRs.

| Dataset | # of full-text<br>n = total<br>(include/<br>exclude) | Prompt | Accuracy | Sensitivity | Specificity |
| --- | --- | --- | --- | --- | --- |
| SeroTracker SR<br>(Test Dataset) | 400<br>(200/200) | Zero-shot | 73.9%<br>294/398 | 48.0%<br>95/198 | 99.5%<br>199/200 |
|  |  | ISO-ScreenPrompt | 96.5%<br>384/398 | 98.5%<br>195/198 | 94.5%<br>189/200 |
| Reinfection SR | 400<br>(141/255) | Zero-shot | 85.0%<br>340/400 | 71.0%<br>103/145 | 92.9%<br>237/255 |
|  |  | ISO-ScreenPrompt | 83.3%<br>333/400 | 89.7%<br>130/145 | 79.6%<br>203/255 |
| PA-Testing SR | 400<br>(45/355) | Zero-shot | 97.3%<br>389/400 | 88.9%<br>40/45 | 98.3%<br>349/355 |
|  |  | ISO-ScreenPrompt | 93.2%<br>372/399 | 100.0%<br>45/45 | 92.4%<br>327/354 |
| PA-Outcomes | 400<br>(16/384) | Zero-shot | 90.8%<br>363/400 | 93.8%<br>15/16 | 90.6%<br>348/384 |
|  |  | ISO-ScreenPrompt | 84.3%<br>337/400 | 100.0%<br>16/16 | 83.6%<br>321/384 |
| Meds-HA SR | 218<br>(17/200) | Zero-shot | 95.5%<br>382/400 | 11.8%<br>2/17 | 99.2%<br>380/383 |
|  |  | ISO-ScreenPrompt | 95.8%<br>383/400 | 100.0%<br>17/17 | 95.6%<br>366/383 |
| Sepsis SR | 400<br>(16/384) | Zero-shot | 99.0%<br>383/387 | 75.0%<br>12/16 | 100.0%<br>371/371 |
|  |  | ISO-ScreenPrompt | 95.3%<br>369/387 | 93.8%<br>15/16 | 95.4%<br>354/371 |
| Spinal SR | 244<br>(13/231) | Zero-shot | 96.7%<br>236/244 | 84.6%<br>11/13 | 97.4%<br>225/231 |
|  |  | ISO-ScreenPrompt | 90.2%<br>220/244 | 100.0%<br>13/13 | 89.6%<br>207/231 |
| Infant-NO | 197<br>(6/191) | Zero-shot | 98.5%<br>194/197 | 66.7%<br>4/6 | 99.5%<br>190/191 |
|  |  | ISO-ScreenPrompt | 98.0%<br>193/197 | 100.0%<br>6/6 | 97.9%<br>187/191 |
| Calcium-HA | 238<br>(14/224) | Zero-shot | 97.5%<br>232/238 | 57.1%<br>8/14 | 100.0%<br>224/224 |
|  |  | ISO-ScreenPrompt | 99.6%<br>237/238 | 92.9%<br>13/14 | 100.0%<br>224/224 |
| SCVF | 167<br>(17/150) | Zero-shot | 95.8%<br>160/167 | 58.8%<br>10/17 | 100.0%<br>150/150 |
|  |  | ISO-ScreenPrompt | 99.4%<br>166/167 | 100.0%<br>17/17 | 99.3%<br>149/150 |

**Supplementary Table 17.**
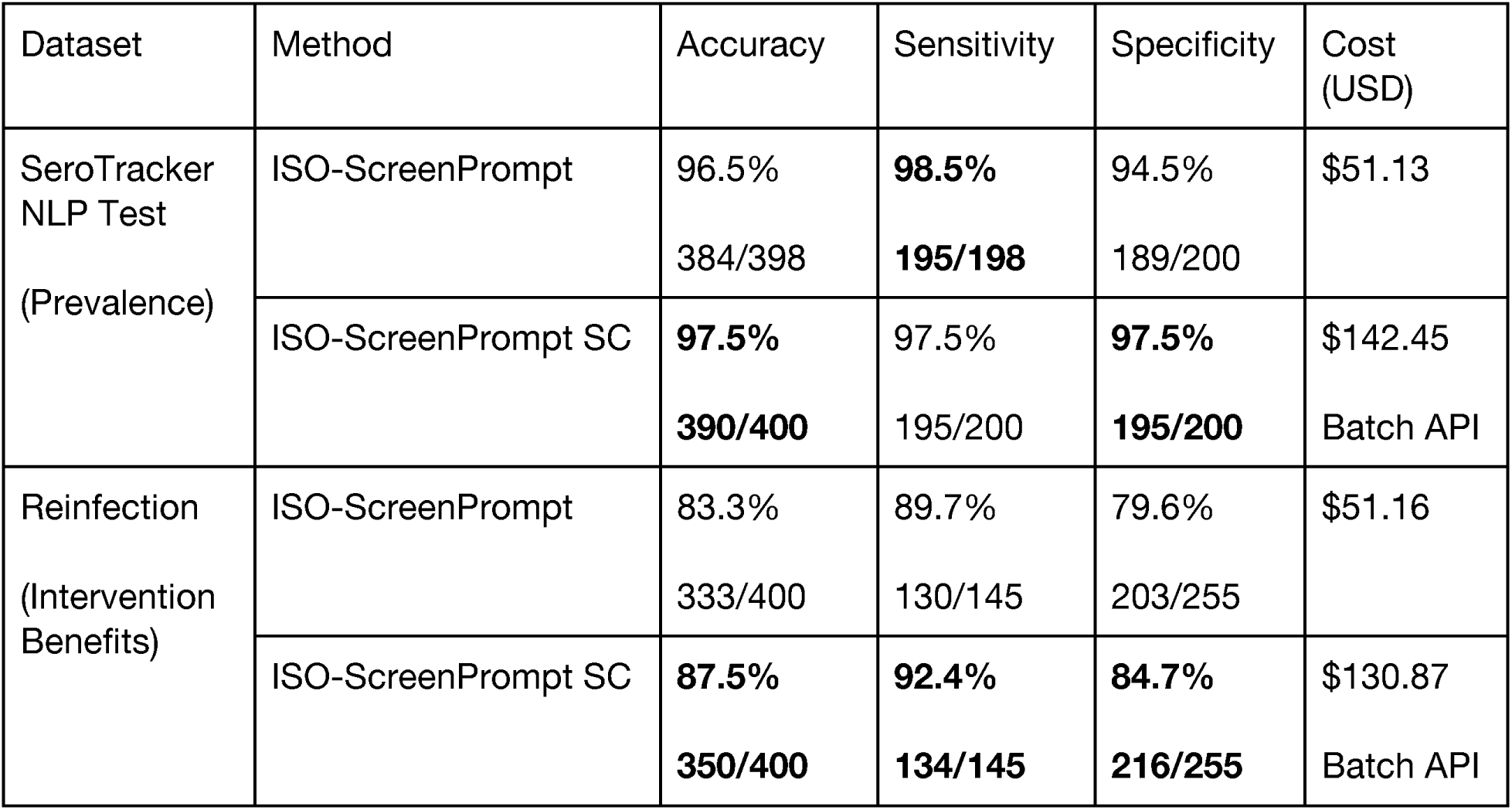
ISO-ScreenPrompt Self-Consistency Analysis.

**Supplementary Table 18.**
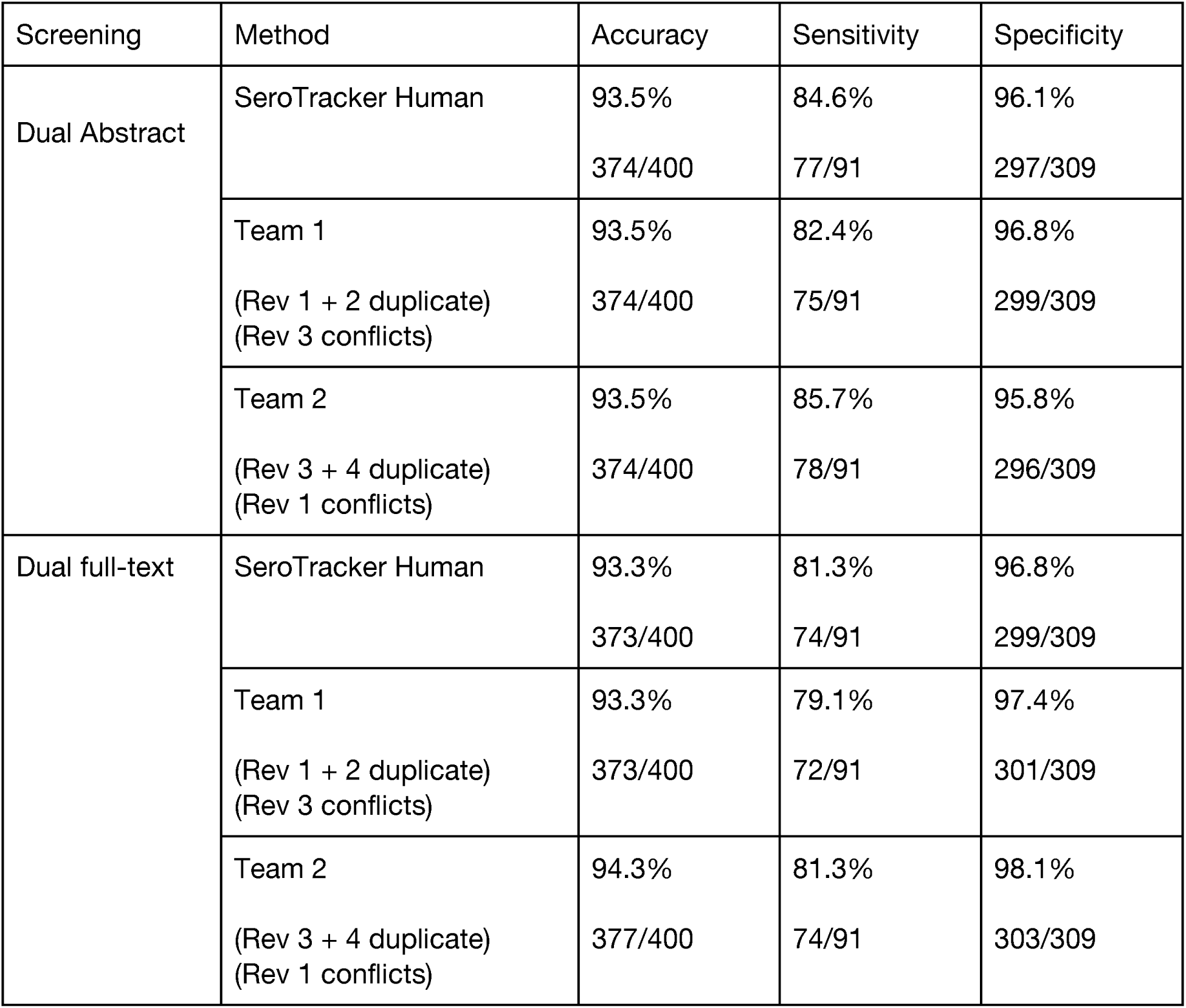
Human Screening Calibration.

**Supplementary Table 19.** Abstract ScreenPrompt vs Human Dual Abstract Screening.

| Dataset | Method | Accuracy | Sensitivity | Specificity |
| --- | --- | --- | --- | --- |
| SeroTracker NLP Test<br>(Prevalence) | ISO-ScreenPrompt | 93.0%<br>372/400 | **96.7%**<br>**88/91** | 91.9%<br>284/309 |
|  | Dual Human<br>(SeroTracker Human) | **93.5%**<br>**374/400** | 84.6%<br>77/91 | **96.1%**<br>297/309 |
| Reinfection<br>(Intervention Benefits) | ISO-ScreenPrompt | 78.3%<br>311/397 | **95.8%**<br>**136/142** | 68.6%<br>175/255 |
|  | Dual Human | **89.5%**<br>**358/400** | 84.1%<br>122/145 | **92.5%**<br>**236/255** |
| PA-Outcomes<br>(Intervention Harms) | ISO-ScreenPrompt | 78.9%<br>310/393 | 100.0%<br>16/16 | 78.0%<br>294/377 |
|  | Dual Human | **94.8%**<br>**379/400** | 100.0%<br>16/16 | **94.5%**<br>**363/384** |
| PA-Testing<br>(Diagnostic Test Accuracy) | ISO-ScreenPrompt | 94.0%<br>375/399 | **100.0%**<br>**44/44** | 93.2%<br>331/355 |
|  | Dual Human | **97.8%**<br>**391/400** | 97.8%<br>44/45 | **97.7%**<br>**347/355** |
| SVCF<br>(Prognosis) | ISO-ScreenPrompt | 94.4%<br>152/161 | **100.0%**<br>**17/17** | 93.8%<br>135/144 |
|  | Dual Human | **97.6%**<br>**163/167** | 88.2%<br>15/17 | **98.7%**<br>**148/150** |

**Supplementary Table 20.**
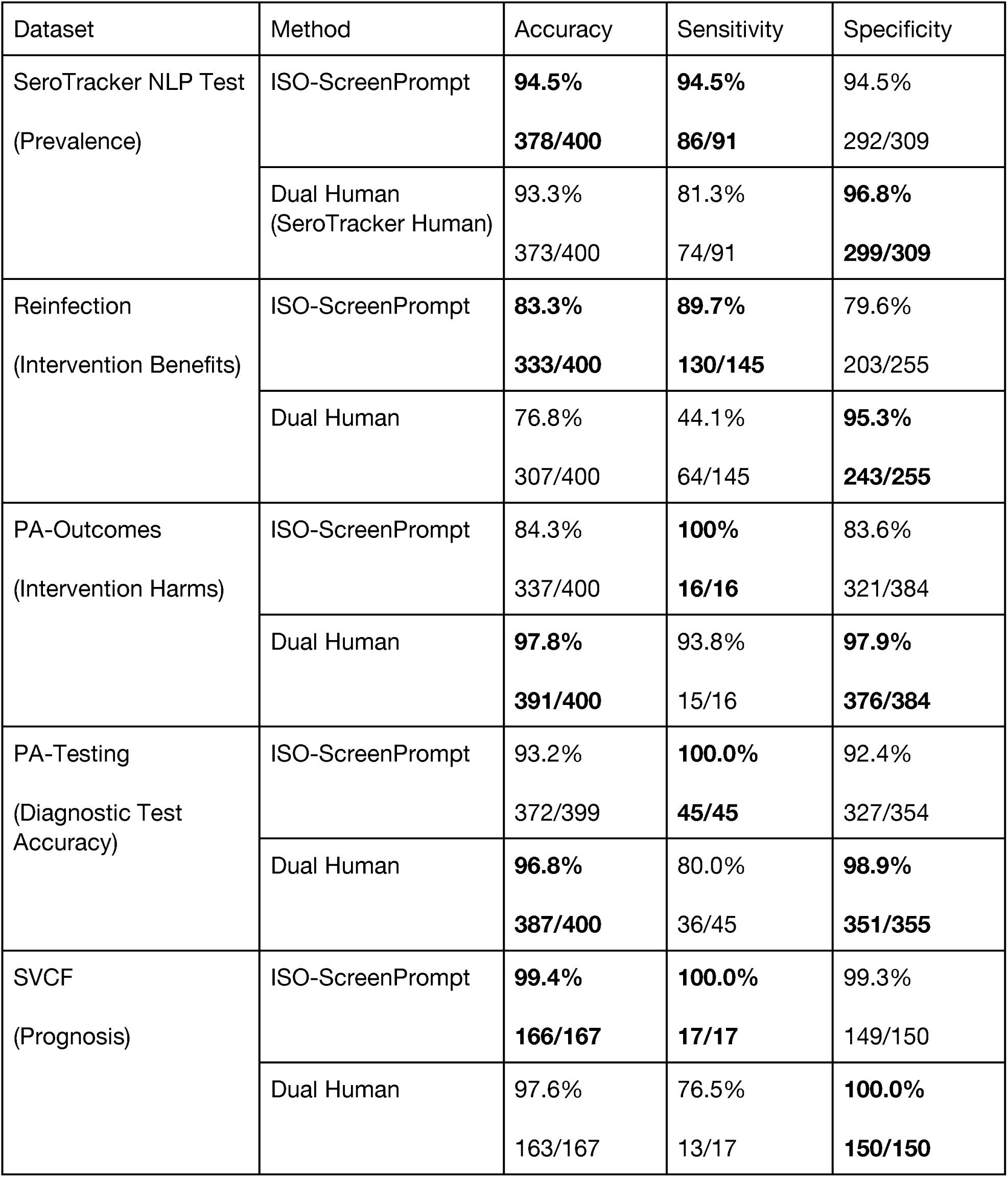
ISO-ScreenPrompt vs Full Dual Screening.

| Dataset | Method | Accuracy | Sensitivity | Specificity |
| --- | --- | --- | --- | --- |
| SeroTracker NLP Test<br>(Prevalence) | ISO-ScreenPrompt | **94.5%**<br>**378/400** | **94.5%**<br>**86/91** | 94.5%<br>292/309 |
|  | Dual Human<br>(SeroTracker Human) | 93.3%<br>373/400 | 81.3%<br>74/91 | **96.8%**<br>**299/309** |
| Reinfection<br>(Intervention Benefits) | ISO-ScreenPrompt | **83.3%**<br>**333/400** | **89.7%**<br>**130/145** | 79.6%<br>203/255 |
|  | Dual Human | 76.8%<br>307/400 | 44.1%<br>64/145 | **95.3%**<br>**243/255** |
| PA-Outcomes<br>(Intervention Harms) | ISO-ScreenPrompt | 84.3%<br>337/400 | **100%**<br>**16/16** | 83.6%<br>321/384 |
|  | Dual Human | **97.8%**<br>**391/400** | 93.8%<br>15/16 | **97.9%**<br>**376/384** |
| PA-Testing<br>(Diagnostic Test Accuracy) | ISO-ScreenPrompt | 93.2%<br>372/399 | **100.0%**<br>**45/45** | 92.4%<br>327/354 |
|  | Dual Human | **96.8%**<br>**387/400** | 80.0%<br>36/45 | **98.9%**<br>**351/355** |
| SVCF<br>(Prognosis) | ISO-ScreenPrompt | **99.4%**<br>**166/167** | **100.0%**<br>**17/17** | 99.3%<br>149/150 |
|  | Dual Human | 97.6%<br>163/167 | 76.5%<br>13/17 | **100.0%**<br>**150/150** |

**Supplementary Table 21.**
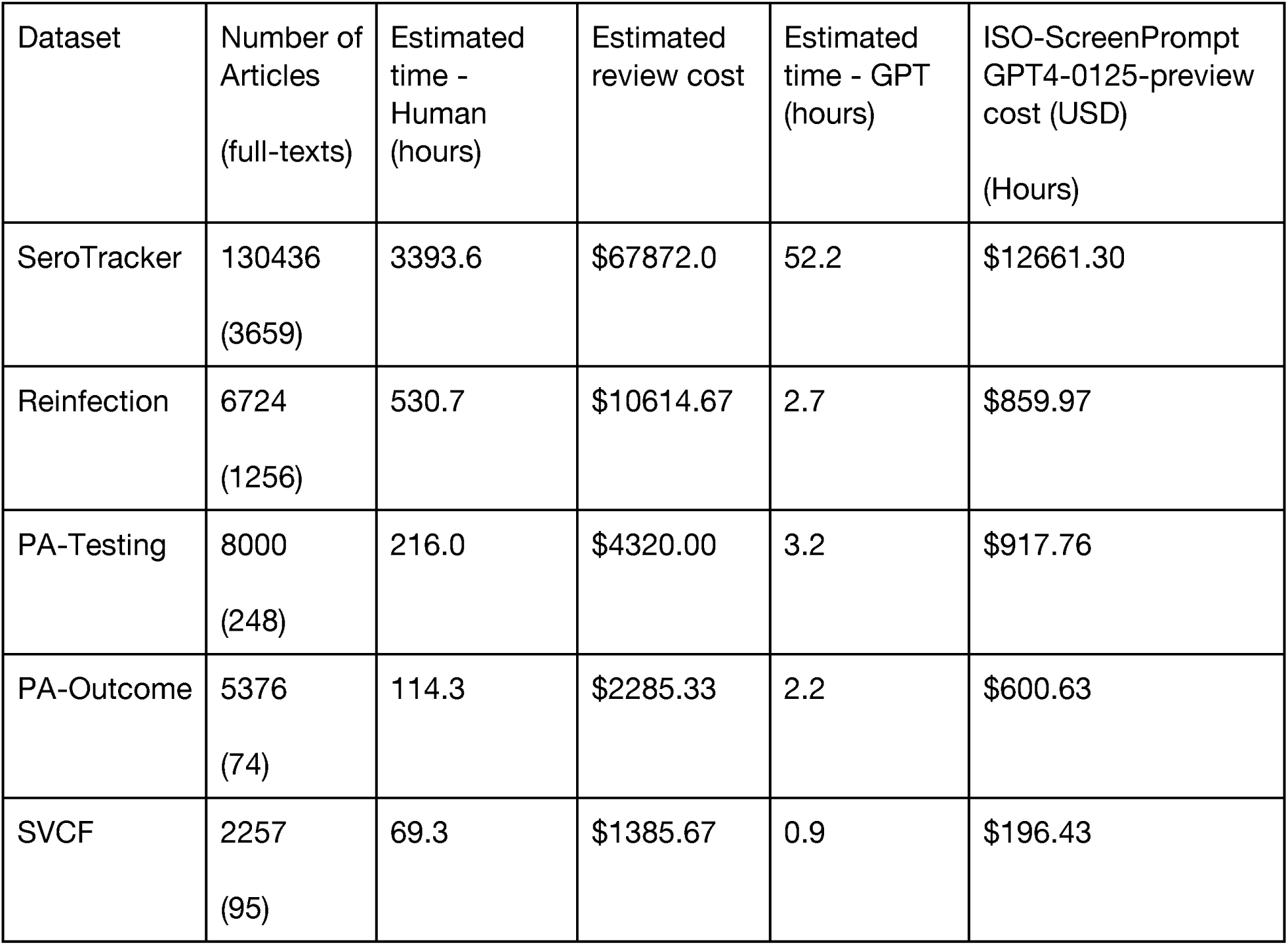
Time and Cost Savings Analysis for ISO-ScreenPrompt and Full-dual Screening.

| Dataset | Number of Articles<br>(full-texts) | Estimated time - Human<br>(hours) | Estimated review cost | Estimated time - GPT<br>(hours) | ISO-ScreenPrompt GPT4-0125-preview cost (USD)<br>(Hours) |
| --- | --- | --- | --- | --- | --- |
| SeroTracker | 130436<br>(3659) | 3393.6 | \$67872.0 | 52.2 | \$12661.30 |
| Reinfection | 6724<br>(1256) | 530.7 | \$10614.67 | 2.7 | \$859.97 |
| PA-Testing | 8000<br>(248) | 216.0 | \$4320.00 | 3.2 | \$917.76 |
| PA-Outcome | 5376<br>(74) | 114.3 | \$2285.33 | 2.2 | \$600.63 |
| SVCF | 2257<br>(95) | 69.3 | \$1385.67 | 0.9 | \$196.43 |

**Supplementary Table 22.** Time and Cost Savings Analysis for Abstract ScreenPrompt and Single Human-Reviewer Screening.

| Dataset | Number of Articles | Estimated time - Human (hours) | Estimated review cost Single Reviewer | Estimated time - GPT (hours) | Abstract ScreenPrompt GPT4-0125- preview cost (USD) |
| --- | --- | --- | --- | --- | --- |
| SeroTracker | 130436 | 1086.97 | \$21739.33 | 15.2 | \$4122.49 |
| Reinfection | 6724 | 56.03 | \$1120.67 | 0.8 | \$244.93 |
| PA-Testing | 8000 | 66.67 | \$1333.40 | 0.9 | \$266.64 |
| PA-Outcome | 5376 | 44.80 | \$896.00 | 0.6 | \$162.55 |
| SVCF | 2257 | 18.81 | \$376.17 | 0.3 | \$55.25 |

## Supplementary Note 1

Below represents the screening documents provided to human reviewers for each selected SR. The documents contain the same study objectives, inclusion and exclusion criteria used in prompting (‘simplified objectives and inclusion/exclusion criteria’), and any additional internal protocol documents provided by the original study authors for each SR.

### SeroTracker

#### Simplified Objectives + Inclusion/Exclusion Criteria

Our systematic review is governed by the following objectives: (i) describe the global prevalence of SARS-CoV-2 antibodies based on serosurveys; (ii) detect variations in seroprevalence arising from study design and geographic factors; (iii) identify populations at high risk for SARS-CoV-2 infection; and (iv) evaluate the extent to which surveillance based on detection of acute infection underestimates the spread of the pandemic.

The following is an excerpt of two sets of criteria. A study is considered included if it meets all the inclusion criteria. If a study meets any of the exclusion criteria, it should be excluded. Here are the two sets of criteria:

##### Inclusion Criteria (all must be fulfilled)

Population

1. Humans of any age
2. Including COVID-19 antigen positive persons and those with suspected disease if not deliberately sampled.

Study design

1. Sero-surveys – defined as the collection and testing of serum (or proxy such as oral fluid) specimens from a sample of a defined population over a specified period of time to estimate the prevalence of antibodies against SARS-CoV-2 as an indicator of immunity
2. Cross-sectional, repeated cross sectional, and cohort study designs, with serology measurements at single time points or repeated at multiple time points

Special design

1. Include systematic reviews and meta-analysis of seroprevalence studies for the purpose of tracking evidence synthesis efforts

Sampling

1. Any sampling method Types of evidence
2. Published or unpublished academic literature, grey literature (government or institutional reports), or media reports. Slide deck presentations were included if we could identify the person giving the presentation and the date of the presentation

Outcome measures

1. Reports a seroprevalence estimate (proportion of the population with detectable antibodies)
2. Reports the number of participants enrolled in the study (denominator)
3. Reports study sampling end date/week
4. Reports the locations at which the study took places such that they could be categorized as neighbourhood, city, state/province/territory, or country

##### Exclusion Criteria (if any met then exclude)

Population

1. Non-human (e.g., in silico, animal, in vitro)
2. The study only included individuals with suspected, active, or previously diagnosed with COVID-19 using PCR, antigen testing, clinical assessment, or self-assessment
3. The study only included individuals vaccinated against SARS-CoV2 Study design
4. Study designs other than cross-sectional or cohort design: case reports, case-control studies, evaluations of serological tests, study protocols

Types of evidence

1. Multimedia sources of data (audio clips, video clips) were excluded due to the feasibility of extracting. Slide deck presentations were excluded if we could not identify the person giving the presentation and the date of the presentation
2. Dashboards not associated with a defined serology study Outcome measures
3. Only reports incidence or prevalence of SARS-CoV-2 antigen (as opposed to antibody) 2. Does not report study sampling end date/week
4. Does not report the number of participants included in the study (sample denominator) 4. Does not report the location at which the study took place

### Original author protocol

### Criteria for including evidence (must meet all the criteria to be included)

| Characteristics | Criteria for inclusion |
| --- | --- |
| Population | · Humans of any age<br>o Including COVID-19 antigen positive persons and those with suspected disease if not deliberately sampled. |
| Study design | · Sero-surveys – defined as the collection and testing of serum (or proxy such as oral fluid) specimens from a sample of a defined population over a specified period of time to estimate the prevalence of antibodies against SARS-CoV-2 as an indicator of immunity<br>· Cross-sectional, repeated cross sectional, and cohort study designs, with serology measurements at single time points or repeated at multiple time points |
| *Special design | · <u>Include</u> systematic reviews and meta-analysis of seroprevalence studies for the purpose of tracking evidence synthesis efforts |
| Sampling | · Any sampling method |
| Types of evidence | · Published or unpublished academic literature, grey literature (government or institutional reports), or media reports. Slide deck presentations were included if we could identify the person giving the presentation and the date of the presentation |
| Outcome measures | · Reports a seroprevalence estimate (proportion of the population with detectable antibodies)<br>· Reports the number of participants enrolled in the study (denominator)<br>· Reports study sampling end date/week<br>· Reports the locations at which the study took places such that they could be categorized as neighbourhood, city, state/province/territory, or country |
| Languages | · Any |

### Criteria for excluding evidence (if any met then exclude)

| Characteristics | Criteria for exclusion |
| --- | --- |
| Population | · Non-human (e.g., *in silico*, animal, *in vitro*)<br>· The study <u>only</u> included individuals with suspected, active, or previously diagnosed with COVID-19 using PCR, antigen testing, clinical assessment, or self-assessment<br>· The study <u>only</u> included individuals vaccinated against SARS-CoV2 |
| Study design | · Study designs other than cross-sectional or cohort design: case reports, case-control studies, evaluations of serological tests, study protocols |
| Sampling | · N/A |
| Types of evidence | · Multimedia sources of data (audio clips, video clips) were excluded due to the feasibility of extracting. Slide deck presentations were excluded if we could not identify the person giving the presentation and the date of the presentation<br>· Dashboards not associated with a defined serology study |
| Outcome measures | · Only reports incidence or prevalence of SARS-CoV-2 antigen (as opposed to antibody)<br>· Does not report study sampling end date/week<br>· Does not report the number of participants included in the study (sample denominator)<br>· Does not report the location at which the study took place |
| Language | · N/A |

### Reinfection

#### Simplified Objectives + Inclusion/Exclusion Criteria

We aimed to systematically review the evidence for the magnitude and duration of the effectiveness of (i) previous infection and (ii) hybrid immunity against multiple clinical outcomes of SARS-CoV-2 infection caused by the omicron variant. We also aimed to examine the comparative protection of hybrid immunity relative to previous infection only, vaccination only, and hybrid immunity with fewer vaccine doses.

The following is an excerpt of two sets of criteria. A study is considered included if it meets all the inclusion criteria. If a study meets any of the exclusion criteria, it should be excluded. Here are the two sets of criteria:

##### Inclusion Criteria (all must be fulfilled)

Population

1. Humans of any age, in any geographical setting. Exposure Group
2. Confirmed case of SARS-CoV-2 infection with or without COVID-19 vaccination.
3. SARS-CoV-2 infection will be defined as a confirmed case according to the following criteria, adapted from WHO case definitions (positive nucleic acid amplification test (NAAT) according to laboratory records or self report, positive SARS-CoV-2 antigen rapid diagnostic test (AgRDT)^a^ with high accuracy according to laboratory records or self report, or a positive serology test from a lab-based assay (i.e. CLIA/ELISA) or an antibody-detecting rapid diagnostic test (Ab-RDT) with high accuracy^a^).
4. Studies will be included if they report on individuals with previously confirmed infection that have documented vaccination (partially, fully, or boosted), as defined in the randomized controlled trials for each vaccine.
5. Partial vaccination will be defined as ≥14 days after a single dose of Pfizer/BioNTech-Comirnaty, <7 days from the second dose for Pfizer/BioNTech-Comirnaty (BNT162b2), ≥14 days after a single dose of AstraZeneca-Vaxzevria, <14 days from the second dose for AstraZeneca-Vaxzevria, ≥14 days after a single dose of Moderna-mRNA-1273, <14 days from the second dose of Moderna-mRNA-1273, <14 days from the first dose of Janssen-Ad26.COV2.S, and > 14 days after a single dose of Sinovac-CoronaVac.
6. Full vaccination will be defined as >7 days from the second dose for Pfizer/BioNTech-Comirnaty, >14 days from the first dose of Janssen-Ad26.COV2.S, >14 days from the second dose for AstraZeneca-Vaxzevria, Moderna-mRNA-1273, or Sinovac-CoronaVac.
7. Booster vaccination one will be defined as >=7 days from an additional dose after full vaccination.
8. Booster vaccination two will be defined as >=7 days from an additional dose after booster vaccination one.

Comparison Group

1. no previous vaccinations and no previously confirmed SARS-CoV-2 infection defined using WHO criteria;
2. previously confirmed SARS-CoV-2 infection defined using WHO criteria; 3. partial vaccination (defined above);
3. full vaccination (defined above);
4. booster vaccination (defined above). Outcome
5. SARS-CoV-2 reinfection defined as a possible, probable, or confirmed reinfection case according to the following criteria, adapted from WHO case definitions.
6. Possible reinfection case will be defined as NAAT or AgRDT SARS-CoV-2 positive case with a history of a primary SARS-CoV-2 infection diagnosed by serology, with at least 60 days between the positive serology test and the subsequent positive NAAT or AgRDT.
7. Probable reinfection case will be defined as NAAT or AgRDT SARS-CoV-2 positive case with a history of a primary SARS-CoV-2 infection diagnosed by NAAT or AgRDT, with at least 90 days between the episodes. Alternatively, genomic evidence for the second episode is available and includes lineage that was not submitted to SARS-Cov-2 genomic databases at the time of first infection.
8. Confirmed reinfection case will be defined as two PCR positive episodes supported by viral genomic data from both episodes of infection revealing different Pango lineages. If viral genomic data reveal two distinct Pango lineages this will qualify as adequate evidence to confirm reinfection, regardless of the time elapsed between the two episodes.

Study Design

1. Test-negative case-control, traditional case-control, cross-sectional, cohort, non-randomized controlled trials, and randomized controlled trials.

Type of literature

1. Published peer-reviewed research articles, preprints, and grey literature in any language. We will prioritize peer-reviewed versions of articles for inclusion and analysis in instances where pre-print versions of peer-reviewed articles are available.

##### Exclusion Criteria (if any met then exclude)

Population

1. N/A

Exposure Group

1. No evidence of prior confirmed case. No information on the timing, brand, or dose number for the vaccination in hybrid immunity studies.

Comparison Group 1. N/A

Outcome

1. Prior infection studies not reporting the period of time between primary infection and reinfection such that determining reinfection according to the inclusion criteria is not possible.
2. Hybrid immunity studies not reporting the period of time between either the determination of primary infection or vaccination.

Study Design

1. Case reports, case series, incomplete randomized controlled trials, and review papers. Type of literature
2. Media, news stories, and conference abstracts.

### Original author protocol

SARS-CoV-2 protective effectiveness of prior infection and hybrid immunity: a systematic review protocol

2.2 Study inclusion and exclusion criteria

**Table 1.** Inclusion criteria.

| Table 1. Inclusion criteria |  |
| --- | --- |
| Population | Humans of any age, in any geographical setting. |
| **Exposure group** | Confirmed case of SARS-CoV-2 infection with or without COVID-19 vaccination.<br>SARS-CoV-2 infection will be defined as a **confirmed case** according to the following criteria, adapted from WHO case definitions<sup>[1]</sup> (positive nucleic acid amplification test (NAAT) according to laboratory records or self report, positive SARS-CoV-2 antigen rapid diagnostic test (AgRDT)<sup>a</sup> with high accuracy according to laboratory records or self report, or a positive serology test from a lab-based assay (i.e. CLIA/ELISA) or an antibody-detecting rapid diagnostic test (Ab-RDT) with high accuracy<sup>a</sup>).<br>Studies will be included if they report on individuals with previously confirmed infection that have documented vaccination (partially, fully, or boosted), as defined in the randomized controlled trials for each vaccine.<br>**Partial vaccination** will be defined as $\geq 14$ days after a single dose of Pfizer/BioNTech-Comirnaty, $< 7$ days from the second dose for Pfizer/BioNTech-Comirnaty (BNT162b2), $\geq 14$ days after a single dose of AstraZeneca-Vaxzevria, $< 14$ days from the second dose for AstraZeneca-Vaxzevria, $\geq 14$ days after a single dose of Moderna-mRNA-1273, $< 14$ days from the second dose of Moderna-mRNA-1273, $< 14$ days from the first dose of Janssen-Ad26.COV2.S, and $\geq 14$ days after a single dose of Sinovac-CoronaVac.<br>**Full vaccination** will be defined as $> 7$ days from the second dose for Pfizer/BioNTech-Comirnaty, $> 14$ days from the first dose of Janssen-Ad26.COV2.S, $> 14$ days from the second dose for AstraZeneca-Vaxzevria, Moderna-mRNA-1273, or Sinovac-CoronaVac.<br>**Booster vaccination one** will be defined as $\geq 7$ days from an additional dose after full vaccination.<br>**Booster vaccination two** will be defined as $\geq 7$ days from an additional dose after booster vaccination one. |
| **Comparison group** | Five comparison groups will be eligible:<br>(1) no previous vaccinations and no previously confirmed SARS-CoV-2 infection defined using WHO criteria;<br>(2) previously confirmed SARS-CoV-2 infection defined using WHO criteria;<br>(3) partial vaccination (defined above);<br>(4) full vaccination (defined above);<br>(5) booster vaccination (defined above). |
| **Outcome** | SARS-CoV-2 reinfection defined as a possible, probable, or confirmed reinfection case according to the following criteria, adapted from WHO case definitions.<br>**Possible reinfection case** will be defined as NAAT or AgRDT SARS-CoV-2 positive case with a history of a primary SARS-CoV-2 infection diagnosed by serology, with at least 60 days between the positive serology test and the subsequent positive NAAT or AgRDT.<br>**Probable reinfection case** will be defined as NAAT or AgRDT SARS-CoV-2 positive case with a history of a primary SARS-CoV-2 infection diagnosed by NAAT or AgRDT, with at least 90 days between the episodes. Alternatively, genomic evidence for the second episode is available and includes lineage that was not submitted to SARS-Cov-2 genomic databases at the time of first infection.<br>**Confirmed reinfection case** will be defined as two PCR positive episodes supported by viral genomic data from both episodes of infection revealing different Pango lineages. If viral genomic data reveal two distinct Pango lineages this will qualify as adequate evidence to confirm reinfection, regardless of the time elapsed between the two episodes. |
| **Study design** | Test-negative case-control, traditional case-control, cross-sectional, cohort, non-randomized controlled trials, and randomized controlled trials. |
| **Type of literature** | Published peer-reviewed research articles, preprints, and grey literature in any language. We will prioritize peer-reviewed versions of articles for inclusion and analysis in instances where pre-print versions of peer-reviewed articles are available. |
| <sup>a</sup>For AgRDT ≥80% sensitivity and ≥97% specificity of test, compared to NAAT, in suspected cases of infection; For AbRDT, ≥90% sensitivity (>14 days post symptom onset) and ≥97% specificity, compared to a reference lab-based test, in suspected cases of infection. |  |

**Table 2.** Exclusion criteria.

| **Table 2. Exclusion criteria** |  |
| --- | --- |
| **Population** | N/A |
| **Exposure group** | No evidence of prior confirmed case. No information on the timing, brand, or dose number for the vaccination in hybrid immunity studies. |
| **Comparison group** | N/A |
| **Outcome** | Prior infection studies not reporting the period of time between primary infection and reinfection such that determining reinfection according to the inclusion criteria is not possible. Hybrid immunity studies not reporting the period of time between either the determination of primary infection or vaccination. |
| **Study design** | Case reports, case series, incomplete randomized controlled trials, and review papers. |
| **Type of literature** | Media, news stories, and conference abstracts. |

### PA-Testing

#### Simplified Objectives + Inclusion/Exclusion Criteria

The purpose of this study was to assess the characteristics of confirmatory tests for primary aldosteronism (PA) and to interpret these in the context of study design and potential risks of bias.

The following is an excerpt of two sets of criteria. A study is considered included if it meets all the inclusion criteria. If a study meets any of the exclusion criteria, it should be excluded. Here are the two sets of criteria:

##### Inclusion Criteria (all must be fulfilled)

1. The study is about primary aldosteronism (PA). Please note that primary hyperaldosteronism is a synonym.
2. The study examined at least one of the guideline-recommended confirmatory tests for PA. We are interested in the saline infusion test (SIT), oral salt loading test (SLT), fludrocortisone suppression test (FST), and the captopril challenge test (CCT). Please note some may have synonyms (e.g., SIT = intravenous saline suppression test [IVSS]).
3. Research articles reporting original data
4. The study is conducted in humans of any age 5. The study is in the English language
5. The performance of the confirmatory test(s) was compared with an independent reference standard A reference standard needs to be present to verify disease status. These may include: (i) clinical response to treatment (adrenalectomy and/or medical therapy), (ii) adrenal vein sampling results, (iii) histopathology, (iv) or another confirmatory test.
6. The confirmatory test was used to diagnose PA, rather than exclusively for subtyping. We are interested in knowing how confirmatory testing works for diagnosing PA, not just subtyping. If the confirmatory test was for subtyping, it is still possible to compare how many people had PA vs. non-PA.
7. The data (as published) is extractable for a 2×2 table (TP, FP, FN, TN). If the data are reported in any of the following formats, a 2×2 table can be reconstructed:(i) 2×2 table given, (ii) TP, FP, FN, TN rates given, (iii) Total number of patients (with disease) and total number of study subjects (with and without disease) is known, and corresponding sensitivity and specificity are given.

##### Exclusion criteria (if any met then exclude)

1. No mention about primary aldosteronism (PA)
2. Use of confirmatory tests that fall outside of guide-line recommendations 3. Non-human (e.g., in silico, animal, in vitro).
3. Conference abstracts, reviews (systematic reviews and narrative reviews), editorials, protocols, and secondary publications (data already published in another study).
4. No comparison of confirmatory test performance with a reference standard.
5. Confirmatory test was used only for subtyping, and unable to compare cases of PA vs. non-PA
6. Data not extractable for 2×2 table

#### Original author protocol

**Confirmatory Testing in Primary Aldosteronism Systematic Review Reference Sheet**

**(Version: May 28, 2021)**

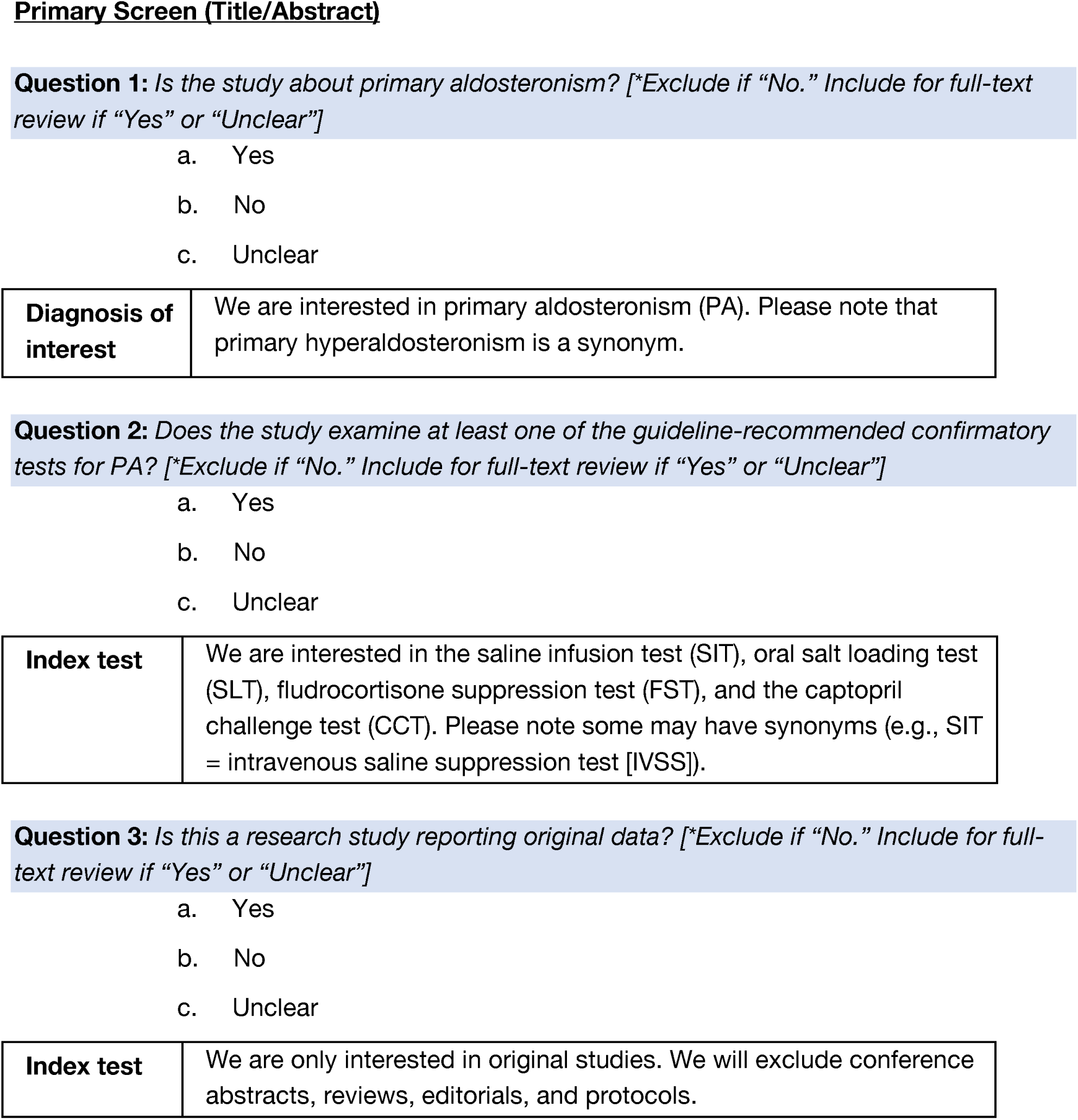
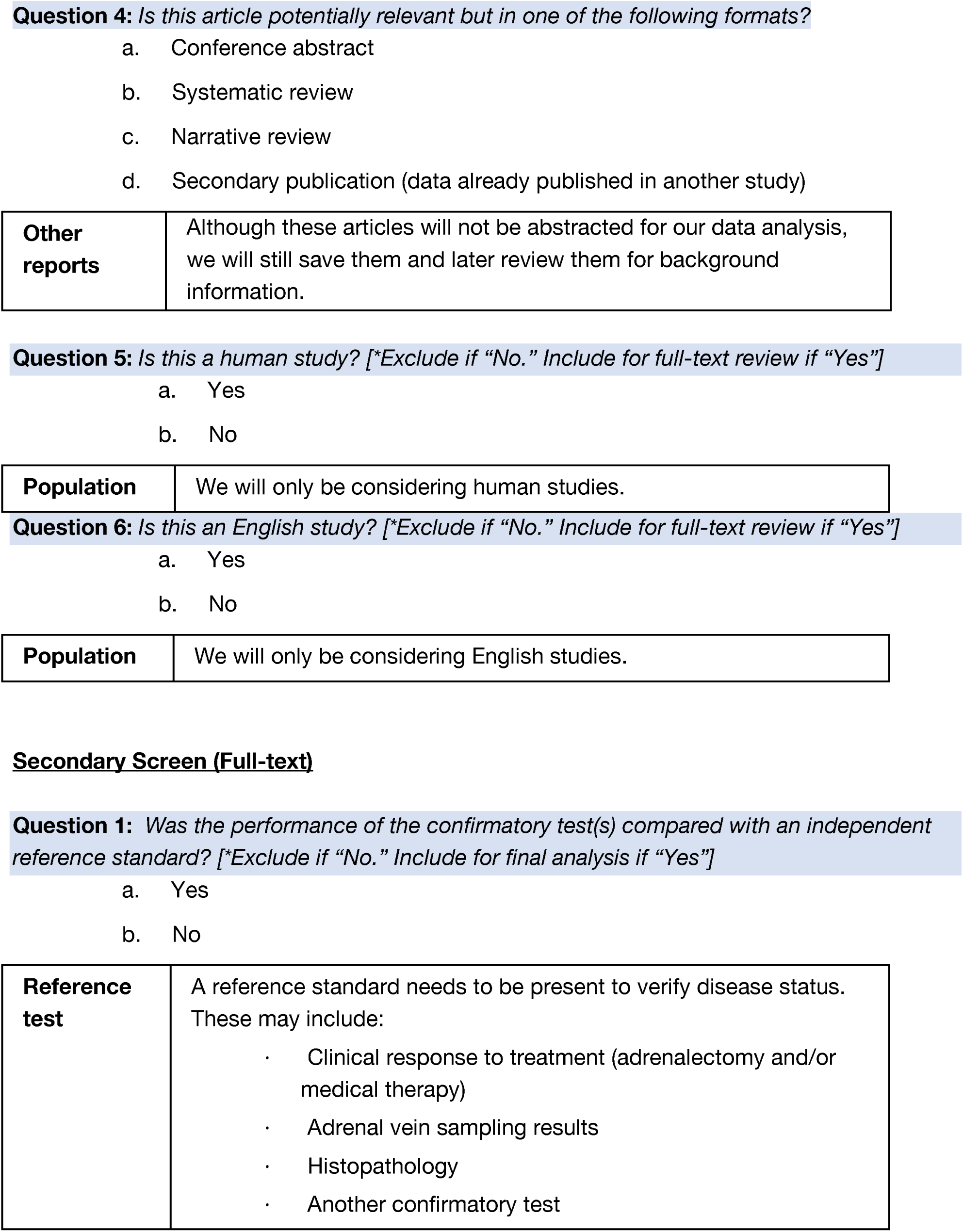
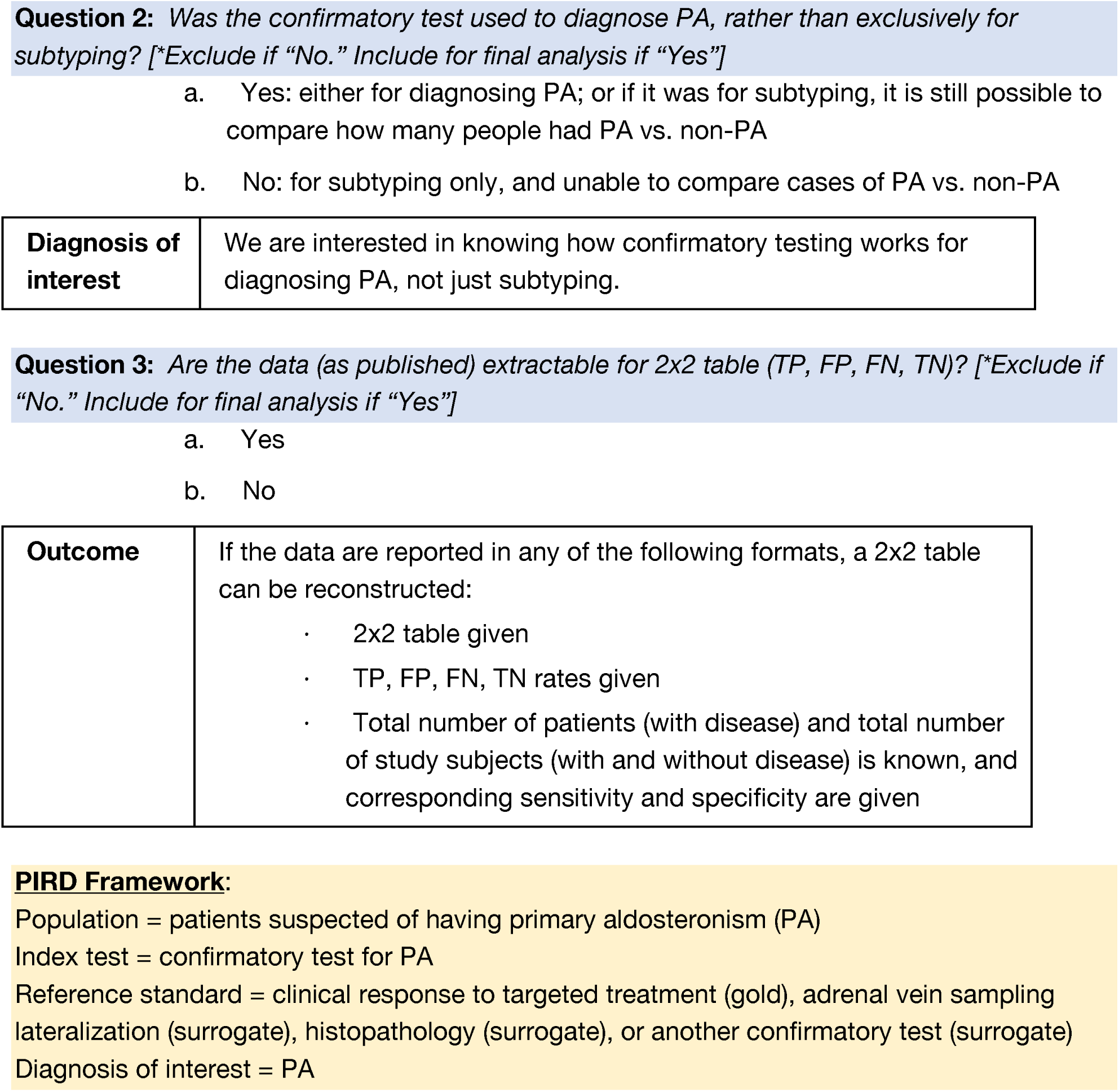

|  |  |
| --- | --- |
| **Diagnosis of interest** | We are interested in primary aldosteronism (PA). Please note that primary hyperaldosteronism is a synonym. |
| **Index test** | We are only interested in original studies. We will exclude conference abstracts, reviews, editorials, and protocols. |
| **Other reports** | Although these articles will not be abstracted for our data analysis, we will still save them and later review them for background information. |
| **Population** | We will only be considering human studies. |
| **Population** | We will only be considering English studies. |
| **Reference test** | A reference standard needs to be present to verify disease status. These may include:<br>· Clinical response to treatment (adrenalectomy and/or medical therapy)<br>· Adrenal vein sampling results<br>· Histopathology<br>· Another confirmatory test |
| **Diagnosis of interest** | We are interested in knowing how confirmatory testing works for diagnosing PA, not just subtyping. |
| **Outcome** | If the data are reported in any of the following formats, a 2x2 table can be reconstructed:<br>· 2x2 table given<br>· TP, FP, FN, TN rates given<br>· Total number of patients (with disease) and total number of study subjects (with and without disease) is known, and corresponding sensitivity and specificity are given |

### PA-Outcomes

#### Simplified Objectives and Inclusion/Exclusion Criteria

We aimed to conduct a meta-analysis to examine the clinical outcomes of surgery vs medical therapy with respect to mortality, composite major adverse cardiovascular events (MACE, and its individual components), progression to chronic kidney disease, and incident diabetes mellitus.

The following is an excerpt of two sets of criteria. A study is considered included if it meets all the inclusion criteria. If a study meets any of the exclusion criteria, it should be excluded. Here are the two sets of criteria:

##### Inclusion Criteria (all must be fulfilled)

1. The study is about primary aldosteronism (PA). Please note that primary hyperaldosteronism is a synonym.
2. Reports surgery (adrenalectomy) or medication (mineralocorticoid receptor antagonists, spironolactone, eplerenone) treatment for primary aldosteronism.
3. Reports mortality, MACE, ACS, stroke, arrhythmia, heart failure, chronic kidney disease, and/or incident diabetes clinical outcomes after treatment.
4. Research articles reporting original data.
5. Randomized clinical trials, cohort studies, and cross-sectional studies.
6. The study is conducted in humans of any age.
7. The study is in the English language.

##### Exclusion criteria (if any met then exclude)

1. Does not report about primary aldosteronism (PA). 2. Does not report treatment for primary aldosteronism.
2. Does not report mortality, major adverse cardiovascular events (MACE), acute coronary syndrome (ACS), stroke, arrhythmia, heart failure, chronic kidney disease, and/or incident diabetes clinical outcomes after treatment.
3. Non-human (e.g., in silico, animal, in vitro).
4. Conference abstracts, case reports, case series, reviews (systematic reviews and narrative reviews), editorials, protocols, and secondary publications (data already published in another study).

#### Original author protocol

**Primary Aldosteronism Treatment Response: Systematic Review Reference Sheet**

**(Version: June 1, 2022 revised)**

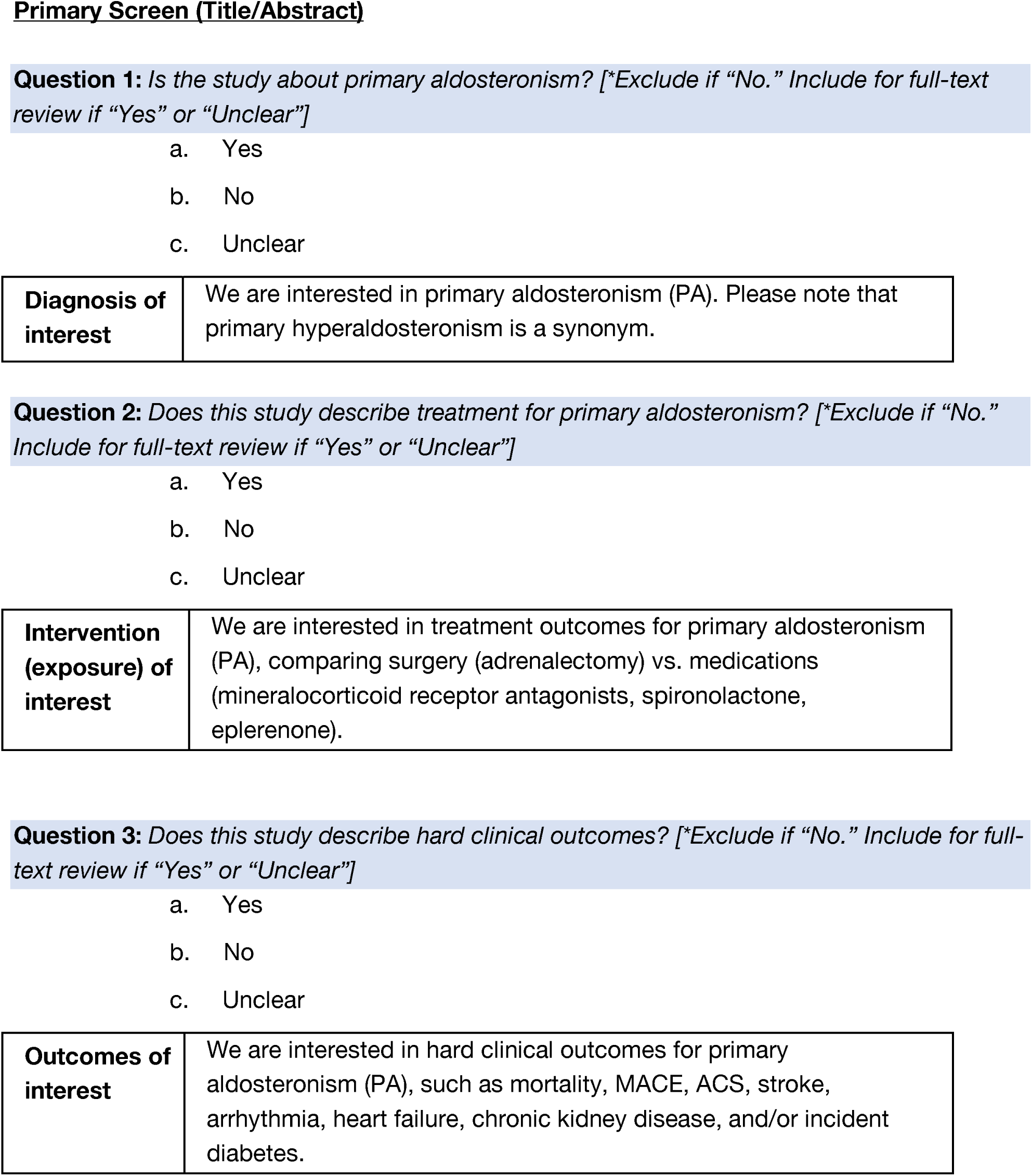
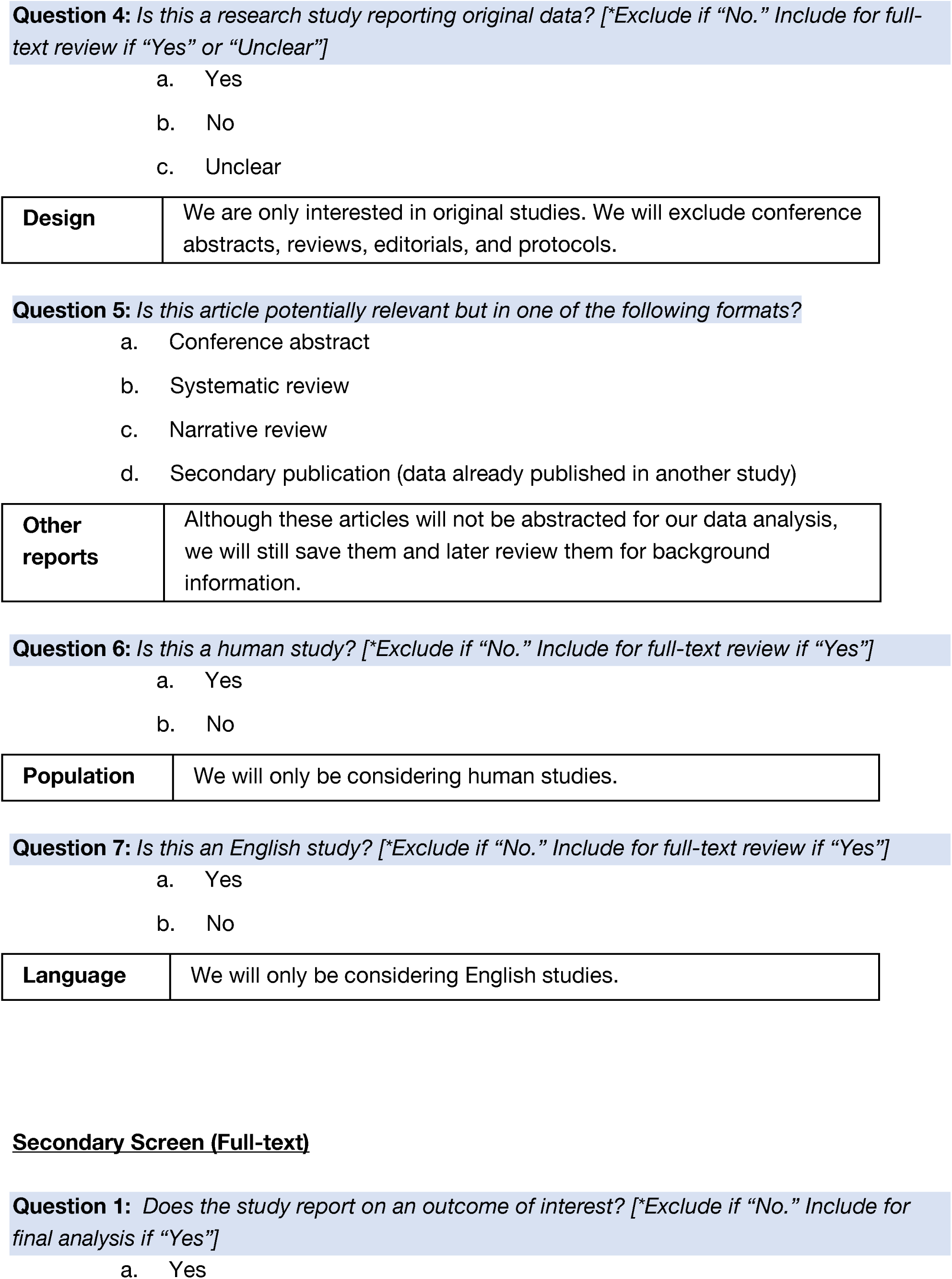
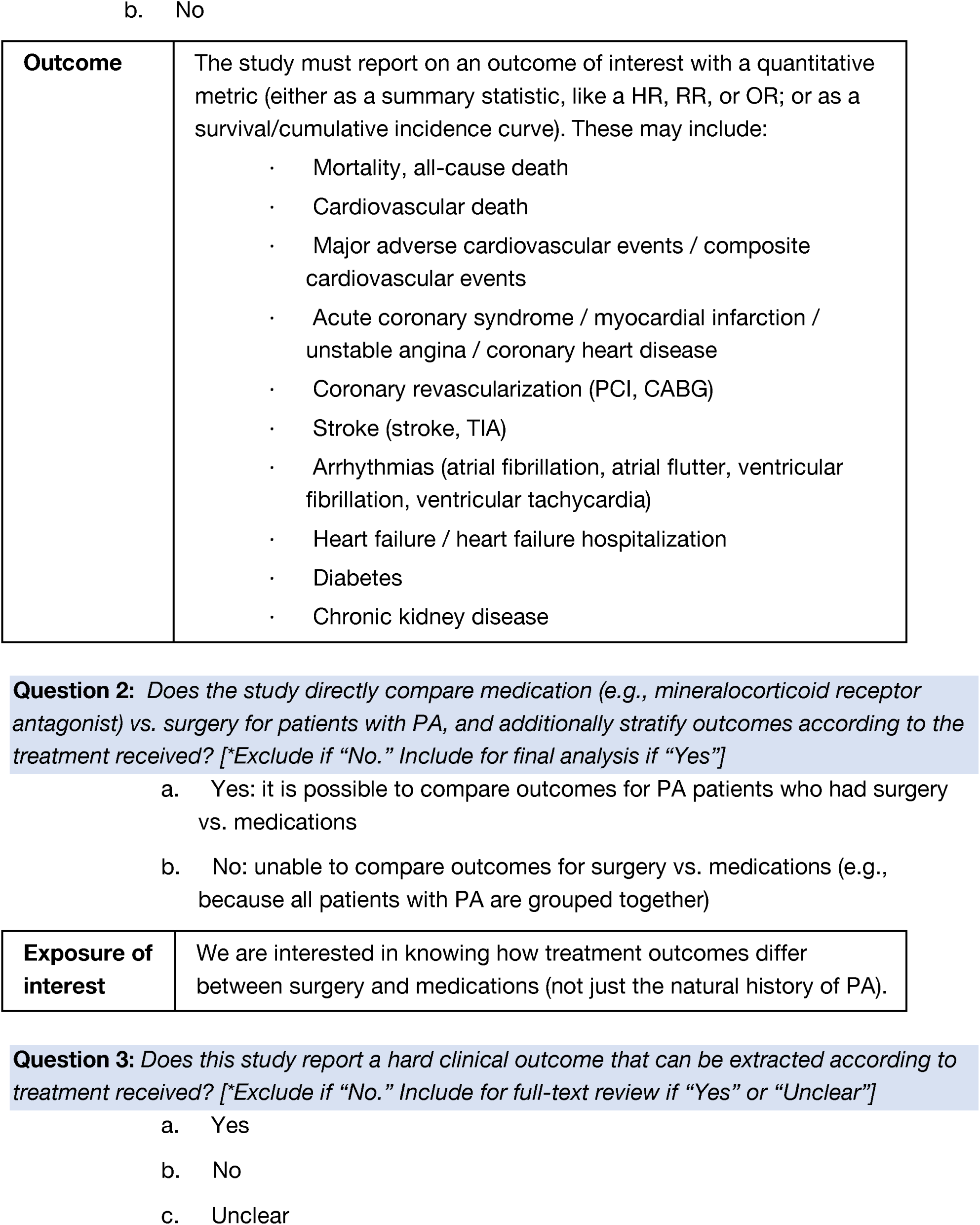
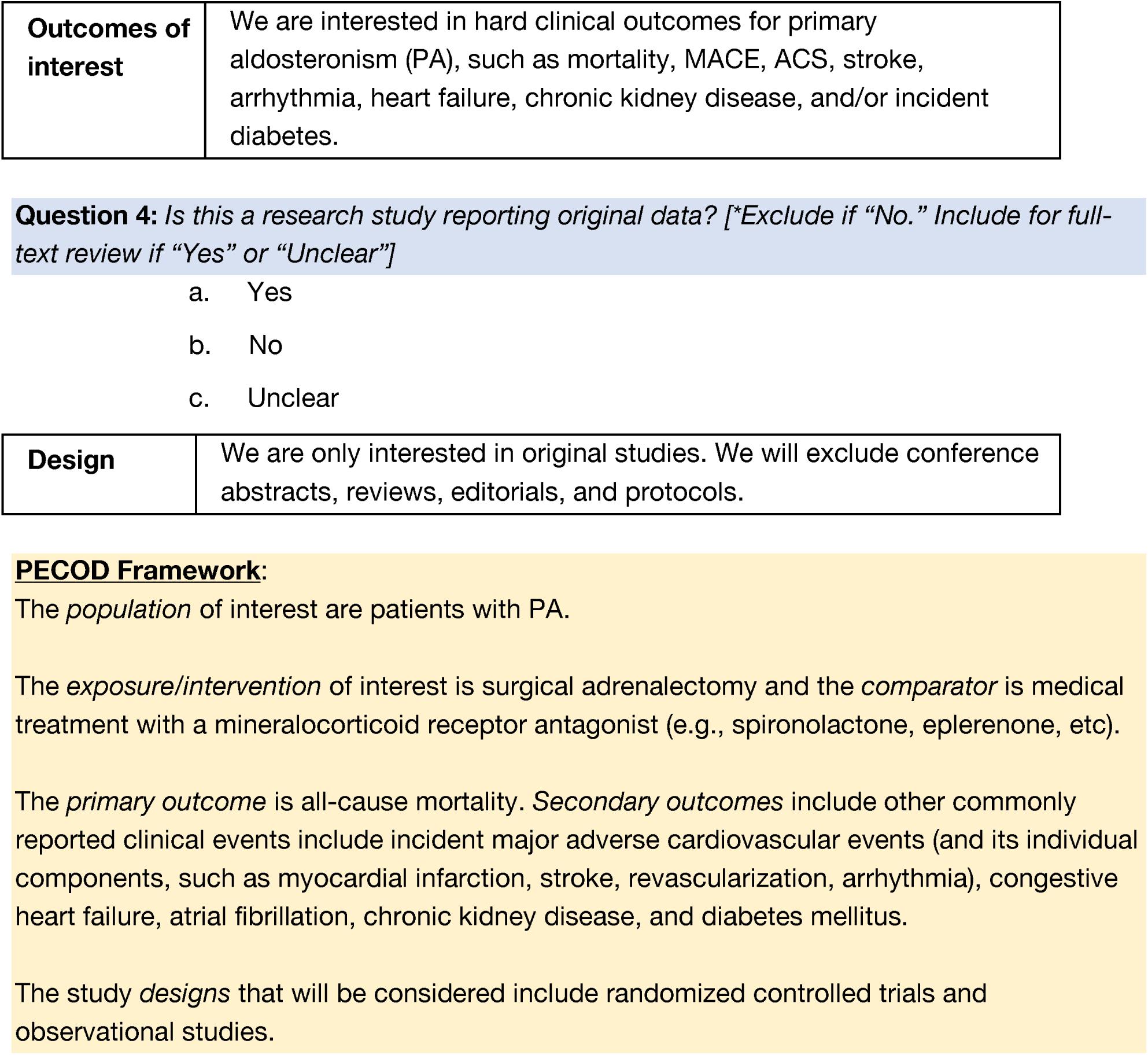

|  |  |
| --- | --- |
| **Diagnosis of interest** | We are interested in primary aldosteronism (PA). Please note that primary hyperaldosteronism is a synonym. |
| **Intervention (exposure) of interest** | We are interested in treatment outcomes for primary aldosteronism (PA), comparing surgery (adrenalectomy) vs. medications (mineralocorticoid receptor antagonists, spironolactone, eplerenone). |
| **Outcomes of interest** | We are interested in hard clinical outcomes for primary aldosteronism (PA), such as mortality, MACE, ACS, stroke, arrhythmia, heart failure, chronic kidney disease, and/or incident diabetes. |
| **Design** | We are only interested in original studies. We will exclude conference abstracts, reviews, editorials, and protocols. |
| **Other reports** | Although these articles will not be abstracted for our data analysis, we will still save them and later review them for background information. |
| **Population** | We will only be considering human studies. |
| **Language** | We will only be considering English studies. |
| **Outcome** | The study must report on an outcome of interest with a quantitative metric (either as a summary statistic, like a HR, RR, or OR; or as a survival/cumulative incidence curve). These may include:<br>• Mortality, all-cause death<br>• Cardiovascular death<br>• Major adverse cardiovascular events / composite cardiovascular events<br>• Acute coronary syndrome / myocardial infarction / unstable angina / coronary heart disease<br>• Coronary revascularization (PCI, CABG)<br>• Stroke (stroke, TIA)<br>• Arrhythmias (atrial fibrillation, atrial flutter, ventricular fibrillation, ventricular tachycardia)<br>• Heart failure / heart failure hospitalization<br>• Diabetes<br>• Chronic kidney disease |
| **Exposure of interest** | We are interested in knowing how treatment outcomes differ between surgery and medications (not just the natural history of PA). |
| **Outcomes of interest** | We are interested in hard clinical outcomes for primary aldosteronism (PA), such as mortality, MACE, ACS, stroke, arrhythmia, heart failure, chronic kidney disease, and/or incident diabetes. |
| **Design** | We are only interested in original studies. We will exclude conference abstracts, reviews, editorials, and protocols. |

### SVCF

#### Simplified Objectives + Inclusion/Exclusion Criteria

Objectives: To evaluate the association of low SVC flow, diagnosed in the first 48 hours after birth echocardiography, with neurological morbidity and mortality, among very preterm neonates.

The following is an excerpt of two sets of criteria. A study is considered included if it meets all the inclusion criteria. If a study meets any of the exclusion criteria, it should be excluded. Here are the two sets of criteria:

##### Inclusion Criteria (all must be fulfilled)

1. Preterm infants <32 weeks gestational age who had echocardiography done within the first 48 hours after birth with evaluation of SVC flow

A. Prognostic factor: Low SVC flow identified by Doppler assessment during echocardiography performed in the first 48 hours after birth.
B. Comparison: Normal SVC Flow
2. Reported outcomes including intraventricular hemorrhage (IVH), presence of periventricular leukomalacia (PVL), all-cause mortality before discharge from neonatal intensive care unit (NICU), neurodevelopmental impairment in early childhood or any diagnosed Cerebral Palsy, visual and/or hearing deficits, or necrotizing enterocolitis (NEC)

A. Any grade IVH diagnosed in the first 7 days after birth by cranial ultrasonography
B. Severe IVH (defined as stage 3 or higher according to Papile’s classification 14) diagnosed in the first 7 days after birth
3. Randomized controlled trials, cohort or case-control studies

##### Exclusion criteria (if any met then exclude)

1. Non-human (e.g., in silico, animal, in vitro)
2. Cross-sectional studies, narrative reviews, case series or case reports

#### Original author protocol

<u>Association of early-life low superior vena cava flow among preterm neonates and death or cerebral haemorrhage: a systematic review and meta-analysis: Protocol</u>

The objective of this study is to systematically review and meta—analyse the association of low SVC flow during the transitional period among preterm neonates < 32 weeks GA with mortalityand adverse neurological morbidity..

##### PICO/PFO outline-

###### Population

Preterm infants <32 weeks gestational age

###### Prognostic factor

Low SVC flow identified by Doppler assessment during echocardiography performed in the first 48 hours after birth (measured as ml/kg/min)

###### Comparison

Patients with normal SVC flow

###### Outcomes

Primary- **Any grade IVH** diagnosed in the first 7 days of life by cranial ultrasonography Secondary-

[1 Severe IVH (defined as stage 3 or higher according to Papile’s classification ^14^) diagnosed in the first 7 days of life,

[1 Presence of PVL (diagnosed by 28 days of life)

[1 Mortality within the neonatal period (defined as the first 28 days of life)

[1 Neurodevelopmental impairment in early childhood (Defined as a composite outcome of any of the following: Cerebral palsy with Gross Motor Function Classification System score >=1 or Bayley-III motor composite <85; Bayley cognitive composite < 85; Bayley language composite <85; Any sensorineural/mixed hearing loss; Any unilateral or bilateral visual impairment)^15^

[1 Necrotizing Enterocolitis (NEC) compared between low and normal SVC flow groups

## METHODOLOGY

This systematic review and meta-analysis will be conducted according to the PRISMA guidelines and Cochrane methodology.

### Eligibility Criteria

Randomized controlled trials, cohort or case-control studies that evaluated the following population characteristics will be included for this study:

Preterm infants <32 weeks gestational age who had echocardiography done within the first 48 hours after birth with evaluation of SVC flow. Studies must have compared any of the above stated clinical outcomes among preterm neonates with low vs. normal SVC flow to be considered for inclusion. Studies will be included only if they exclusively include human subjects and there will be no language limitations.

#### Exclusion criteria

Cross-sectional studies, narrative reviews, case series or case reports on this topic will be excluded. We will also exclude any animal studies. Any identified studies without any available full text from the included databases or the original authors will also be excluded.

